# Classifying and Differentiating Clinical Coding of Respiratory Syncytial Virus, Influenza, and COVID-19 in OpenSAFELY Between 2016 and 2024

**DOI:** 10.64898/2026.04.09.26350495

**Authors:** Em Prestige, Charlotte Warren-Gash, Jennifer K Quint, Dave Evans, Ruth E Costello, Amir Mehrkar, Seb Bacon, Ben Goldacre, Sarah Barley-McMullen, Farheen Yameen, Michael Natt, Yvonne Alder, William J. Hulme, Edward P. K. Parker, Rosalind M. Eggo

**Affiliations:** Faculty of Epidemiology and Population Health, London School of Hygiene & Tropical Medicine, Keppel St, London. WC1E 7HT, United Kingdom; Faculty of Medicine, School of Public Health, Imperial College London, London, United Kingdom; Bennett Institute for Applied Data Science, Oxford University, Oxford, United Kingdom; Winter Pressures PPIE Advisory Board, London School of Hygiene & Tropical Medicine, Keppel St, London. WC1E 7HT, United Kingdom

**Author notes:** indicates equal contribution (William J. Hulme, Edward P. K. Parker, Rosalind M. Eggo).

**Keywords:** electronic health records, phenotype, identification, classification, respiratory viruses, Covid-19, influenza, RSV

## Abstract

Electronic health records (EHRs) are a rich source of data which can be used to analyse health outcomes using computable phenotypes. With the approval of NHS England we used the OpenSAFELY secure analytics platform to design and assess phenotypes to classify three key respiratory viruses – respiratory syncytial virus (RSV), influenza, and COVID-19 – in English coded health data between September 2016 and August 2024. We compared narrow and broad phenotypes to one another and to publicly available surveillance data. Cases from both phenotypes showed similar seasonal patterns to surveillance data. Broad phenotypes led to increased risk of misclassification than narrow phenotypes for mild cases. For severe cases the risk of misclassification was higher in infants than for older adults, irrespective of the phenotype used. The phenotypes presented here offer a potential route to classify respiratory viruses from coded health records in the absence of testing information.

## Background

Respiratory Syncytial Virus (RSV), influenza, and COVID-19 are major respiratory viruses and key drivers of healthcare utilisation (1–8), but often present with a similar upper respiratory tract infection (URTI) syndrome. Electronic Health Records (EHRs) provide a rich source of information on these viruses, their burden on healthcare systems, and the risk factors for disease. However, the viruses are rarely tested for outside of secondary care - excepting extraordinary circumstances, such as the widespread testing during the COVID-19 state of emergency. Additionally, while molecular testing is regularly conducted in hospital settings, test results are rarely coded in EHRs, making them unavailable for research. The absence of testing data presents a key challenge to accurately classifying and differentiating RSV, influenza, and COVID-19 in EHRs. The ability to detect and distinguish the clinical coding of these viruses could broaden research capabilities and improve outbreak detection.

When individuals present to healthcare providers, such as general practitioners (GPs) and hospitals, their consultations are recorded with clinical codes (9). Clinical codes can correspond to specific conditions, symptoms or prescriptions. In secondary care settings in the UK, hospital episodes are classified using ICD-10 codes (10), which can denote pathogen-specific diagnoses (e.g. RSV), diseases (e.g. bronchiolitis), or pathogen-associated presentations (e.g. unspecified lower respiratory tract infections (LRTIs)). In primary care settings, clinical events are recorded using a much more diverse system of SNOMED-CT codes (11), again ranging from highly specific diagnoses to broader symptoms.

Current methods of investigating respiratory viruses often rely on using testing data from surveillance systems, for which reporting varies both over time and between viruses (12). Many surveillance systems are reported at an aggregate level and as such cannot be used to investigate individual-level risk factors for disease; those that are individual-level use a smaller sample of practices when compared to broader EHR systems. We aimed to develop a series of computable phenotypes, comprising codelists and corresponding sets of logical rules (13), to classify individuals with RSV, influenza, and COVID-19 in EHRs. For each virus, we developed:

(i) phenotypes designed to be specific to each virus; (ii) phenotypes intended to capture cases relating to each virus, with greater sensitivity (but potentially lower specificity); and (iii) phenotypes devised to capture overall respiratory burden which may be attributed to any of the three viruses. We then evaluated the ability of these phenotypes to classify and differentiate cases in primary and secondary care settings. These phenotypes have scope to be used for: (i) real-time surveillance purposes; (ii) regular reporting on trends for service planning and monitoring; and (iii) epidemiologic research into the impact of new interventions or service disruptions.

## Methods

### Data Source

Through the OpenSAFELY analytics platform we extracted linked GP, hospital admissions, and A&E data for 42% of the population of England. All practices running SystmOne software made by the health technology company TPP were included, which includes over 2500 practices. The patient population was representative of the general population with some regional variations (14). We used consultations within primary care, secondary care (Admitted Patient Care Spells - APCS), and emergency care (Emergency Care Data Set - ECDS).

Emergency care attendances were available from late 2017, but were incomplete until late 2019 (15). We extracted information on prescriptions from primary care EHRs. Ethnicity from primary care records was supplemented with ethnicity recorded in secondary care due to missingness in ethnicity records. OpenSAFELY requires all outputs to undergo statistical disclosure control. As such, all results have been rounded to the nearest midpoint-10; all true zeroes have been retained.

### Patient and Public Involvement

A panel of six patient and public members were involved during framing of the research questions and how the results were presented. The accessibility of these results has been improved through the input of the panel. Panel members vary by age, gender, ethnicity, socioeconomic status, chronic illness/disability, and lived experience of respiratory virus infections personally or as a carer.

### Study Population

We split the study population into four age cohorts (older adults, 65 years or over; adults, 18-64 years; children and young people, 2-17 years; and infants, 0-2 years). We extracted eight annual cohorts from 2016 to 2024, with each cohort including data from 1 September to 31 August of the subsequent year. Individuals were included in any annual cohort for which they met the inclusion criteria of 3 months of continuous registration prior to follow-up; infants were included from birth if they were registered within 3 months of birth. We also explored a subgroup of infants who could be linked to their birthing parent; linked infants were included if their birthing parent had at least 1 year of continuous registration prior to birth.

Follow-up continued until the first of: death, practice deregistration, aging out of age cohort, or end of annual cohort (31 August). Individuals were excluded from the study if they had missing information on age, sex, or IMD quintile, due to potential poor data quality. Care residents (using a care home flag within TPP) were also excluded as this setting is not representative of the epidemiological burden in the general population. Infants were excluded if they were eligible for Palivizumab due to high risk of complications due to RSV (16) (via the presence of ventilation codes for those under 12 months or those 12-23 months with cardiovascular-related diagnosis codes).

### Respiratory Virus Phenotypes

We defined virus phenotypes which applied across all cohorts with specific criteria for infants and older adults. We developed the phenotypes in collaboration with medical professionals in respiratory and primary care. To classify and differentiate cases of each respiratory virus, we created two phenotypes: one narrow which aimed to capture cases more likely to be attributable to specific viruses; and one more broad, aiming to capture as many relevant cases as possible. In addition, we separated cases into mild or severe, where mild was primary or emergency care (where relevant) and severe was in secondary care (Hospital Episode Statistics - HES). To enforce temporal ordering of case severity, individuals could be classified as both mild and severe cases, with follow-up for mild cases stopped if a severe case occurred - with respect to each virus. We also created an overall respiratory virus definition, which aimed to capture the overall burden of the three respiratory viruses of interest, without distinguishing the pathogen driving it. All phenotypes used a pre-specified episode length definition of 14 days. For all phenotypes, case classification was separate for each pathogen, such that cases could be counted as multiple pathogens simultaneously. Overlapping cases were included in virus-specific case counts.For both mild and severe phenotypes, the narrow phenotypes were a subset of the broad phenotypes for each virus and severity level (supplementary Table S1, Figure 1A). As such, all cases classified using narrow phenotypes were also classified using the broad phenotypes.

**Figure 1.**
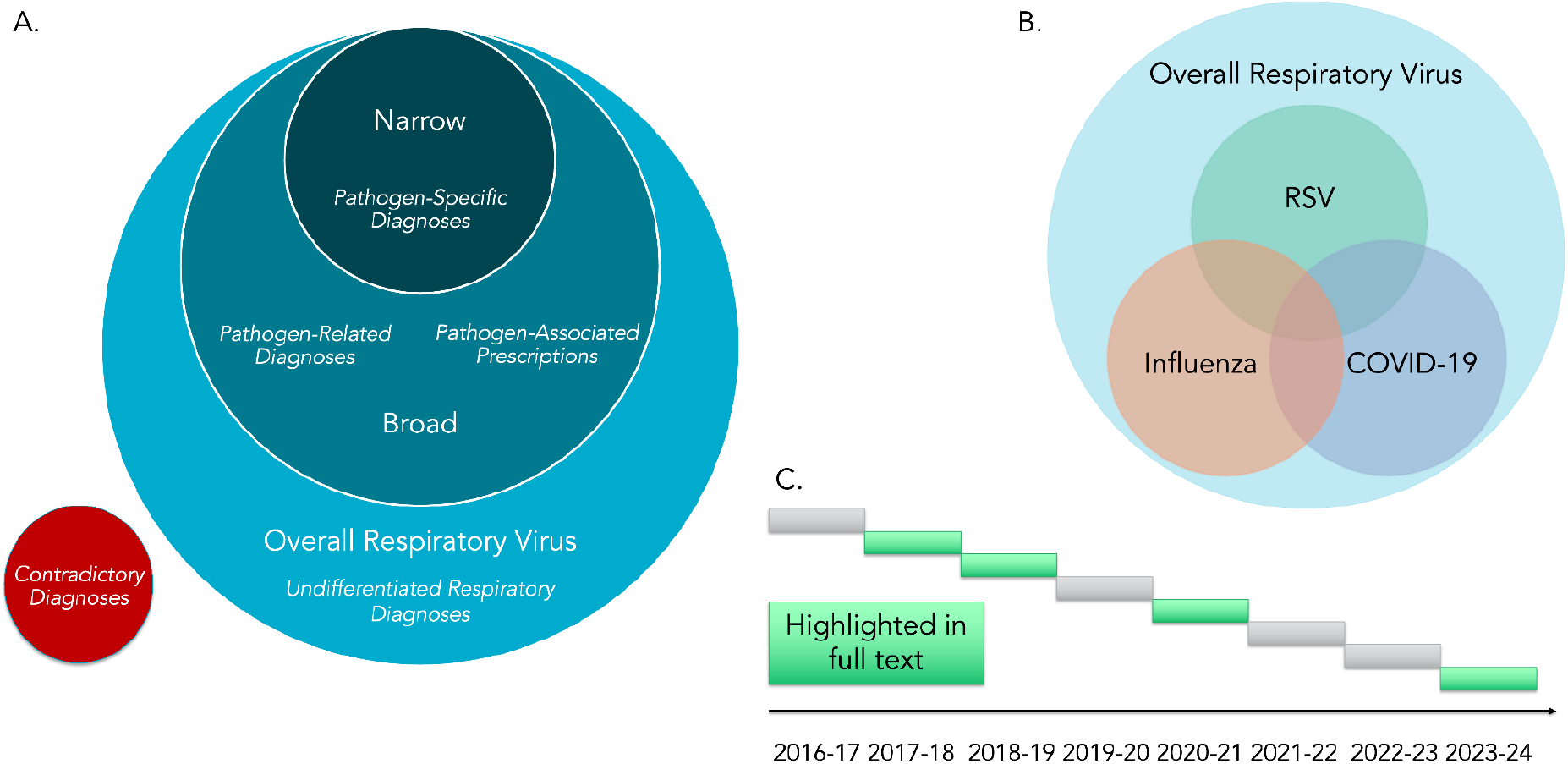
Summary of analysis structure. A: Schematic of relationship between phenotypes, showing how the narrow phenotype is a subset of the broad and so forth. Contradictory diagnoses (red) only apply to broad phenotypes. B: Cases can be assigned to more than one virus, which we term “overlap”. All virus-specific phenotyped cases are captured by the overall phenotype (light blue). C: Each of 8 annual cohorts runs from 1st September, and the annual cohorts shown in the main text are highlighted in green.

For narrow mild phenotypes, each virus included at least one pathogen-specific SNOMED CT code in the primary care record. Bronchiolitis was used as a virus-specific feature for RSV (17,18). We used emergency attendances for bronchiolitis (from ECDS) in infants as an age-specific feature, to capture mild RSV cases out-of-hours, which we limited to those with ‘discharge to home’ as the discharge destination.

For broad mild phenotypes, all phenotypes: (i) met the narrow phenotype definition; (ii) had two pathogen-associated clinical event SNOMED-CT codes, e.g. respiratory symptoms within an episode (19–22); (iii) or one prescription code (in dm+d) within the same episode as one pathogen-associated clinical event code. To improve the accuracy of broad phenotypes, we excluded individuals with codes indicative of non-target infection a month before or after a relevant event or prescription (supplementary Table S1). Age-specific features included exacerbations of COPD (23) and asthma (24) for overall respiratory virus cases in older adults. A virus-specific feature for RSV was that we did not use any prescriptions to classify cases.

Virus-specific features for influenza were: (i) the inclusion of codes indicating the ‘suspicion’ of influenza; or (ii) the presence of influenza-like-illness (ILI), adapted from the WHO definition (25), which required the presence of at least one code indicating an acute respiratory illness (ARI) **and** at least one code indicating a fever, occurring within a 14-day period. We compared our definition of ILI (see supplementary Table S1) to the definition developed, and validated, by the Royal College of General Practitioners (RCGP) (26). As both phenotypes were developed from the WHO definition of ILI (25), they were closely aligned. Minor differences were present, such as the RCGP definition using a 10-day window for ascertainment (versus our 14 days) and our combining of codes for ‘cough’ in our ARI definition. We decided not to enforce a required presence of cough, fever, and ARI due to the frequency with which codes for all three were likely to appear on records; however, we retained the ‘cough’ codes due to high usage suggested by clinical code recording data (27).

For narrow severe phenotypes, all viruses were defined as requiring a pathogen-specific ICD-10 code in first or second position of the hospital record (APCS). For broad severe phenotypes, common features across viruses included: (i) meeting the narrow phenotype definition; or (ii) the presence of a relevant pathogen-associated ICD-10 code in first or second position. To improve the accuracy of the broad phenotypes, we excluded individuals with codes indicative of non-target infection within a month prior to or proceeding the case (supplementary Table S1). Virus-specific features included the use of COPD (23) or asthma (24) exacerbations as a proxy for severe overall respiratory virus cases.

To define the phenotypes above, we used a total of 36 codelists (20 in SNOMED CT format, 14 in ICD-10 format, and 2 in dm+d format). Eight of these codelists comprised single SNOMED CT or ICD-10 codes. Six were existing codelists available on OpenCodelists (28). Two codelists were adapted by converting codelists used in CPRD (29) and supplementing the conversion by re-running the searches used to build the original codelist. Another two codelists were developed using validated search terms (30), with codes then reviewed by a clinician. The remaining 18 codelists were newly developed with clinician input. This involved working collaboratively with clinicians to develop a list of relevant search terms, and verifying the subsequent codelist. Codelists are available for re-use on opencodelists.org, with corresponding search terms, for details on codelists used see the relevant references included within supplementary Table S1.

#### Phenotype Design Sensitivity Analyses

To explore the drivers of case classifications, we examined the effect of the most commonly used codes in our broad phenotypes (27). We used openly available SNOMED-CT and ICD-10 code usage data to identify one-two codes for each of our broad phenotypes which were most commonly used. For mild phenotypes, ‘cough (finding)’ had the highest usage across all three pathogens. For severe phenotypes, the ICD-10 code (J22 ‘unspecified acute lower respiratory infection’) was the only difference between narrow and broad RSV phenotypes and so a direct comparison was possible. For influenza, we explored ICD-10 codes J069 (‘acute upper respiratory infection, unspecified’) and J219 (‘acute bronchiolitis, unspecified’). Finally, for COVID-19 we explored the ICD-10 code B972 (‘Coronavirus as the cause of diseases classified elsewhere’). We also explored the potential contribution of cases that would be additionally classified if we included antivirals (oseltamivir and zanamivir) in our RSV phenotype. Here we defined an inclusion as broad mild RSV to be the presence of a prescription with at least one pathogen-associated symptom code within two weeks of each other, and expanded our exclusion criteria to align with influenza and COVID-19 phenotypes. For each comparison, we looked at the change in the number of cases classified and reinfections occurring for infants and older adults in 2023-24.

### Internal Validation of Mild Phenotypes

To provide an internal validation of our mild phenotypes, we selected severe cases classified using our narrow phenotypes (in which pathogen-specific ICD-10 codes were likely to have been informed by in-hospital testing), then identified mild cases phenotyped in the 14 days preceding hospitalisation. We included mild cases with: (i) narrow phenotypes; (ii) broad phenotypes; (iii) the overall respiratory virus phenotypes; and (iv) an extended phenotype of broader respiratory outcomes (supplementary Section S1.1). We then quantified the percentage agreement of mild cases with the severe reference cases. The validation was stratified by annual cohort, age cohort, and by narrow versus broad mild phenotype.

#### Reinfections

We explored decisions made when designing the phenotypes, such as episode length and reinfections, using the narrow phenotypes. We quantified the proportion of cases who went on to be classified a second time with the same case-type within the same annual cohort. We then quantified the proportion of those reinfections occurring within 28 days over time and by severity. A high proportion of reinfections within 28 days would suggest that the defined episode length of 14 days may be too short. We explored this via a sensitivity analysis comparing 14-day and 28-day episode length.

#### Case Classification Types

In order to evaluate whether the phenotypes were achieving the expected differences in sensitivity we compared phenotypes to one another by calculating the change in the number of classified cases using narrow versus broad phenotypes (Figure 1A). We also quantified the change in overlap in case classification – cases simultaneously attributable to multiple viruses indicating either co-infection (particularly when using narrow phenotypes) or misclassification of cases (Figure 1B). In addition, we reviewed the proportion of cases classified as overall respiratory virus that were also classified as either RSV, influenza, or COVID-19. Below we highlight results for the groups with the highest burden of disease in the main text (older adults and infants) for annual cohorts of: highest burden (2017–18, 2018–19), initial emergence of COVID-19 (2020–21), and most recency (2023–24) (Figure 1C).

### Comparison to Surveillance Data

We compared case counts for each of our EHR phenotypes to time-series surveillance data from the WHO RespiMart (31). Data submissions to RespiMart come from national reporting systems, in this case the UKHSA, and contain information on both sentinel and non-sentinel surveillance. Countries submit laboratory-confirmed testing volumes and will be included for each virus respectively if at least one specimen per week is tested. The time-series surveillance data were provided as weekly case counts, which we aggregated to monthly counts. We also explored how our estimates compared to UKHSA data from 2016-2020 (32), through the same descriptive comparisons. UKHSA data were not available from 2020 due to changes in reporting procedures, after which only test positivity was publicly available.

We visually compared surveillance data with phenotyped cases over time to explore the alignment of timing and scale of cases classified as RSV, influenza, and COVID-19 when contrasted with testing data. We then used Pearson’s correlation coefficient (33) to ascertain levels of correlation between monthly case counts obtained by phenotyped cases and surveillance data. Correlations were calculated using data for all months (combining across the annual cohorts) and for annual cohorts separately.

## Results

### Study Population

Each annual cohort included between 21-25 million individuals (supplementary Table S2 and Figure S1-S5), of which 4-5 million were older adults (65+), 12-15 million adults (18–64), 4-5 million children and young people (2–17), and 600-700 thousand infants (<2). Of the infants, between 95-150 thousand infants each year were linked to their birthing parent. Baseline characteristics for participants remained broadly stable each year (supplementary Tables S2-7), although population-level changes over time led to fluctuations in the distributions of some characteristics such as Index of Multiple Deprivation (IMD) and age (34–36). There were some changes in ethnicity profile over time reflecting changes in the recording of ethnicity data in EHRs (37,38).

### Respiratory Virus Phenotypes

Case classification rates, per 1,000 person-years, were consistently higher when using broad phenotypes as compared to narrow phenotypes (Table 1). Across all age cohorts, the rate of cases classified as RSV has increased over time - aside from in 2020-21 and 2021-22. RSV case classification rates were highest in infants, irrespective of the phenotypes used. Case classification rates of influenza were more variable over time. When using narrow phenotypes, rates of influenza case classification were highest in older adults; using broad phenotypes led to rates of influenza classification being highest in infants. COVID-19 case classification rates were highest in 2020-21 and 2021-22 before decreasing from 2022-23 onwards. Classification rates of severe COVID-19, across all cohorts, were very similar when using both narrow and broad phenotypes. Mild COVID-19 case classification rates were highest in adults and children and young people, while severe COVID-19 case classification rates were highest in older adults.

**Table 1.** Rates of cases classified as RSV, influenza, and COVID-19 per 1,000 person-years. Rates are reported separately by virus, severity, phenotype used, age, and annual cohort.

| Older Adults |  |  |  |  |  |  |  |  |  |
| --- | --- | --- | --- | --- | --- | --- | --- | --- | --- |
| Annual Cohort |  | 2016-17 | 2017-18 | 2018-19 | 2019-20 | 2020-21 | 2021-22 | 2022-23 | 2023-24 |
| RSV |  |  |  |  |  |  |  |  |  |
| Narrow | Mild | 0.06 | 0.07 | 0.09 | 0.11 | 0.03 | 0.07 | 0.22 | 0.25 |
|  | Severe | 0.06 | 0.09 | 0.16 | 0.24 | 0.02 | 0.07 | 0.29 | 0.31 |
| Broad | Mild | 8.68 | 8.38 | 7.82 | 5.92 | 2.57 | 4.19 | 6.14 | 6.57 |
|  | Severe | 6.31 | 6.71 | 6.57 | 5.50 | 3.27 | 3.87 | 5.04 | 5.73 |
| Influenza |  |  |  |  |  |  |  |  |  |
| Narrow | Mild | 0.39 | 1.11 | 0.78 | 0.86 | 0.17 | 0.21 | 0.92 | 0.57 |
|  | Severe | 0.43 | 1.65 | 1.11 | 0.90 | 0.004 | 0.13 | 1.78 | 1.06 |
| Broad | Mild | 1.49 | 2.58 | 1.94 | 1.96 | 0.51 | 0.82 | 2.24 | 2.19 |
|  | Severe | 1.08 | 2.32 | 1.75 | 1.37 | 0.22 | 0.44 | 2.20 | 1.57 |
| COVID-19 |  |  |  |  |  |  |  |  |  |
| Narrow | Mild |  |  |  | 2.42 | 27.74 | 102.04 | 24.26 | 7.53 |
|  | Severe |  |  |  | 3.51 | 9.88 | 11.28 | 7.67 | 4.96 |
| Broad | Mild |  |  |  | 17.11 | 55.22 | 134.6 | 63.09 | 49.66 |
|  | Severe |  |  |  | 3.52 | 9.90 | 11.30 | 7.68 | 4.98 |
| Adults |  |  |  |  |  |  |  |  |  |
| Annual Cohort |  | 2016-17 | 2017-18 | 2018-19 | 2019-20 | 2020-21 | 2021-22 | 2022-23 | 2023-24 |
| RSV |  |  |  |  |  |  |  |  |  |
| Narrow | Mild | 0.02 | 0.02 | 0.03 | 0.03 | 0.01 | 0.03 | 0.06 | 0.08 |
|  | Severe | 0.01 | 0.02 | 0.02 | 0.03 | 0.01 | 0.02 | 0.04 | 0.04 |
| Broad | Mild | 5.16 | 5.19 | 4.81 | 3.82 | 1.42 | 2.98 | 4.46 | 4.81 |
|  | Severe | 1.12 | 1.18 | 1.32 | 1.08 | 0.55 | 0.81 | 1.14 | 1.48 |
| Influenza |  |  |  |  |  |  |  |  |  |
| Narrow | Mild | 0.24 | 0.73 | 0.66 | 0.40 | 0.25 | 0.11 | 0.54 | 0.38 |
|  | Severe | 0.09 | 0.34 | 0.41 | 0.24 | 0.002 | 0.05 | 0.43 | 0.33 |
| Broad | Mild | 1.38 | 2.16 | 2.06 | 2.05 | 0.91 | 1.58 | 3.62 | 3.51 |
|  | Severe | 0.37 | 0.64 | 0.73 | 0.49 | 0.11 | 0.24 | 0.70 | 0.67 |
| COVID-19 |  |  |  |  |  |  |  |  |  |
| Narrow | Mild |  |  |  | 2.95 | 91.86 | 239.24 | 16.18 | 3.27 |
|  | Severe |  |  |  | 0.70 | 2.75 | 2.66 | 0.88 | 0.59 |
| Broad | Mild |  |  |  | 8.66 | 99.3 | 248.3 | 26.72 | 15.52 |
|  | Severe |  |  |  | 0.70 | 2.75 | 2.66 | 0.88 | 0.59 |
| Children and Young People |  |  |  |  |  |  |  |  |  |
| Annual Cohort |  | 2016-17 | 2017-18 | 2018-19 | 2019-20 | 2020-21 | 2021-22 | 2022-23 | 2023-24 |
| RSV |  |  |  |  |  |  |  |  |  |
| Narrow | Mild | 0.26 | 0.31 | 0.37 | 0.28 | 0.14 | 0.38 | 0.37 | 0.44 |
|  | Severe | 0.06 | 0.06 | 0.08 | 0.14 | 0.08 | 0.21 | 0.21 | 0.25 |
| Broad | Mild | 10.98 | 10.94 | 10.06 | 8.28 | 3.05 | 7.11 | 9.82 | 12.23 |
|  | Severe | 1.67 | 1.63 | 1.81 | 1.53 | 0.88 | 1.68 | 1.64 | 2.59 |
| *Influenza* |  |  |  |  |  |  |  |  |  |
| Narrow | Mild | 0.13 | 0.35 | 0.28 | 0.39 | 0.05 | 0.07 | 0.60 | 0.42 |
|  | Severe | 0.04 | 0.18 | 0.31 | 0.30 | 0.01 | 0.07 | 0.66 | 0.47 |
| Broad | Mild | 3.88 | 4.50 | 4.27 | 4.88 | 1.69 | 4.01 | 11.1 | 10.85 |
|  | Severe | 1.76 | 2.29 | 2.38 | 2.02 | 1.18 | 1.74 | 2.28 | 2.12 |
| *COVID-19* |  |  |  |  |  |  |  |  |  |
| Narrow | Mild |  |  |  | 0.73 | 65.35 | 306.21 | 3.27 | 0.89 |
|  | Severe |  |  |  | 0.07 | 0.31 | 0.94 | 0.24 | 0.24 |
| Broad | Mild |  |  |  | 2.98 | 69.93 | 314.57 | 12.83 | 13.24 |
|  | Severe |  |  |  | 0.07 | 0.31 | 0.94 | 0.25 | 0.25 |
| *Infants* |  |  |  |  |  |  |  |  |  |
| Annual Cohort |  | 2016-17 | 2017-18 | 2018-19 | 2019-20 | 2020-21 | 2021-22 | 2022-23 | 2023-24 |
| *RSV* |  |  |  |  |  |  |  |  |  |
| Narrow | Mild | 43.39 | 48.08 | 62.31 | 57.16 | 28.71 | 73.23 | 81.59 | 90.32 |
|  | Severe | 24.74 | 25.58 | 30.27 | 26.97 | 13.7 | 31.32 | 32.56 | 37.14 |
| Broad | Mild | 95.57 | 99.94 | 113.73 | 98.38 | 54.6 | 117.24 | 127.23 | 139.95 |
|  | Severe | 31.65 | 32.70 | 37.84 | 32.72 | 17.12 | 37.20 | 37.54 | 43.47 |
| *Influenza* |  |  |  |  |  |  |  |  |  |
| Narrow | Mild | 0.16 | 0.38 | 0.66 | 0.71 | 0.06 | 0.14 | 1.80 | 1.25 |
|  | Severe | 0.18 | 0.77 | 1.53 | 1.33 | 0.06 | 0.36 | 3.34 | 2.31 |
| Broad | Mild | 16.56 | 17.70 | 18.56 | 14.11 | 8.12 | 13.33 | 20.91 | 22.44 |
|  | Severe | 39.92 | 45.05 | 49.41 | 40.32 | 26.64 | 38.91 | 40.78 | 44.46 |
| *COVID-19* |  |  |  |  |  |  |  |  |  |
| Narrow | Mild |  |  |  | 0.73 | 25.44 | 68.39 | 6.87 | 5.35 |
|  | Severe |  |  |  | 0.38 | 1.91 | 8.54 | 4.61 | 5.62 |
| Broad | Mild |  |  |  | 8.88 | 43.36 | 93.08 | 31.15 | 37.1 |
|  | Severe |  |  |  | 0.41 | 1.93 | 8.67 | 4.74 | 5.70 |

#### Older Adult Cohort

In older adults, when using the narrow mild phenotypes, the number of phenotyped cases was highest for influenza until 2020, after which COVID-19 cases dominated (Figure 2A). Around 3,000-4,500 cases of mild influenza were phenotyped per annual cohort pre-COVID-19, while over 100,000 cases of COVID-19 were phenotyped in 2020-21. There were much lower numbers of RSV phenotyped cases (100-1,000 per annual cohort), with a larger number in the most recent annual cohort (2023–24). For the narrow phenotype, overlap of virus classification was minimal for mild cases - at most 0.3% (supplementary Figure S6A).

**Figure 2.**
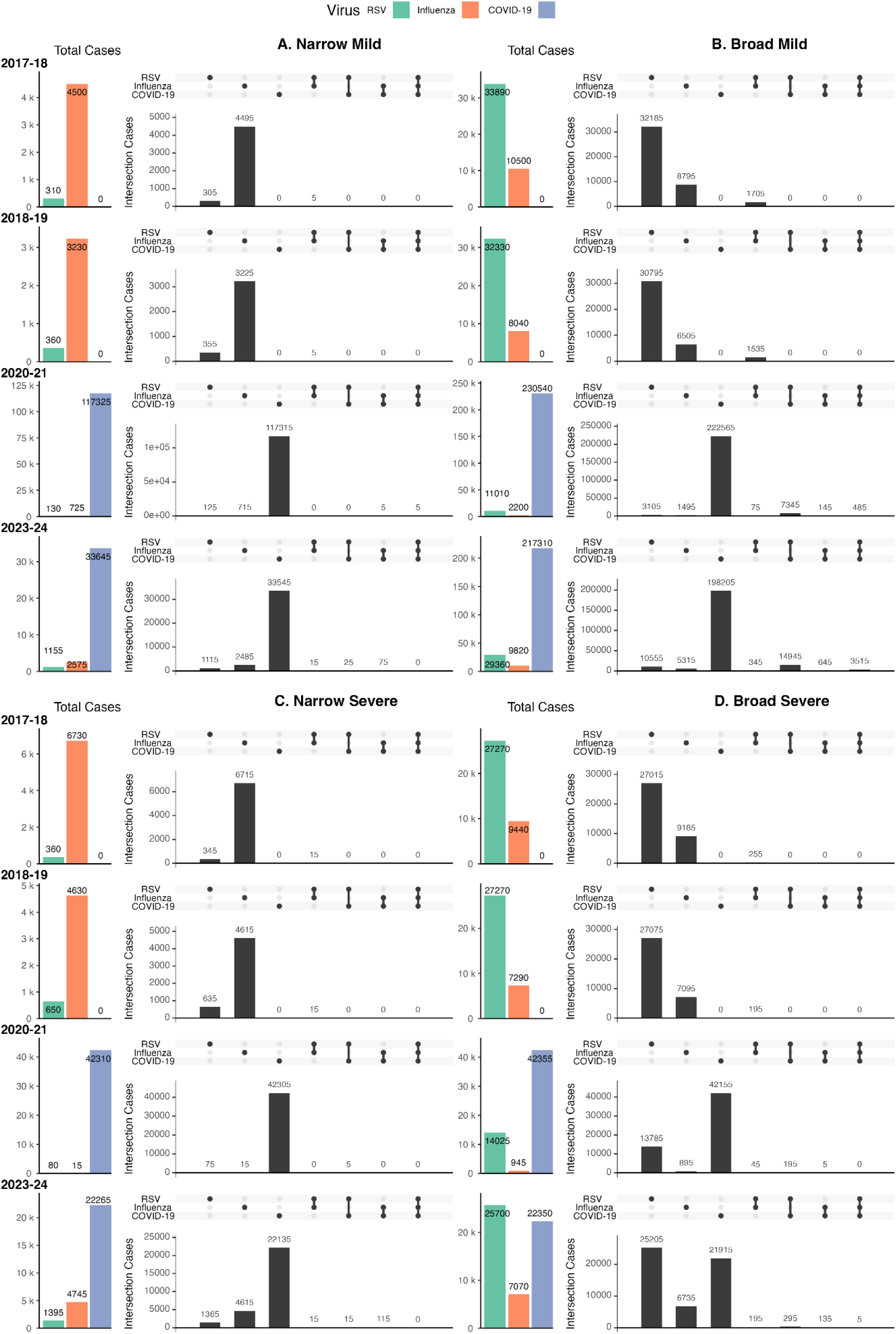
Classification of respiratory viruses in older adults. We. compared narrow phenotypes (A, C) and broad phenotypes (B, D), where cases included RSV (green), influenza (orange) or COVID-19 (purple). Mild cases (A, B) and severe cases (C, D) were classified independently.

We found much higher numbers of potential RSV cases when using the broad mild phenotypes (Figure 2B), ranging from 25-100 fold higher than the narrow phenotype. This meant that depending on phenotype chosen, the “dominant” winter virus in England was different. When compared to using the narrow phenotypes, the number of cases classified as influenza and COVID-19 by the broad phenotypes was slightly higher (∼2.5 fold higher for influenza and ∼2-6 fold higher for COVID-19). The level of overlap in case classification was also higher when using broad compared with narrow phenotypes (∼300 fold before 2020-21 and ∼150 fold subsequently). From 2020-21 onwards, we found many cases of overlap between RSV and COVID-19 - with over 50% of RSV phenotyped cases being classified simultaneously as COVID-19. Compared with the narrow phenotype, which had no more than 0.5% of phenotyped cases overlapping across all classified cases, use of the broad phenotype led to between ∼2-6% overlapping phenotyped cases (supplementary Figure S6B).

For severe cases in older adults classified using the narrow phenotypes, we found the same dominant viruses as with mild cases - influenza dominated until COVID-19 emerged and became the dominant virus (Figure 2C). Except for 2020-21, ∼5,000 severe cases were classified as influenza in each season - whereas there were between ∼20,000 (2023–24) and ∼40,000 (2020–21) severe cases classified as COVID-19 in OpenSAFELY.

As observed for mild RSV, severe RSV was detected at a lower rate - however, this increased over time (∼350 in 2017-18 versus ∼1400 in 2023-24). Overlap in the classification of phenotyped severe cases was rare when using the narrow phenotypes (<0.75%) (supplementary Figure S6C).

Use of broad severe phenotypes, compared to narrow phenotypes, led to similar numbers of phenotyped cases classified as COVID-19 (∼50-100 additional cases) and a moderate number of additional phenotyped cases classified as influenza (∼3,000 outside of 2020-21) (Figure 2D). However, use of the broad phenotype led to a much larger number of severe RSV phenotyped cases (between ∼20-175 fold increase). Notably, the key change between narrow and broad phenotypes for severe RSV was the addition of an ICD-10 diagnosis code for unspecified acute lower respiratory infection in the first or second position in HES, meaning that this code accounted for the additional phenotyped cases when using the broad severe RSV phenotype. Overlapping classification occurred in ∼0.5-1.5% of all cases when using the broad severe phenotypes (supplementary Figure S6D). In most annual cohorts we found ∼200-500 cases of RSV additionally classified as either influenza or COVID-19; there were often more cases of RSV additionally classified as COVID-19, compared to influenza (Figure 2D).

We found consistent results in all annual cohorts (2016–24) for both mild and severe phenotyped cases, with some variation over time (supplementary Figures S7-S8).

#### Adult and Children and Young People Cohorts

In both adults and children and young people, narrow phenotypes detected very low rates (<1 case per 1,000 person-years) of RSV and influenza (Table 1). Broad phenotypes increased the rate of phenotyped RSV cases over 100-fold in adults, and over 10-fold in children and young people and the rate of phenotyped influenza cases over 5-fold in adults and over 2-fold in children and young people. Mild phenotyped COVID-19 cases, irrespective of phenotype, were much more common than RSV or influenza. Rates were initially higher in adults, but in 2021-22 were highest in children and young people - reaching over 300 per 1,000 person-years. Phenotyped severe COVID-19 cases were rarer, never going above 3 cases per 1,000 person-years in adults or 1 case per 1,000 person-years in children and young people.

For adults, the introduction of COVID-19 increased the level of overlap in case classification (supplementary Figures S9). Overlap was higher for broader phenotype definitions, particularly for mild cases compared to severe (supplementary Figures S10-S11). The impact of phenotype on overlap in case classification in children and young people was very similar (supplementary Figure S12). Broader definitions led to higher levels of case classification overlap in mild phenotyped cases when compared to severe (supplementary Figures S13-S14).

#### Infant Cohorts

When using the narrow mild phenotypes in infants, we found most phenotyped cases were RSV (Figure 3A); between ∼12,000-33,000 mild cases annually. In each annual cohort, we classified 200-500 cases as influenza, except for 2020-21, and ∼2,000-10,000 cases as COVID-19. RSV remained dominant even during 2020-21 when COVID-19 phenotyped cases were also very common. Across all annual cohorts, a small amount of overlap was present for narrow mild RSV phenotyped cases (supplementary Figure S15A), with cases additionally classified either as influenza ( ∼5-25) or COVID-19 (∼5-25) (Figure 3A).

**Figure 3.**
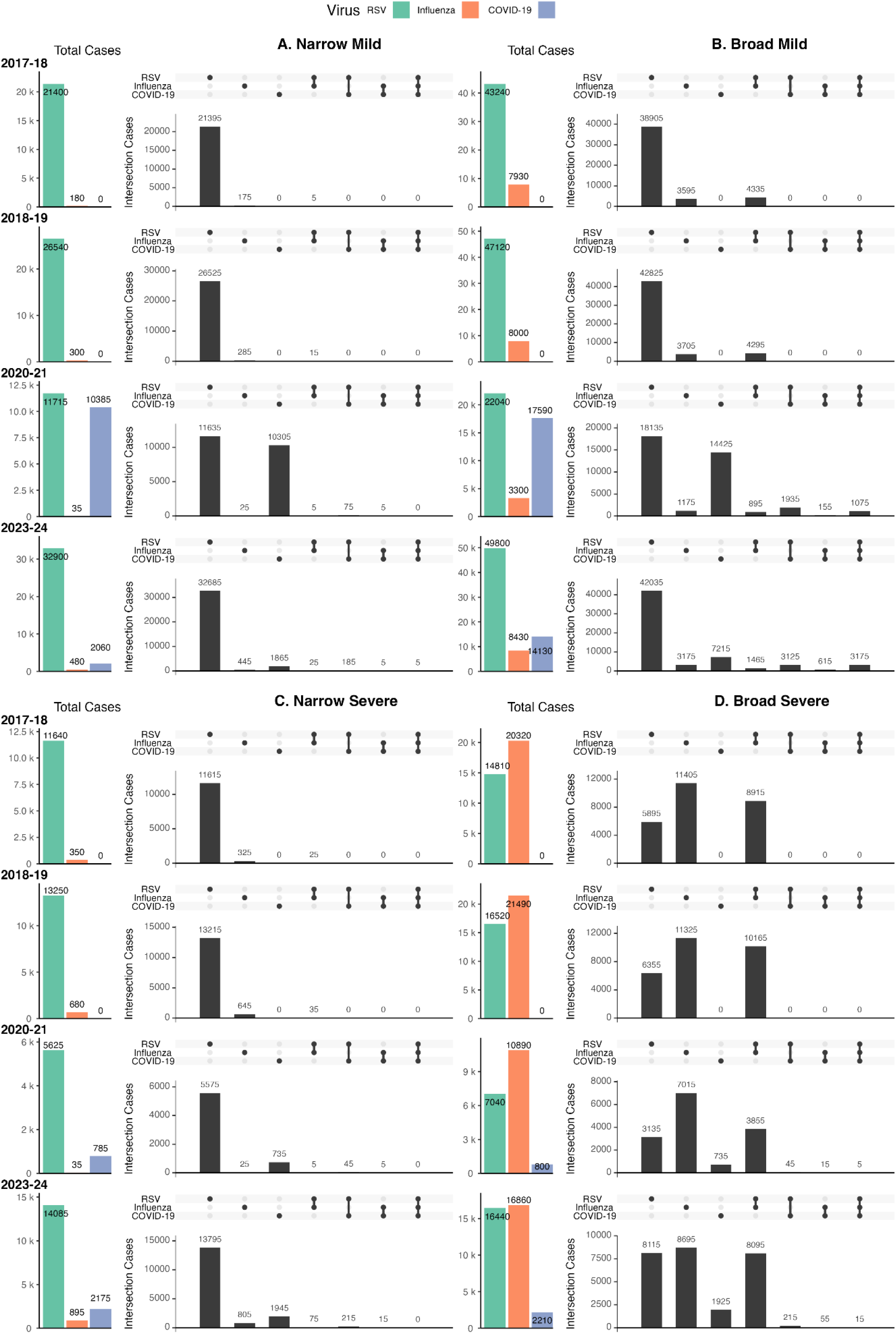
Classification of respiratory viruses in infants. We compared narrow phenotypes (A, C) and broad phenotypes (B, D), where cases included RSV (green), influenza (orange) or COVID-19 (purple). Mild cases (A, B) and severe cases (C, D) were classified independently.

The broad mild phenotype in infants resulted in much higher numbers of phenotyped cases (Figure 3B): RSV was ∼2 fold higher, influenza was ∼20-100 fold higher, and COVID-19 was ∼2-5 fold higher. The latter two corresponded to a large number which were classified simultaneously as another virus (often RSV): case classification overlap for influenza (pre-COVID-19: ∼4,500 cases; post-COVID-19: ∼1,000-1,500 cases) and COVID-19 (∼2,000-3,000 cases). Around 8% of cases were classified as more than one virus (supplementary Figure S15B), with a large number of mild cases being classified as all three viruses (between ∼1,000-3,000) (Figure 3B).

RSV was the dominant virus when using the narrow severe phenotypes, resulting in ∼5,500-15,000 phenotyped cases annually (Figure 3C). Influenza phenotyped cases increased between 2017-18 (350) and 2023-24 (895), with an exception in 2020-21. Severe COVID-19 phenotyped cases were more common than influenza (∼1,000 more cases of COVID-19 in 2020-21 and 2023-24 respectively) but still much less common than RSV. For most annual cohorts, the degree to which cases overlapped was minimal (<500 cases - with cases often classified as RSV). The percentage of phenotyped cases which had overlapping classification increased from <0.25% prior to 2019-20 to between 0.7-2.5% subsequently (supplementary Figure S15C), many of which included COVID-19 overlapping with RSV (Figure 3C).

Using broad severe phenotypes meant influenza phenotyped cases became dominant (Figure 3D). The number of severe COVID-19 phenotyped cases changed very little between narrow and broad phenotypes. The change in dominance was also reflected in a change in case overlap (>100-fold increase), with most phenotyped cases in each annual cohort being classified as both RSV and influenza. This change, which was not seen in older adults (Figure 2), indicates a difference in how clinicians coded episodes, with pathogen-associated codes more likely in infant records. The change in overlap in severe cases was also notable, <3% versus almost 30% of cases for narrow versus broad phenotypes (supplementary Figure S15D).

The differences between narrow and broad phenotyped cases for mild and severe disease was consistent across annual cohorts (supplementary Figures S16-S17). Similarly, the overlap in case classification was consistent in maternally-linked infants (supplementary Figure S18). Mild cases and severe cases were also impacted in the same manner as with the full infant cohort - suggesting that the clinical coding patterns in the records of this subgroup were not different (supplementary Figures S19-S20).

#### Phenotype Design Sensitivity Analyses

When we explored the relative contribution of common codes to case ascertainment, we found that in mild phenotypes, the most common broad symptom code (‘cough (finding)’) contributed more cases in older adults (42.1%) compared to infants (11.7%) for RSV. For influenza, this code contributed a similar proportion of cases in infants (20.4%) and older adults (23.5%), while for COVID-19 this code contributed more cases in infants (28%) compared to older adults (6.2%). For severe influenza, broad codes were far bigger drivers in infants (J069: 33.2%; J219: 43.9%) compared to older adults (J069: 13.6%; J219: 0.99%). The code B972 contributed <1% of cases for severe COVID-19. When we explored the relative case contribution of antivirals for broad mild RSV, we found <1% of cases attributable to these prescriptions. See supplementary Table S13 for more detail.

#### Internal Validation of Mild Phenotypes

Across age cohorts, the majority of severe cases classified using narrow phenotypes were not preceded by a mild case, thus reflecting either direct presentations to secondary care or those preceded by presentations that did not meet our mild definitions (supplementary Figure S21). Among the severe reference cases with a mild presentation in the 14 days preceding hospitalisation, a subset of mild cases could be attributed to RSV, influenza, or COVID-19, with the proportion varying by phenotype and age cohort (supplementary section S3.6.2). Among the subset of mild cases attributable to one or more of the pathogens of interest, agreement with the severe reference cases was high, particularly for narrow phenotypes. Median agreement across annual cohorts, stratified by age cohort and phenotype, was generally highest for COVID-19 (87.3-99.7%). For influenza, median agreement in non-infant cohorts was 97.1-99.2% (narrow) and 93.6-94.6% (broad), while median agreement in the infant cohort was 72.3% (narrow) and 73.4% (broad). For RSV, median agreement in the infant cohort was 99.6% (narrow) and 98.5% (broad), while in non-infant cohorts median agreement was 84.7-96.2% (narrow) and 76.9-89.7% (broad).

#### Reinfections

Reinfections occurred at different levels depending on the virus and case severity, but rarely occurred in more than 15% of individuals (supplementary Figure S34). Phenotype sensitivity impacted the proportion of individuals being reinfected, where broader phenotypes had more reinfections.

Due to low counts classified by the narrow phenotypes, it was difficult to explore the proportion of reinfections that occurred within 28 days for RSV and influenza. In infants, where RSV reinfections were more common, the proportion of reinfections occurring within 28 days was consistently ∼25%. For COVID-19, narrow phenotypes consistently highlighted higher proportions of reinfections within 28 days when compared to broad phenotypes (supplementary Figure S35). When we explored extending the COVID-19 episode length to 28 days it had minimal impact (<5% change) on case counts and reinfections, apart from severe reinfections which were decreased by 40-45% (supplementary Table S13).

#### Case Classification Types

Overlapping case classifications were more common when using broad phenotypes, across all age cohorts (e.g. older adults in 2023-24 narrow versus broad: 0.3% versus 6.2% (mild) and 0.5% versus 1.1% (severe)). However, overlapping classifications did occur when using narrow phenotypes. This was more common in infants than other age cohorts (e.g. in 2023-24: 0.6% (mild) and 1.8% (severe); supplementary Figures S6, S9, S12, S15, and S18.

The percentage of overall respiratory virus cases that were also RSV, influenza, or COVID-19 phenotyped cases increased over time. Prior to 2020-21, for mild cases in older adults, adults, and children and young people, between ∼10-35% of overall respiratory virus cases were classified as either RSV, influenza, or COVID-19; this rose to over 70% of cases in 2020-21 and 2021-22, settling between ∼30-60% subsequently (supplementary Figures S36A, S37A, and S38A). For severe cases, in these age cohorts, the percentage of overall respiratory virus cases classified as RSV, influenza, or COVID-19 rose slightly over time averaging ∼50% (supplementary Figures S36B, S37B, and S38B). For infants and maternally-linked infants the percentage of both mild and severe overall cases being classified as RSV, influenza, or COVID-19 was more stable at ∼60-70% (mild) and ∼85-90% (severe) (supplementary Figures S39-S40).

### Comparison to Surveillance Data

Surveillance sources varied slightly in the number of cases reported, and years with data available (supplementary Figure S41).

#### Descriptive Comparisons

The timing of peaks in surveillance data was reflected in cases classified by both narrow and broad phenotypes (Figure 4 and supplementary Figure S42). The relative scale of reported cases varied across viruses and phenotypes (supplementary Figure S43); which is expected as OpenSAFELY covers 42% of the population.

**Figure 4.**
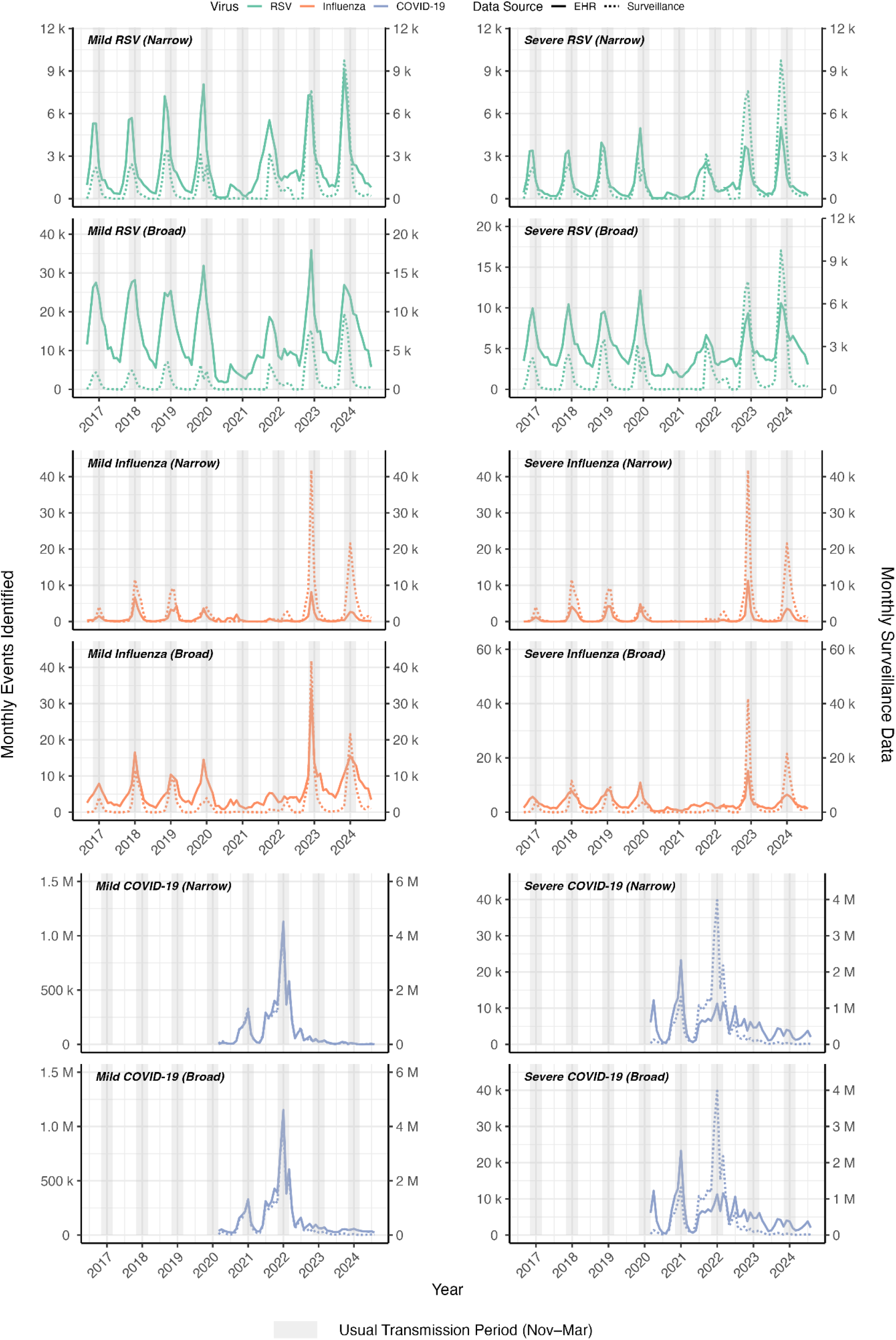
Comparison of phenotyped cases in OpenSAFELY (left axis) and surveillance data (right axis). WHO RespiMart surveillance data for RSV and influenza were for England and for COVID-19 were for the UK. Results are shown separately for mild (left panels) and severe cases (right panels), with narrow cases appearing in upper panels and broad cases in lower panels for each virus - where RSV is green, influenza is orange, and COVID-19 is purple. Surveillance data are indicated by dotted lines, with phenotyped cases indicated by solid lines.

For RSV, phenotyped cases exceeded those recorded in surveillance data (often over 2 (narrow) to 6 (broad) fold) until 2022-23 where surveillance recordings increased. Initially, similar numbers of cases were classified by narrow influenza phenotypes and recorded in surveillance data. However, in more recent years surveillance recordings of influenza cases increased; by 2022 roughly half of cases reported in surveillance data were phenotyped in EHR data - in line with the population proportion in OpenSAFELY. The broad phenotype classified ∼2-15-fold higher influenza cases than recorded in surveillance data, which was reflected in wider epidemic curves - as opposed to sharper peaks.

Between 2020-22, surveillance data reported very few cases of RSV and influenza compared to phenotyped cases from EHRs - irrespective of severity and phenotype (Figure 4). This corresponds to a period in which respiratory virus presentations were likely to be tested for COVID-19 but may not have been coded as such by clinicians. Mild cases of COVID-19 reported via OpenSAFELY aligned closely, in the timing of peaks, with surveillance data; although 4 times more cases were recorded in surveillance data (supplementary Figure S43).

Some peaks in severe COVID-19 phenotyped cases were not shown in surveillance data, likely due to an ICD-10 code used for COVID-19 (U07.2) which was informed by clinical diagnosis without test confirmation.

#### Quantitative Comparisons

We found high temporal correlation between surveillance data and phenotyped cases from OpenSAFELY (Figure 5). The degree of correlation varied both overall (Table 2) and by annual cohort (supplementary Figure S44 and Table S14). The annual cohorts affected by lockdowns had much lower correlation for RSV and influenza, because there were no annual epidemics during these years. In the same period, testing for COVID-19 was common and reflected milder cases. As a result, correlation between surveillance data for severe COVID-19 was consistently lower than for mild COVID-19. When freely available testing ceased, correlations for mild and severe COVID-19 became more similar. Correlations varied more across annual cohorts than by the phenotype employed, however, for influenza in particular, phenotyped cases from broad phenotypes were often less correlated with surveillance data than narrow phenotypes.

**Figure 5.**
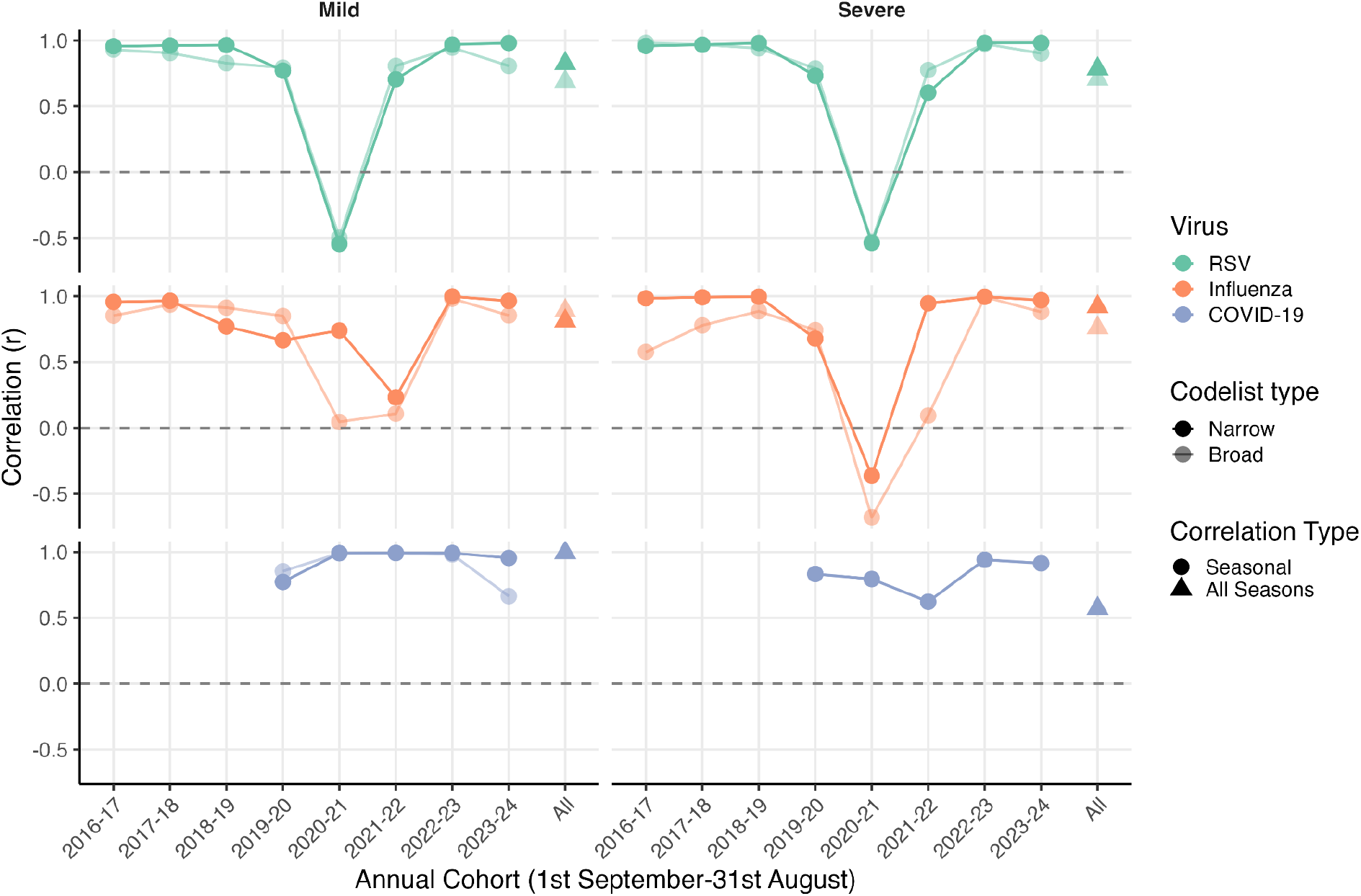
Pearson’s correlation coefficient for each virus, severity, and phenotype combination. . Tests were performed individually for each annual cohort. The colour of the line indicates the virus (RSV in green, influenza in orange, and COVID-19 in purple); the darkness of the shade indicates phenotype, where lighter shades are broad phenotypes; the shape of the point indicates whether the correlation is on a seasonal level or for all annual cohorts (seasonal: filled circle; all annual cohorts: filled triangle).

**Table 2.** Pearson’s correlation coefficient with corresponding 95% CI for each case and phenotype combination. The results are for all annual cohorts simultaneously, for each virus/severity/phenotype respectively.

| Classification | Correlation Coefficient (95% CI) |  |
| --- | --- | --- |
|  | Narrow | Broad |
| Mild RSV | 0.82 (0.75, 0.88) | 0.69 (0.57, 0.78) |
| Severe RSV | 0.78 (0.69, 0.85) | 0.70 (0.59, 0.79) |
| Mild Influenza | 0.81 (0.73, 0.87) | 0.89 (0.84, 0.92) |
| Severe Influenza | 0.92 (0.88, 0.95) | 0.76 (0.66, 0.83) |
| Mild COVID-19 | 1.00 (0.99, 1.00) | 0.99 (0.99, 1.00) |
| Severe COVID-19 | 0.57 (0.36, 0.73) | 0.57 (0.36, 0.73) |

## Discussion

We developed phenotypes to classify and differentiate cases of RSV, influenza, and COVID-19 from routinely-collected health data in England. We performed an internal validation using a subset of phenotyped cases and compared phenotyped cases to national surveillance data.

Prior work has validated phenotypes for acute respiratory infections for surveillance and early warning systems (26), and here we focus on differentiating the key viruses which contribute towards acute respiratory infections. Our study has detailed methods for classifying cases for three key respiratory viruses in healthcare data across primary and secondary care, without the use of testing information. These methods could be valuable for quantifying the burden of these viruses experienced in primary care, and may deepen understanding of how these common respiratory viruses impact patients and GP practices. This is crucial for understanding the healthcare burden and patterns of primary and secondary care utilisation of these major respiratory viruses, for which vaccines are in use.

In older adults, the narrow phenotypes classified similar rates of mild and severe cases for both RSV and influenza, and rates of severe cases at times exceeded mild cases. This represents an inversion of the usual clinical pyramid (wherein a subset of mild cases progress to severe disease) and highlights the suboptimal sensitivity of pathogen-specific coding for respiratory virus presentations in primary care. This was likely impacted by extremely limited testing in primary care. Conversely, mild COVID-19 cases far exceeded severe cases in the 2020-21, 2021-22, and 2022-23 annual cohorts because testing was widely available outside of hospital settings during this time. Rates of severe phenotyped cases, particularly for RSV and influenza, were much lower when using narrow phenotypes compared to broad, this may be due to pathogen-specific ICD-10 coding which may underestimate case burden (39), whereas pathogen-associated ICD-10 codes may lead to case misclassification. While overlap in severe phenotyped cases was rare, even when using the broad phenotypes, this does not mean that misclassification was not present. In mild cases, broad phenotypes led to increased overlap in classification of COVID-19 and RSV phenotyped cases. However, surveillance data suggests that these were COVID-19 cases being misclassified as RSV by our broad phenotypes. This highlights how difficult it is to distinguish viruses leading to respiratory infection in EHR data and how pandemic shocks can disrupt these surveillance efforts.

Comparisons in older adults of the narrow phenotypes and the broader phenotypes changed which virus appeared dominant (influenza versus RSV) prior to 2020-21. While there is existing evidence that RSV burden may be underestimated (40), the large discrepancy between RSV and influenza burden shown by broad phenotypes suggests poor influenza ascertainment (4). In mild cases this was largely driven by symptom-based codes in the broad phenotypes.

However, for influenza these had a smaller impact on case ascertainment due to the requirement of fever codes being present - in line with the WHO ILI definition (25). In severe cases this was due to the use of ICD-10 code J22 (‘unspecified acute lower respiratory infection’) which was included only for the RSV phenotype as informed by previous research (41).

In infants, we found low numbers of influenza cases using narrow phenotypes, suggesting these cases were difficult to ascertain in primary care; however, the broad phenotype had higher overlap, suggesting broader case definitions made influenza hard to distinguish from other viruses. Broad phenotypes also resulted in more cases for RSV and COVID-19; however, as with influenza, this often corresponded to greater overlap in classification compared with the narrow phenotypes. This is likely due to bronchiolitis codes, which, while a very common presentation for RSV (17,18), could also be listed in cases with influenza or COVID-19 codes leading to simultaneous classification with RSV. Overlap in case classification was seen, particularly in infants, in both broad and narrow phenotyped cases, indicating difficulty distinguishing syndromic presentation or potential co-infection.

Clinical coding approaches appeared to differ by age cohort. In infants, there was more overlap in case classification for broad severe phenotyped cases compared to non-infant cohorts; suggesting that pathogen-associated codes were more common. This was further highlighted by the larger proportions of cases attributable to common individual pathogen-associated ICD-10 codes in infants versus older adults. Our results align with preliminary findings across European healthcare settings, including England, of age-dependent coding biases, including greater divergence between respiratory tract infection (RTI)- and RSV-coded hospitalisation diagnosis groups recorded in younger versus older populations (42).

We used severe cases with relevant ICD-10 codes (classified by our severe narrow phenotypes) as a reference for internal validation of our mild phenotypes. A high proportion of mild presentations that preceded severe cases could not be attributed to RSV, influenza, or COVID-19, reflecting the limited availability of testing in primary care settings and consequent constraints in pathogen-specific case detection. However, for the subset of mild cases attributable to one or more of our pathogens of interest, we observed high consistency with the severe reference case. This reflects positively on the specificity for our mild phenotypes, although our internal validation should be interpreted with caution given that the subset of mild cases resulting in hospitalisations may not be generalisable to cases detected only in primary care. Further evaluation of the diagnostic validity of our phenotypes with a ‘gold standard’ reference, e.g. PCR-confirmed infections, would be a valuable extension if the necessary data linkages can be established.

When evaluating phenotypes we found that reinfections were relatively rare across the population. A high proportion of COVID-19 reinfections occurred within 28 days, suggesting that the episode length definition (14 days) might have been too short particularly for severe cases. Future work concerned with distinguishing separate COVID-19 hospitalisations should consider using a 28-day episode length. We also found that a high proportion of overall respiratory virus cases were not classified as RSV, influenza, or COVID-19. This was indicative of how common undifferentiated respiratory codes were, which is in line with both historic and current findings on RSV, influenza, and COVID-19 prevalence. A previous study found that ∼30% of NHS Direct respiratory calls could be attributed to RSV or influenza, with variation by age (43). Additionally, recent virological data shows that RSV, influenza, and COVID-19 accounted for less than half of respiratory virus samples, with proportions differing by age (44).

We found the timing and size of epidemics changed over time in both surveillance data and in EHRs. In addition to differences in epidemic sizes during this time, this change may also be in part due to changes in clinical coding behaviours, surveillance data reporting, or testing practices. Testing, for example, was limited to hospital settings for RSV and influenza until recently. However, for COVID-19, NHS testing outside of hospital was common between 2020-2022. Therefore, narrow phenotypes classified fewer mild cases of RSV and influenza but for COVID-19 may have ascertained cases which would not usually present to health services - e.g. very mild or asymptomatic cases. Ascertainment by RSV and influenza narrow phenotypes, both mild and severe, increased in more recent annual cohorts, while broad phenotypes did not exhibit consistent changes. This may be due to increased panel testing implemented in part as a response to COVID-19, resulting in increased usage of pathogen-specific codes (27).

The phenotypes outlined here could be expanded to include other key respiratory viruses such as adenovirus or rhinovirus, and future development and strengthening of the findings could include securing linkage of testing data, including vaccine history of patients, and the potential for analysis of free text in the EHR, which is not available at this time. Downstream analyses using these phenotypes also have the potential to investigate patient-level characteristics associated with clinical coding specificity, including understanding of the major health inequalities in burden of these viruses in England.

### Strengths and Limitations

We leveraged very large scale EHR data through OpenSAFELY with detailed coding on clinical events and medications prescribed within primary care, plus linked secondary care including emergency attendances, which gave high granularity for designing phenotypes. We did not have access to free text fields, testing data, dispensation of prescriptions, or medications administered within secondary care. Free text constitutes up to 80% of data within health records (45), such that case ascertainment may underestimate true burden.

While designing EHR phenotypes we identified difficulties associated with the absence of linked testing data alongside uncertainty surrounding information flow between primary and secondary care. Notably, we observed a high proportion of undifferentiated respiratory viruses across age groups in both primary and secondary care settings, highlighting the potential for more systematic pathogen-specific coding in settings where testing data are available. By enforcing temporal progression of case severity from mild to severe we also ensured that those who presented only to secondary care were not classified in primary care due to communication across systems. Another difficulty was distinguishing symptom types within a limited number of codelists (e.g. one codelist for possible RSV symptoms); in order to enforce the presence of two distinct symptoms in broader phenotypes, symptom-based codelists would need to be used. However, this adds further layers of computational complexity.

## Conclusion

EHR data phenotypes can be used to classify mild and severe cases of three key respiratory viruses. This may provide a useful picture of respiratory virus burden within primary and secondary care, particularly for vaccine-preventable respiratory diseases. For younger patient populations, broader case definitions had higher chances of misclassification than narrower phenotypes. The relative scale and timing of peaks in EHR-recorded cases was generally well aligned with surveillance data; however, diagnostic validity was only explored against an internal reference standard and has not yet been established against an independent gold standard. We highlight the difficulty of distinguishing characteristics of viral epidemiology and evolving coding behaviour, both across age and over time. The phenotypes outlined here offer a plausible adjunct to existing surveillance efforts and may support further epidemiological evaluation of respiratory viruses.

## Data Availability

All data were linked, stored and analysed securely using the OpenSAFELY platform, https://www.opensafely.org/, as part of the NHS England OpenSAFELY COVID-19 service. Data include pseudonymised data such as coded diagnoses, medications and physiological parameters. No free text data are included. No GP data from patients who have registered a Type-1 Opt out with their GP surgery were included in this study. Detailed pseudonymised patient data is potentially re-identifiable and therefore not shared. Primary care records managed by the GP software provider, TPP, were linked to hospital episode statistics (HES) and ONS death data through OpenSAFELY.
NHS England is the data controller of the NHS England OpenSAFELY COVID-19 Service; TPP is the data processor; all study authors using OpenSAFELY have the approval of NHS England (83). This implementation of OpenSAFELY is hosted within the TPP environment which is accredited to the ISO 27001 information security standard and is NHS IG Toolkit compliant (84).
Patient data has been pseudonymised for analysis and linkage using industry standard cryptographic hashing techniques; all pseudonymised datasets transmitted for linkage onto OpenSAFELY are encrypted; access to the NHS England OpenSAFELY COVID-19 service is via a virtual private network (VPN) connection; the researchers hold contracts with NHS England and only access the platform to initiate database queries and statistical models; all database activity is logged; only aggregate statistical outputs leave the platform environment following best practice for anonymisation of results such as statistical disclosure control for low cell counts (85).
The service adheres to the obligations of the UK General Data Protection Regulation (UK GDPR) and the Data Protection Act 2018. The service previously operated under notices initially issued in February 2020 by the the Secretary of State under Regulation 3(4) of the Health Service (Control of Patient Information) Regulations 2002 (COPI Regulations), which required organisations to process confidential patient information for COVID-19 purposes; this set aside the requirement for patient consent (86). As of 1 July 2023, the Secretary of State has requested that NHS England continue to operate the Service under the COVID-19 Directions 2020 (87). In some cases of data sharing, the common law duty of confidence is met using, for example, patient consent or support from the Health Research Authority Confidentiality Advisory Group (88).
Taken together, these provide the legal bases to link patient datasets using the service. GP practices, which provide access to the primary care data, are required to share relevant health information to support the public health response to the pandemic, and have been informed of how the service operates.
All code is shared openly for review and re-use under MIT open license in the following repository: https://github.com/opensafely/disparities-comparison.

## Declarations

### Ethics Approval and Consent to Participate

This study was approved, in accordance with the declaration of Helsinki, by the Human Research Authority (HRS) (IRAS ID: 340706) and London School of Hygiene & Tropical Medicine (LSHTM) Ethics (29879). NHS England is the data controller of the NHS England OpenSAFELY COVID-19 Service; TPP is the data processor; all study authors using OpenSAFELY have the approval of NHS England (1). This implementation of OpenSAFELY is hosted within the TPP environment which is accredited to the ISO 27001 information security standard and is NHS IG Toolkit compliant (2).

Patient data has been pseudonymised for analysis and linkage using industry standard cryptographic hashing techniques; all pseudonymised datasets transmitted for linkage onto OpenSAFELY are encrypted; access to the NHS England OpenSAFELY COVID-19 service is via a virtual private network (VPN) connection; the researchers hold contracts with NHS England and only access the platform to initiate database queries and statistical models; all database activity is logged; only aggregate statistical outputs leave the platform environment following best practice for anonymisation of results such as statistical disclosure control for low cell counts (3).

The service adheres to the obligations of the UK General Data Protection Regulation (UK GDPR) and the Data Protection Act 2018. The service previously operated under notices initially issued in February 2020 by the Secretary of State under Regulation 3(4) of the Health Service (Control of Patient Information) Regulations 2002 (COPI Regulations), which required organisations to process confidential patient information for COVID-19 purposes; this set aside the requirement for patient consent (4). As of 1 July 2023, the Secretary of State has requested that NHS England continue to operate the Service under the COVID-19 Directions 2020 (5). In some cases of data sharing, the common law duty of confidence is met using, for example, patient consent or support from the Health Research Authority Confidentiality Advisory Group (6).

Taken together, these provide the legal bases to link patient datasets using the service. GP practices, which provide access to the primary care data, are required to share relevant health information to support the public health response to the pandemic, and have been informed of how the service operates.

1. The NHS England OpenSAFELY COVID-19 service - privacy notice. NHS Digital (Now NHS England). https://digital.nhs.uk/coronavirus/coronavirus-covid-19-response-information-governance-hub/the-nhs-england-opensafely-covid-19-service-privacy-notice (accessed 4 July 2023). ↩
2. Data Security and Protection Toolkit - NHS Digital. NHS Digital (Now NHS England). (accessed 4 July 2023) [archived here: https://web.archive.org/web/20250405100349/ https://digital.nhs.uk/services/data-security-and-protection-toolkit/data-security-and-protection-toolkit]. ↩
3. ISB1523: Anonymisation Standard for Publishing Health and Social Care Data. NHS Digital (Now NHS England). (accessed 4 July 2023) [archived here: https://web.archive.org/web/20250430135246/standards.nhs.uk/published-standards/anonymisation-standard-for-publishing-health-and-social-care-data]. ↩
4. Coronavirus (COVID-19): notice under regulation 3(4) of the Health Service (Control of Patient Information) Regulations 2002 – general. 2022. https://www.gov.uk/government/publications/coronavirus-covid-19-notification-of-data-controllers-to-share-information/coronavirus-covid-19-notice-under-regulation-34-of-the-health-service-control-of-patient-information-regulations-2002-general--2 (accessed 5 July 2023). ↩
5. Secretary of State for Health and Social Care - UK Government. COVID-19 Public Health Directions 2020: notification to NHS Digital. (accessed 4 July 2023) [archived here: https://web.archive.org/web/20250414105355/ https://digital.nhs.uk/about-nhs-digital/corporate-information-and-documents/directions-and-data-provision-notices/data-provision-notices-dpns/opensafely-covid-19-service-data-provision-notice]. ↩
6. Confidentiality Advisory Group. Health Research Authority. [archived here: https://web.archive.org/web/20250408154826/ https://www.hra.nhs.uk/about-us/committees-and-services/confidentiality-advisory-group/]. ↩

## Consent for Publication

Not applicable.

## Availability of Data and Materials

All data were linked, stored and analysed securely using the OpenSAFELY platform, https://www.opensafely.org/, as part of the NHS England OpenSAFELY COVID-19 service. Data include pseudonymised data such as coded diagnoses, medications and physiological parameters. No free text data are included. No GP data from patients who have registered a Type-1 Opt out with their GP surgery were included in this study. Detailed pseudonymised patient data is potentially re-identifiable and therefore not shared. Primary care records managed by the GP software provider, TPP, were linked to hospital episode statistics (HES) and ONS death data through OpenSAFELY. All code is shared openly for review and re-use under MIT open license in the following repository: https://github.com/opensafely/disparities-comparison.

## Author Contributions

EP, RME, EPKP, WH, CWG, and JKQ contributed to the conception and design of the work; BG, SB, AM, DE, WH contributed to data acquisition; EP, RME, EPKP, WH, DE, REC, SBMM, FY, MN, YA contributed to the analysis and/or interpretation of the data; BG, SB, AM, DE, RE, WH contributed to the creation of the software used within the work; and EP, RME, EPKP, WH, REC, CWG, JKQ, SBMM, FY, MN, YA contributed to the drafting and revision of the work.

## Acknowledgements

We would like to thank the PPIE advisory group for their insightful contributions to this research. We are grateful to the named contributors, Yvonne Alder, Sarah Barley-McMullen, Janice Elliott, Michael Natt, Farheen Yameen, as well as to those members who have chosen not to be named, whose perspectives were integral to shaping this study. We would like to acknowledge the contribution of Sophie Graham and Francis Collett-White for providing support on the design of COVID-19 primary care codelists; and the contributions of Michael Marks, Luisa Pettigrew, and William Elson for reviewing respiratory phenotypes. We would also like to acknowledge the continual support from the tech-team at the Bennett Institute for Applied Data Science, as well as fellow researchers using OpenSAFELY for their insight and advice. We are very grateful for all the support received from the TPP Technical Operations team throughout this work, and for generous assistance from the information governance and database teams at NHS England and the NHS England Transformation Directorate.

