## Supplementary Material for "Classifying and Differentiating Clinical Coding of Respiratory Syncytial Virus, Influenza, and COVID-19 in OpenSAFELY Between 2016 and 2024"

#### S1. Final Phenotypes

| *RSV* | |
|---|---|
| *Mild (Primary Care, Emergency Attendances)* | *Severe (Secondary Care)* |
| Narrow | |
| ● At least one RSV diagnosis code, or bronchiolitis diagnosis code, in SNOMED CT (1), OR,<br>● **In infant population**, bronchiolitis diagnosis code (2), in SNOMED CT, in first or second position in ECDS (A&E) - availability: partial 2018-2020; full 2020 onwards (3) | At least one ICD-10 diagnosis code for RSV or bronchiolitis in the first or second position in the HES APC (4) |
| Broad | |
| *Inclusion Criteria*<br><br>● Meets criteria for 'Narrow' phenotype, OR,<br>● At least two of the following respiratory symptoms recorded in SNOMED CT within a two-week period (two on the same day is acceptable): rhinitis, cough, fever, breathing abnormalities, and viral infections (5) OR,<br>● **In infant population**, at least one bronchiolitis (2) or viral wheeze (6) diagnosis code, in SNOMED CT, in first or second position in ECDS (A&E) - availability: partial 2018-2020; full 2020 onwards (3)<br><br>*Exclusion Criteria*<br><br>Diagnosis code in SNOMED CT for Influenza, pneumonia, human metapneumovirus, rhinovirus, adenovirus, parainfluenza, enterovirus, mycoplasma pneumonia, bronchitis, sinusitis, tonsillitis, laryngitis, pertussis, or coronavirus, including COVID-19, within the one-month period (±) of respiratory symptoms (7) | *Inclusion Criteria*<br><br>● Meets criteria for 'Narrow' phenotype, OR,<br>● ICD-10 diagnosis code for unspecified acute lower respiratory infection (8) in first or second position in the HES APC<br><br>*Exclusion Criteria*<br><br>ICD-10 diagnosis code in first or second position in HES APC for influenza, pneumonia, human metapneumovirus, rhinovirus, adenovirus, parainfluenza, enterovirus, mycoplasma pneumonia, bronchitis, sinusitis, tonsillitis, laryngitis, pertussis, or coronavirus, including COVID-19, within the one-month period (±) of respiratory symptoms (9) |

| *Influenza* | |
|---|---|
| *Mild (Primary Care)* | *Severe (Secondary Care)* |
| Narrow | |
| At least one SNOMED CT code for Influenza (10) | At least one ICD-10 diagnosis code for influenza in the first or second position in the HES APC (11) |
| Broad | |
| *Inclusion Criteria*<br><br>● Meets criteria for 'Narrow' phenotype, OR,<br>● At least one SNOMED CT code for influenza, suspected influenza, influenza-like-illness, test run for influenza (12), OR,<br>● Record of an acute respiratory illness (13) with a measured temperature of ≥ 38 °C (14) within the same episode (adapted from definition of influenza-like-illness according to WHO (15))<br>● At least one prescription for antivirals (oseltamivir or zanamivir) in Pseudo BNF relevant for the treatment of infectious respiratory diseases (16)<br><br>*Exclusion criteria*<br><br>Diagnosis code in SNOMED CT for RSV, pneumonia, human metapneumovirus, rhinovirus, adenovirus, parainfluenza, enterovirus, mycoplasma pneumonia, tonsillitis, pertussis, or coronavirus, including COVID-19, within the one-month period (±) of respiratory symptoms or receipt of a relevant prescription as noted in the inclusion criteria above (17) | *Inclusion Criteria*<br><br>● Meets criteria for 'Broad' phenotype, OR,<br>● At least one ICD-10 diagnosis code for acute respiratory illness in the first of second position in the HES APC (18)<br><br>*Exclusion criteria*<br><br>ICD-10 diagnosis code in first or second position in HES APC for RSV, pneumonia, human metapneumovirus, rhinovirus, adenovirus, parainfluenza, enterovirus, mycoplasma pneumonia, tonsillitis, pertussis, or coronavirus, including COVID-19, within the one-month period (±) of respiratory symptoms (19) |

<table>
<tr><th colspan="2">COVID-19</th></tr>
<tr><th>Mild (Primary Care)</th><th>Severe (Secondary Care)</th></tr>
<tr><td colspan="2">Narrow</td></tr>
<tr><td>At least one SNOMED CT code for COVID-19 (20)</td><td>At least one ICD-10 diagnosis code for COVID-19 in the first or second position in the HES APC (21)</td></tr>
<tr><td colspan="2">Broad</td></tr>
<tr><td>Inclusion criteria<ul><li>Meets criteria for ‘Narrow’ phenotype, OR,</li><li>At least one SNOMED CT code for COVID-19, coronavirus (without specific strain), suspected COVID-19, tests performed and referrals to support, OR, at least two of the following symptoms of COVID-19 recorded in SNOMED CT within a two-week period (two on the same day is acceptable): fever, cough, change to sense of smell or taste, shortness of breath, fatigue, body aches/muscle aches, headache, sore throat, rhinitis, loss of appetite, diarrhoea, nausea and vomiting (22), OR,</li><li>At least one prescription for antivirals (nirmatrelvir plus ritonavir (Paxlovid) and molnupiravir (Lagevrio)) in Pseudo BNF (23)</li></ul>Exclusion criteria<br><br>Diagnosis code in SNOMED CT for RSV, Influenza, pneumonia, human metapneumovirus, rhinovirus, adenovirus, parainfluenza, enterovirus, mycoplasma pneumonia, bronchitis, sinusitis, tonsillitis, pertussis, laryngitis, or specific coronavirus strains which are not COVID-19 within the one-month period (±) of respiratory symptoms or receipt of a relevant prescription as noted in the inclusion criteria above (24)</td><td>Inclusion criteria<ul><li>Meets criteria for ‘Narrow’ phenotype, OR,</li><li>ICD-10 diagnosis code for Coronavirus as the cause of diseases classified to other chapters (25) in first or second position in the HES APC, OR,</li><li>ICD-10 diagnosis code for Coronavirus infection, unspecified site (26), in first or second position in the HES APC</li></ul>Exclusion Criteria<br><br>ICD-10 diagnosis code in first or second position in HES APC for RSV, influenza, pneumonia, human metapneumovirus, rhinovirus, adenovirus, parainfluenza, enterovirus, mycoplasma pneumonia, bronchitis, sinusitis, tonsillitis, pertussis, laryngitis, or specific coronavirus strains which are not COVID-19 within the one-month period (±) of respiratory symptoms (27)</td></tr>
</table>

<table>
<tr><th colspan="2">Overall Respiratory Virus</th></tr>
<tr><th>Mild (Primary Care, Emergency Attendances)</th><th>Severe (Secondary Care)</th></tr>
<tr><td colspan="2">Broad</td></tr>
<tr><td>Inclusion criteria<br><br>● Identified from RSV broad inclusion criteria, OR,<br>● Identified from influenza broad inclusion criteria, OR,<br>● Identified from COVID-19 broad inclusion criteria, OR,<br>● Identified from codelist with non-specific conditions associated with respiratory viruses (28), OR,<br>● At least one SNOMED CT code, in first or second position in ECDS (A&E), for LRTI (29) or UTRI (30) - availability: partial 2018-2020; full 2020 onwards (3), OR,<br>● In older adult population, identified as having an exacerbation for COPD (31) or Asthma (32), in SNOMED CT, OR,<br>● In older adult population, acute exacerbation of COPD diagnosis code (33), in SNOMED CT, in first or second position in ECDS (A&E) - availability: partial 2018-2020; full 2020 onwards (3)<br><br>Exclusion criteria<br><br>Diagnosis code in SNOMED CT for metapneumovirus, rhinovirus, adenovirus, parainfluenza, enterovirus, mycoplasma pneumonia or pertussis within the one-month period (±) of respiratory symptoms (34)</td><td>Inclusion criteria<br><br>● Identified from RSV broad inclusion criteria, OR,<br>● Identified from influenza broad inclusion criteria, OR,<br>● Identified from COVID-19 broad inclusion criteria, OR,<br>● Identified from codelist with non-specific conditions associated with respiratory viruses (35), in ICD-10, in first or second position in HES APC, OR,<br>● In older adult population, identified as having an exacerbation for COPD (36) or Asthma (37), in ICD-10, in first or second position in HES APC<br><br>Exclusion criteria<br><br>ICD-10 diagnosis code in first or second position in HES APC for metapneumovirus, rhinovirus, adenovirus, parainfluenza, enterovirus, mycoplasma pneumonia or pertussis within the one-month period (±) of respiratory symptoms (38)</td></tr>
</table>

Table S1: Respiratory virus phenotypes. We used codes from Systematised Nomenclature Of Medicine clinical terms (SNOMED CT), International Classification of Diseases (tenth edition) (ICD-10), pseudo British National Formulary (Pseudo-BNF) - prescription codelists are built using Pseudo BNF and are then mapped onto dictionary of medicines and devices (dm+d) codes which can be identified in patient records.

#### S1.1 Broad Respiratory Attendance

Adapted from the overall respiratory virus phenotype in Table S1, we defined a broad mild respiratory attendance as:

- At least one RSV diagnosis code, or bronchiolitis diagnosis code, in SNOMED CT (1), OR,
- In infant population, at least one bronchiolitis (2) or viral wheeze (6) diagnosis code, in SNOMED CT, in first or second position in ECDS (A&E) - availability: partial 2018-2020; full 2020 onwards (3), OR,
- At least two of the following respiratory symptoms recorded in SNOMED CT within a two-week period (two on the same day is acceptable): rhinitis, cough, fever, breathing abnormalities, and viral infections (5), OR,
- At least one SNOMED CT code for influenza (10), OR,
- At least one SNOMED CT code for influenza, suspected influenza, influenza-like-illness, test run for influenza (12), OR,
- Record of an acute respiratory illness (13) with a measured temperature of ≥ 38 °C (14) within the same episode (adapted from definition of influenza-like-illness according to WHO (15)
- At least one prescription for antivirals (oseltamivir or zanamivir) in Pseudo BNF relevant for the treatment of infectious respiratory diseases (16), OR,
- At least one SNOMED CT code for COVID-19 (20), OR,
- At least one SNOMED CT code for COVID-19, coronavirus (without specific strain), suspected COVID-19, tests performed and referrals to support (21), OR,
- At least two of the following symptoms of COVID-19 recorded in SNOMED CT within a two-week period (two on the same day is acceptable): fever, cough, change to sense of smell or taste, shortness of breath, fatigue, body aches/muscle aches, headache, sore throat, rhinitis, loss of appetite, diarrhoea, nausea and vomiting (22), OR,
- At least one prescription for antivirals (nirmatrelvir plus ritonavir (Paxlovid) and molnupiravir (Lagevrio)) in Pseudo BNF (23), OR,
- Identified from codelist with non-specific conditions associated with respiratory viruses (28), OR,
- At least one SNOMED CT code, in first or second position in ECDS (A&E), for LRTI (29) or UTRI (30) - availability: partial 2018-2020; full 2020 onwards (3), OR,
- In older adult population, identified as having an exacerbation for COPD (31) or Asthma (32), in SNOMED CT
- In older adult population, acute exacerbation of COPD diagnosis code (33), in SNOMED CT, in first or second position in ECDS (A&E) - availability: partial 2018-2020; full 2020 onwards (3)

### S2. Patient Inclusion

#### S2.1 Included Individuals

| *All Age Groups (Older Adults, Adults, Children and Young People, Infants)* | | | | | | | | |
|---|---|---|---|---|---|---|---|---|
| Annual Cohort | 2016-17 | 2017-18 | 2018-19 | 2019-20 | 2020-21 | 2021-22 | 2022-23 | 2023-24 |
| All included | 23117515 | 23643885 | 24063765 | 24341555 | 24666805 | 24724805 | 25043565 | 25230015 |
| *Older Adults* | | | | | | | | |
| Annual Cohort | 2016-17 | 2017-18 | 2018-19 | 2019-20 | 2020-21 | 2021-22 | 2022-23 | 2023-24 |
| Total of correct age (65y+) | 5180475 | 5288135 | 5392415 | 5496195 | 5576255 | 5669865 | 5775645 | 5885775 |
| Not eligible for inclusion | 781080 | 783570 | 793950 | 803990 | 804430 | 832810 | 838810 | 846820 |
| Eligible for inclusion | 4399395 | 4504565 | 4598465 | 4692205 | 4771825 | 4837055 | 4936835 | 5038955 |
| Excluded due to missing sex | 50 | 50 | 50 | 60 | 50 | 50 | 50 | 60 |
| Excluded due to missing IMD | 64320 | 57970 | 58830 | 65830 | 72890 | 82570 | 94000 | 108060 |
| Excluded due to residing in care home | 119170 | 121690 | 123380 | 127450 | 120330 | 126080 | 134990 | 145270 |
| Included in final pop. | 4216445 | 4325295 | 4416765 | 4499625 | 4579645 | 4629765 | 4709865 | 4788275 |
| % of eligible included | 95.84 | 96.02 | 96.05 | 95.90 | 95.97 | 95.71 | 95.40 | 95.03 |
| Adults | | | | | | | | |
| Annual Cohort | 2016-17 | 2017-18 | 2018-19 | 2019-20 | 2020-21 | 2021-22 | 2022-23 | 2023-24 |
| Total of correct age (18-64y) | 21171805 | 21329955 | 21452895 | 21546055 | 21623385 | 21674465 | 21709605 | 21730885 |
| Not eligible for inclusion | 7284670 | 7124210 | 6945330 | 6796730 | 6576240 | 6491870 | 6198970 | 5980740 |
| Eligible for inclusion | 13887135 | 14205745 | 14507565 | 14749325 | 15047145 | 15182595 | 15510635 | 15750145 |
| Excluded due to missing sex | 460 | 510 | 550 | 610 | 670 | 710 | 790 | 870 |
| Excluded due to missing IMD | 178500 | 165510 | 186170 | 239290 | 300890 | 384660 | 470400 | 562590 |
| Excluded due to residing in care home | 22140 | 24160 | 25820 | 27690 | 30600 | 34790 | 38960 | 43450 |
| Included in final pop. | 13686275 | 14015745 | 14295215 | 14481945 | 14715265 | 14762855 | 15001035 | 15144055 |
| % of eligible included | 98.55 | 98.66 | 98.54 | 98.19 | 97.79 | 97.24 | 96.71 | 96.15 |
| *Children and Young People* | | | | | | | | |
| Annual Cohort | 2016-17 | 2017-18 | 2018-19 | 2019-20 | 2020-21 | 2021-22 | 2022-23 | 2023-24 |
| Total of correct age (2-17y) | 6756465 | 6626345 | 6506245 | 6399005 | 6303795 | 6204725 | 6104555 | 6002065 |
| Not eligible for inclusion | 2375370 | 2143230 | 1931270 | 1752310 | 1588710 | 1476750 | 1324790 | 1165480 |
| Eligible for inclusion | 4381095 | 4483115 | 4574975 | 4646695 | 4715085 | 4727975 | 4779765 | 4836585 |

| | | | | | | | | |
|---|---|---|---|---|---|---|---|---|
| Excluded due to missing sex | 530 | 480 | 450 | 400 | 360 | 290 | 260 | 200 |
| Excluded due to missing IMD | 54460 | 55800 | 70210 | 98850 | 130690 | 168470 | 207580 | 252070 |
| Excluded due to residing in care home | 530 | 570 | 570 | 590 | 630 | 830 | 990 | 1290 |
| Included in final pop. | 4325595 | 4426265 | 4503755 | 4546855 | 4583425 | 4558395 | 4570955 | 4583055 |
| % of eligible included | 98.73 | 98.73 | 98.44 | 97.85 | 97.21 | 96.41 | 95.63 | 94.76 |
| *Infants* | | | | | | | | |
| Annual Cohort | 2016-17 | 2017-18 | 2018-19 | 2019-20 | 2020-21 | 2021-22 | 2022-23 | 2023-24 |
| Total of correct age (<2y) | 1038205 | 1012135 | 975405 | 936615 | 899545 | 865955 | 827495 | 789915 |
| Not eligible for inclusion | 134460 | 124120 | 112780 | 103050 | 91840 | 77720 | 60670 | 43140 |
| Eligible for inclusion | 903745 | 888015 | 862625 | 833565 | 807705 | 788235 | 766825 | 746775 |
| Excluded due to missing sex | 10 | 10 | 10 | 10 | 10 | 0 | 0 | 10 |
| Excluded due to missing IMD | 156740 | 150150 | 148730 | 150420 | 144570 | 135950 | 122870 | 113610 |
| Excluded due to residing in care home | 50 | 60 | 60 | 60 | 60 | 70 | 100 | 140 |
| Excluded due to RSV treatment eligibility | 10700 | 14640 | 14880 | 14690 | 13960 | 13770 | 14700 | 16030 |
| Included in final pop. | 738265 | 725225 | 701095 | 670665 | 651345 | 640445 | 631355 | 619505 |
| % of eligible included | 81.69 | 81.67 | 81.27 | 80.46 | 80.64 | 81.25 | 82.33 | 82.96 |
| *Maternally-Linked Infants* | | | | | | | | |
| Annual Cohort | 2016-17 | 2017-18 | 2018-19 | 2019-20 | 2020-21 | 2021-22 | 2022-23 | 2023-24 |
| Total of correct age (<2y) with maternal linkage | 215565 | 218305 | 214425 | 211195 | 203085 | 198585 | 194765 | 141435 |
| Not eligible for inclusion | 52890 | 53080 | 51830 | 50920 | 47390 | 46170 | 44930 | 33400 |
| Eligible for inclusion | 162675 | 165225 | 162595 | 160275 | 155695 | 152415 | 149835 | 108035 |
| Excluded due to missing sex | 0 | 0 | 0 | 0 | 0 | 0 | 0 | 10 |
| Excluded due to missing IMD | 9820 | 10980 | 12620 | 14890 | 15840 | 16410 | 16690 | 11010 |
| Excluded due to residing in care home | 20 | 20 | 30 | 20 | 20 | 10 | 20 | 20 |
| Excluded due to RSV treatment eligibility | 2140 | 3150 | 3320 | 3240 | 3070 | 2970 | 3200 | 2140 |
| Included in final pop. | 150935 | 151355 | 146935 | 142465 | 137125 | 133345 | 130355 | 95125 |
| % of eligible included | 92.78 | 91.61 | 90.37 | 88.89 | 88.07 | 87.49 | 87.00 | 88.05 |

Table S2: Participants included and excluded for each annual cohort, by age group.

#### S2.2 Inclusion Flow Chart

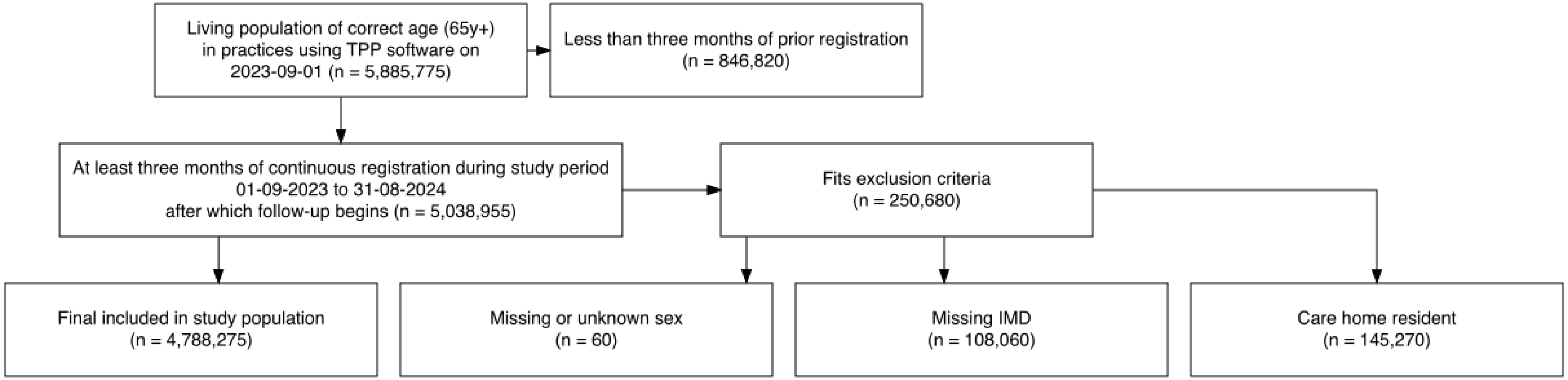

Figure S1: Inclusion flow chart for older adults in 2023-24. The figure shows an example exclusion process, for older adults identified between 01/09/2023 and 31/08/2024. Full data across annual cohorts are provided in Table S1.

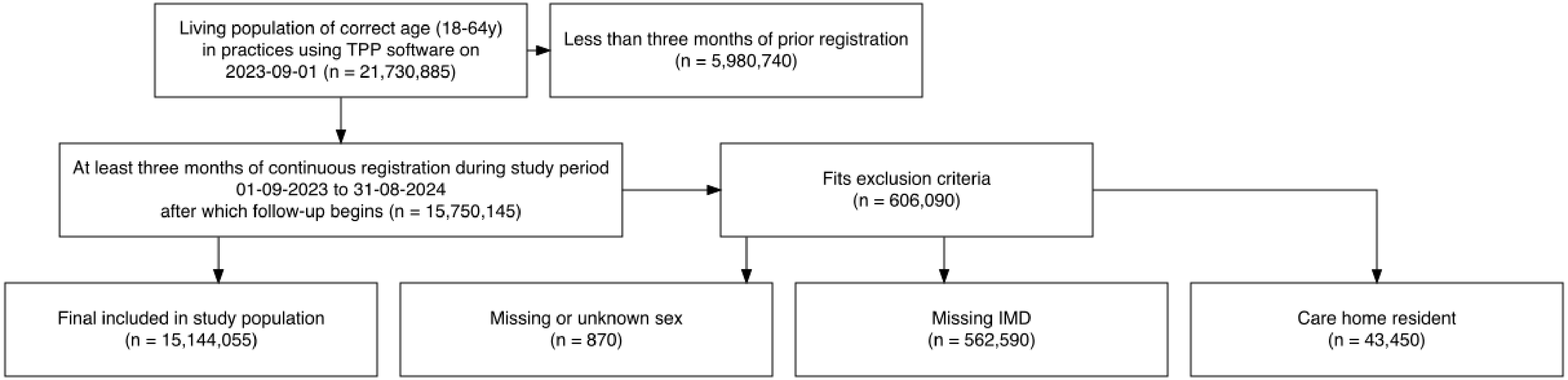

Figure S2: Inclusion flow chart for adults in 2023-24. The figure shows an example exclusion process, for adults identified between 01/09/2023 and 31/08/2024. Full data across annual cohorts are provided in Table S1.

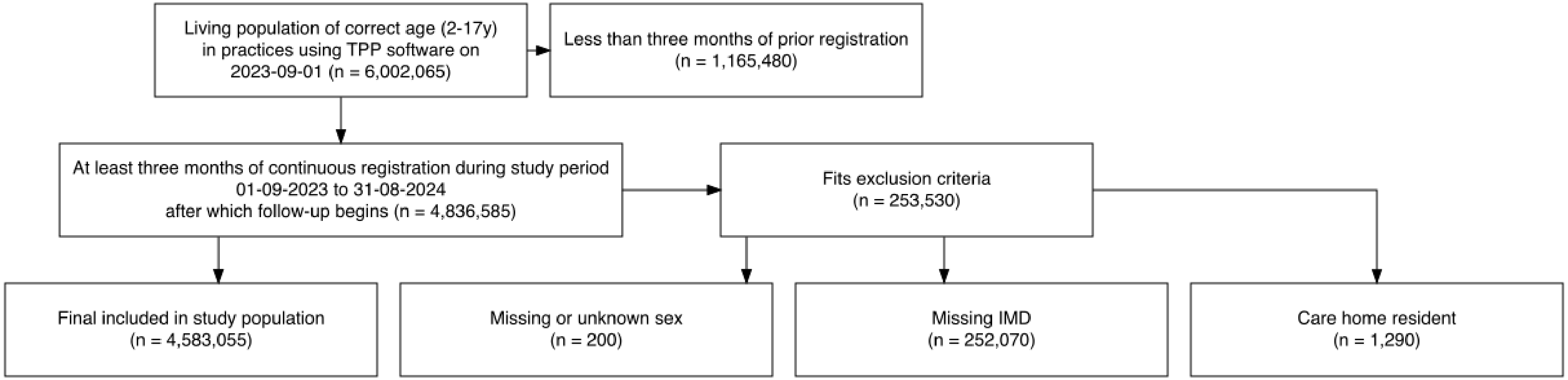

Figure S3: Inclusion flow chart for children and young people in 2023-24. The figure shows an example exclusion process, for children and young people identified between 01/09/2023 and 31/08/2024. Full data across annual cohorts are provided in Table S1.

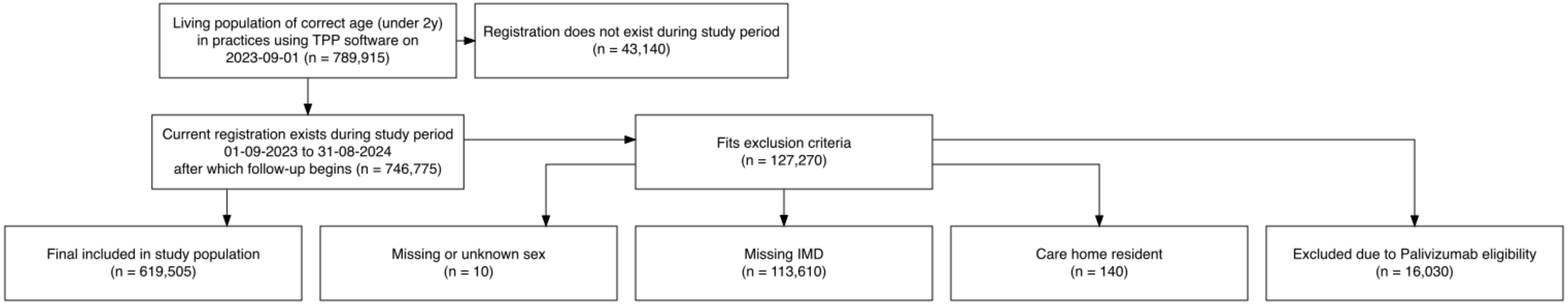

Figure S4: Inclusion flow chart for infants in 2023-24. The figure shows an example exclusion process, for infants identified between 01/09/2023 and 31/08/2024. Full data across annual cohorts are provided in Table S1.

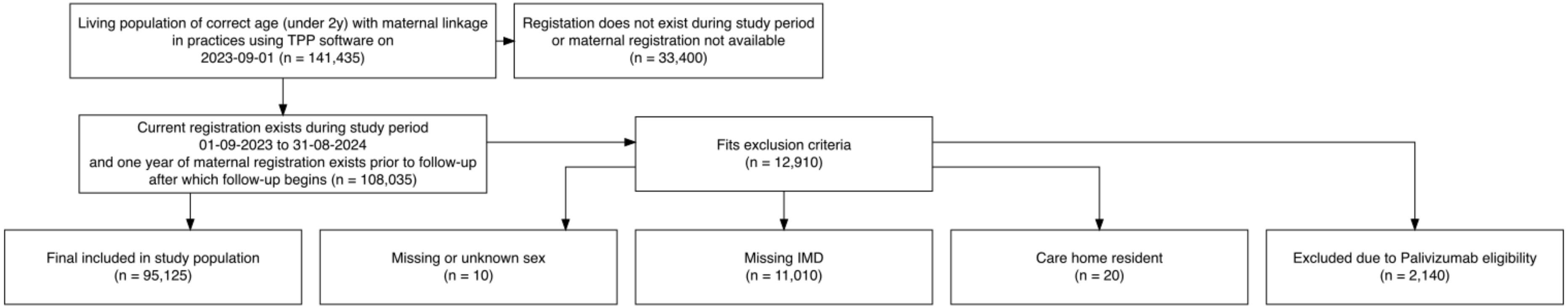

Figure S5: Inclusion flow chart for maternally-linked infants in 2023-24. The figure shows an example exclusion process, for maternally-linked infants identified between 01/09/2023 and 31/08/2024. Full data across annual cohorts are provided in Table S1.

#### S2.3 Participant Characteristics

| *Older Adults* | | | | | | | | | |
|---|---|---|---|---|---|---|---|---|---|
| | Annual Cohort | 2016-17 | 2017-18 | 2018-19 | 2019-20 | 2020-21 | 2021-22 | 2022-23 | 2023-24 |
| *Total* | | 4216445 (100%) | 4325295 (100%) | 4416765 (100%) | 4499625 (100%) | 4579645 (100%) | 4629765 (100%) | 4709865 (100%) | 4788275 (100%) |
| *Age group* | 65-74y | 2461525 (58.4%) | 2521775 (58.3%) | 2561775 (58%) | 2577535 (57.3%) | 2613015 (57.1%) | 2614615 (56.5%) | 2593115 (55.1%) | 2607435 (54.5%) |
| | 75-89y | 1592955 (37.8%) | 1637675 (37.9%) | 1686095 (38.2%) | 1747495 (38.8%) | 1788695 (39.1%) | 1835575 (39.6%) | 1935725 (41.1%) | 2000405 (41.8%) |
| | 90y+ | 161965 (3.8%) | 165835 (3.8%) | 168895 (3.8%) | 174585 (3.9%) | 177935 (3.9%) | 179575 (3.9%) | 181025 (3.8%) | 180445 (3.8%) |
| *Sex* | Female | 2250935 (53.4%) | 2303815 (53.3%) | 2347855 (53.2%) | 2388295 (53.1%) | 2427405 (53%) | 2453385 (53%) | 2493485 (52.9%) | 2533785 (52.9%) |
| | Male | 1965515 (46.6%) | 2021485 (46.7%) | 2068905 (46.8%) | 2111325 (46.9%) | 2152245 (47%) | 2176385 (47%) | 2216385 (47.1%) | 2254495 (47.1%) |
| *Ethnicity* | White | 3856705 (91.5%) | 3955735 (91.5%) | 4034685 (91.3%) | 4103275 (91.2%) | 4165445 (91%) | 4226225 (91.3%) | 4312425 (91.6%) | 4390995 (91.7%) |
| | Mixed | 13355 (0.3%) | 14185 (0.3%) | 15185 (0.3%) | 16075 (0.4%) | 17015 (0.4%) | 17665 (0.4%) | 18825 (0.4%) | 20365 (0.4%) |
| | Asian or Asian British | 109925 (2.6%) | 118375 (2.7%) | 126855 (2.9%) | 134765 (3%) | 142945 (3.1%) | 148795 (3.2%) | 160635 (3.4%) | 170635 (3.6%) |
| | Black or Black British | 31275 (0.7%) | 33065 (0.8%) | 35205 (0.8%) | 37585 (0.8%) | 39955 (0.9%) | 41795 (0.9%) | 45485 (1%) | 50005 (1%) |
| | Chinese or Other | 25665 (0.6%) | 27575 (0.6%) | 29395 (0.7%) | 31425 (0.7%) | 33645 (0.7%) | 35405 (0.8%) | 38875 (0.8%) | 42455 (0.9%) |
| | Unknown | 179515 (4.3%) | 176355 (4.1%) | 175435 (4%) | 176505 (3.9%) | 180635 (3.9%) | 159865 (3.5%) | 133625 (2.8%) | 113825 (2.4%) |

| | | | | | | | | | |
|---|---|---|---|---|---|---|---|---|---|
| *IMD quintile* | 5 (least deprived) | 942725 (22.4%) | 968435 (22.4%) | 990345 (22.4%) | 1010165 (22.4%) | 1031135 (22.5%) | 1045225 (22.6%) | 1064355 (22.6%) | 1081855 (22.6%) |
| | 4 | 980875 (23.3%) | 1007735 (23.3%) | 1030285 (23.3%) | 1050995 (23.4%) | 1070805 (23.4%) | 1085145 (23.4%) | 1105765 (23.5%) | 1125525 (23.5%) |
| | 3 | 946185 (22.4%) | 971325 (22.5%) | 992415 (22.5%) | 1012195 (22.5%) | 1029615 (22.5%) | 1041745 (22.5%) | 1059355 (22.5%) | 1076755 (22.5%) |
| | 2 | 737315 (17.5%) | 756115 (17.5%) | 771955 (17.5%) | 785665 (17.5%) | 799015 (17.4%) | 806015 (17.4%) | 819975 (17.4%) | 833735 (17.4%) |
| | 1 (most deprived) | 609335 (14.5%) | 621665 (14.4%) | 631755 (14.3%) | 640605 (14.2%) | 649065 (14.2%) | 651635 (14.1%) | 660415 (14%) | 670405 (14%) |
| *Vaccinated against influenza in prior annual cohort* | | 2799805 (66.4%) | 2800225 (64.7%) | 2879565 (65.2%) | 2907335 (64.6%) | 3001495 (65.5%) | 3612985 (78%) | 3794175 (80.6%) | 3740495 (78.1%) |

Table S3: Participant characteristics for older adults over all annual cohorts.

| *Adults* | | | | | | | | | |
|---|---|---|---|---|---|---|---|---|---|
| | Annual Cohort | 2016-17 | 2017-18 | 2018-19 | 2019-20 | 2020-21 | 2021-22 | 2022-23 | 2023-24 |
| *Total* | | 13686275 (100%) | 14015745 (100%) | 14295215 (100%) | 14481945 (100%) | 14715265 (100%) | 14762855 (100%) | 15001035 (100%) | 15144055 (100%) |
| *Age group* | 18-39y | 6406185 (46.8%) | 6604895 (47.1%) | 6771915 (47.4%) | 6857405 (47.4%) | 6957775 (47.3%) | 6942205 (47%) | 7093815 (47.3%) | 7181395 (47.4%) |
| | 40-64y | 7280085 (53.2%) | 7410855 (52.9%) | 7523305 (52.6%) | 7624535 (52.6%) | 7757485 (52.7%) | 7820645 (53%) | 7907215 (52.7%) | 7962655 (52.6%) |
| *Sex* | Female | 6731545 (49.2%) | 6885935 (49.1%) | 7015635 (49.1%) | 7094705 (49%) | 7206135 (49%) | 7218675 (48.9%) | 7315825 (48.8%) | 7384665 (48.8%) |
| | Male | 6954725 (50.8%) | 7129815 (50.9%) | 7279585 (50.9%) | 7387235 (51%) | 7509125 (51%) | 7544175 (51.1%) | 7685205 (51.2%) | 7759395 (51.2%) |
| *Ethnicity* | White | 10740085 (78.5%) | 10972705 (78.3%) | 11159485 (78.1%) | 11267525 (77.8%) | 11384365 (77.4%) | 11411545 (77.3%) | 11566755 (77.1%) | 11593475 (76.6%) |
| | Mixed | 176545 (1.3%) | 189485 (1.4%) | 202565 (1.4%) | 213035 (1.5%) | 225705 (1.5%) | 235755 (1.6%) | 254455 (1.7%) | 269915 (1.8%) |
| | Asian or Asian British | 975205 (7.1%) | 1026365 (7.3%) | 1074485 (7.5%) | 1119915 (7.7%) | 1176655 (8%) | 1217855 (8.2%) | 1318625 (8.8%) | 1435755 (9.5%) |
| | Black or Black British | 314675 (2.3%) | 333895 (2.4%) | 352195 (2.5%) | 369255 (2.5%) | 389305 (2.6%) | 403275 (2.7%) | 439375 (2.9%) | 498715 (3.3%) |
| | Chinese or Other | 271405 (2%) | 297295 (2.1%) | 321785 (2.3%) | 344275 (2.4%) | 367485 (2.5%) | 385755 (2.6%) | 433205 (2.9%) | 467055 (3.1%) |
| | Unknown | 1208365 (8.8%) | 1196005 (8.5%) | 1184695 (8.3%) | 1167945 (8.1%) | 1171745 (8%) | 1108665 (7.5%) | 988615 (6.6%) | 879135 (5.8%) |
| *IMD quintile* | 5 (least deprived) | 2481485 (18.1%) | 2522115 (18%) | 2552475 (17.9%) | 2570495 (17.7%) | 2599545 (17.7%) | 2605075 (17.6%) | 2631285 (17.5%) | 2639505 (17.4%) |
| | 4 | 2736785 (20%) | 2793155 (19.9%) | 2839185 (19.9%) | 2869935 (19.8%) | 2909645 (19.8%) | 2923045 (19.8%) | 2964205 (19.8%) | 2977155 (19.7%) |
| | 3 | 2871655 (21%) | 2937695 (21%) | 2994955 (21%) | 3035055 (21%) | 3080425 (20.9%) | 3092435 (20.9%) | 3141725 (20.9%) | 3166995 (20.9%) |
| | 2 | 2765515 (20.2%) | 2840955 (20.3%) | 2912885 (20.4%) | 2960905 (20.4%) | 3018385 (20.5%) | 3026115 (20.5%) | 3087885 (20.6%) | 3128605 (20.7%) |
| | 1 (most deprived) | 2830825 (20.7%) | 2921825 (20.8%) | 2995715 (21%) | 3045545 (21%) | 3107255 (21.1%) | 3116175 (21.1%) | 3175925 (21.2%) | 3231795 (21.3%) |
| *Vaccinated against influenza in prior annual cohort* | | 1357815 (9.9%) | 1370215 (9.8%) | 1464125 (10.2%) | 1420225 (9.8%) | 1420635 (9.7%) | 2806115 (19%) | 3439205 (22.9%) | 3116725 (20.6%) |

Table S4: Participant characteristics for adults over all annual cohorts.

| Children and Young People | | | | | | | | | |
|---|---|---|---|---|---|---|---|---|---|
| | Annual Cohort | 2016-17 | 2017-18 | 2018-19 | 2019-20 | 2020-21 | 2021-22 | 2022-23 | 2023-24 |
| *Total* | | 4325595 (100%) | 4426265 (100%) | 4503755 (100%) | 4546855 (100%) | 4583425 (100%) | 4558395 (100%) | 4570955 (100%) | 4583055 (100%) |
| *Age group* | 2-5y | 1291145 (29.8%) | 1294675 (29.2%) | 1280665 (28.4%) | 1260105 (27.7%) | 1233865 (26.9%) | 1190015 (26.1%) | 1159145 (25.4%) | 1139555 (24.9%) |
| | 6-9y | 1085405 (25.1%) | 1120985 (25.3%) | 1140355 (25.3%) | 1144155 (25.2%) | 1138315 (24.8%) | 1115155 (24.5%) | 1103905 (24.2%) | 1096485 (23.9%) |
| | 10-13y | 1001075 (23.1%) | 1044605 (23.6%) | 1088735 (24.2%) | 1119645 (24.6%) | 1150525 (25.1%) | 1165505 (25.6%) | 1179555 (25.8%) | 1186665 (25.9%) |
| | 14-17y | 947975 (21.9%) | 966005 (21.8%) | 994005 (22.1%) | 1022955 (22.5%) | 1060715 (23.1%) | 1087725 (23.9%) | 1128345 (24.7%) | 1160345 (25.3%) |
| *Sex* | Female | 2101055 (48.6%) | 2151645 (48.6%) | 2190485 (48.6%) | 2212355 (48.7%) | 2231785 (48.7%) | 2220675 (48.7%) | 2227885 (48.7%) | 2233825 (48.7%) |
| | Male | 2224535 (51.4%) | 2274625 (51.4%) | 2313265 (51.4%) | 2334495 (51.3%) | 2351635 (51.3%) | 2337725 (51.3%) | 2343065 (51.3%) | 2349225 (51.3%) |
| *Ethnicity* | White | 3217685 (74.4%) | 3288805 (74.3%) | 3353215 (74.5%) | 3391465 (74.6%) | 3422745 (74.7%) | 3409705 (74.8%) | 3410555 (74.6%) | 3391845 (74%) |
| | Mixed | 126005 (2.9%) | 136355 (3.1%) | 147025 (3.3%) | 156275 (3.4%) | 165585 (3.6%) | 172045 (3.8%) | 180785 (4%) | 189195 (4.1%) |
| | Asian or Asian British | 372685 (8.6%) | 392185 (8.9%) | 408755 (9.1%) | 421765 (9.3%) | 433315 (9.5%) | 441055 (9.7%) | 460595 (10.1%) | 484465 (10.6%) |
| | Black or Black British | 116515 (2.7%) | 123525 (2.8%) | 129395 (2.9%) | 134795 (3%) | 140225 (3.1%) | 142885 (3.1%) | 151845 (3.3%) | 172505 (3.8%) |
| | Chinese or Other | 86375 (2%) | 92905 (2.1%) | 98685 (2.2%) | 103195 (2.3%) | 107515 (2.3%) | 110775 (2.4%) | 120455 (2.6%) | 128755 (2.8%) |
| | Unknown | 406325 (9.4%) | 392485 (8.9%) | 366675 (8.1%) | 339365 (7.5%) | 314025 (6.9%) | 281945 (6.2%) | 246725 (5.4%) | 216295 (4.7%) |
| *IMD quintile* | 5 (least deprived) | 784195 (18.1%) | 795205 (18%) | 802605 (17.8%) | 805145 (17.7%) | 807185 (17.6%) | 801605 (17.6%) | 800005 (17.5%) | 797565 (17.4%) |
| | 4 | 809145 (18.7%) | 824865 (18.6%) | 837145 (18.6%) | 844815 (18.6%) | 851245 (18.6%) | 846095 (18.6%) | 849555 (18.6%) | 848815 (18.5%) |
| | 3 | 838955 (19.4%) | 857715 (19.4%) | 872675 (19.4%) | 882105 (19.4%) | 889485 (19.4%) | 885775 (19.4%) | 889065 (19.5%) | 892805 (19.5%) |
| | 2 | 850985 (19.7%) | 873395 (19.7%) | 891505 (19.8%) | 902025 (19.8%) | 910725 (19.9%) | 906175 (19.9%) | 910725 (19.9%) | 916575 (20%) |
| | 1 (most deprived) | 1042315 (24.1%) | 1075085 (24.3%) | 1099825 (24.4%) | 1112755 (24.5%) | 1124775 (24.5%) | 1118735 (24.5%) | 1121595 (24.5%) | 1127285 (24.6%) |
| *Vaccinated against influenza in prior annual cohort* | | 752725 (17.4%) | 914815 (20.7%) | 1148695 (25.5%) | 1260875 (27.7%) | 1362655 (29.7%) | 1602065 (35.1%) | 1880175 (41.1%) | 1628095 (35.5%) |

Table S5: Participant characteristics for children and young people over all annual cohorts.

| *Infants* | | | | | | | | | |
|---|---|---|---|---|---|---|---|---|---|
| Annual Cohort | | 2016-17 | 2017-18 | 2018-19 | 2019-20 | 2020-21 | 2021-22 | 2022-23 | 2023-24 |
| *Total (infants)* | | 738265 (100%) | 725225 (100%) | 701095 (100%) | 670665 (100%) | 651345 (100%) | 640445 (100%) | 631355 (100%) | 619505 |
| *Total (infant months)* | | 5893615 (100%) | 5774905 (100%) | 5576935 (100%) | 5345245 (100%) | 5163575 (100%) | 5073775 (100%) | 5031195 (100%) | 4829685 (100%) |
| *Age group* | 0-2m | 620205 (10.5%) | 600545 (10.4%) | 579215 (10.4%) | 555655 (10.4%) | 533965 (10.3%) | 542825 (10.7%) | 526195 (10.5%) | 529935 (11%) |
| | 3-5m | 784365 (13.3%) | 763475 (13.2%) | 735545 (13.2%) | 694875 (13%) | 676075 (13.1%) | 688505 (13.6%) | 652905 (13%) | 628965 (13%) |
| | 6-11m | 1504685 (25.5%) | 1457795 (25.2%) | 1397985 (25.1%) | 1336455 (25%) | 1290355 (25%) | 1290395 (25.4%) | 1251835 (24.9%) | 1181895 (24.5%) |
| | 12-23m | 2984355 (50.6%) | 2953095 (51.1%) | 2864195 (51.4%) | 2758265 (51.6%) | 2663185 (51.6%) | 2552055 (50.3%) | 2600255 (51.7%) | 2488875 (51.5%) |
| *Sex* | Female | 2876915 (48.8%) | 2823125 (48.9%) | 2726915 (48.9%) | 2615555 (48.9%) | 2528075 (49%) | 2484705 (49%) | 2462975 (49%) | 2367265 (49%) |
| | Male | 3016705 (51.2%) | 2951785 (51.1%) | 2850015 (51.1%) | 2729685 (51.1%) | 2635505 (51%) | 2589075 (51%) | 2568215 (51%) | 2462415 (51%) |
| *Ethnicity* | White | 4354735 (73.9%) | 4410135 (76.4%) | 4267935 (76.5%) | 4065135 (76.1%) | 3891525 (75.4%) | 3782955 (74.6%) | 3674205 (73%) | 3409545 (70.6%) |
| | Mixed | 258725 (4.4%) | 277835 (4.8%) | 282875 (5.1%) | 283325 (5.3%) | 282725 (5.5%) | 287155 (5.7%) | 290355 (5.8%) | 284745 (5.9%) |
| | Asian or Asian British | 546225 (9.3%) | 565565 (9.8%) | 551915 (9.9%) | 544975 (10.2%) | 557125 (10.8%) | 578855 (11.4%) | 614625 (12.2%) | 659875 (13.7%) |
| | Black or Black British | 162715 (2.8%) | 173845 (3%) | 173105 (3.1%) | 170115 (3.2%) | 166875 (3.2%) | 172055 (3.4%) | 202335 (4%) | 240345 (5%) |
| | Chinese or Other | 147585 (2.5%) | 157075 (2.7%) | 154825 (2.8%) | 150245 (2.8%) | 146325 (2.8%) | 145645 (2.9%) | 147905 (2.9%) | 141325 (2.9%) |
| | Unknown | 423635 (7.2%) | 190445 (3.3%) | 146275 (2.6%) | 131455 (2.5%) | 119005 (2.3%) | 107095 (2.1%) | 101755 (2%) | 93855 (1.9%) |
| *IMD quintile* | 5 (least deprived) | 913915 (15.5%) | 892455 (15.5%) | 861335 (15.4%) | 825635 (15.4%) | 797135 (15.4%) | 795855 (15.7%) | 781245 (15.5%) | 731835 (15.2%) |
| | 4 | 1058205 (18%) | 1042795 (18.1%) | 1009665 (18.1%) | 965425 (18.1%) | 932735 (18.1%) | 930455 (18.3%) | 916035 (18.2%) | 863415 (17.9%) |
| | 3 | 1168445 (19.8%) | 1142565 (19.8%) | 1104845 (19.8%) | 1062685 (19.9%) | 1029315 (19.9%) | 1012455 (20%) | 1003245 (19.9%) | 958065 (19.8%) |
| | 2 | 1242605 (21.1%) | 1216975 (21.1%) | 1179695 (21.2%) | 1127785 (21.1%) | 1090505 (21.1%) | 1067185 (21%) | 1064475 (21.2%) | 1033445 (21.4%) |
| | 1 (most deprived) | 1510445 (25.6%) | 1480125 (25.6%) | 1421395 (25.5%) | 1363715 (25.5%) | 1313895 (25.4%) | 1267815 (25%) | 1266185 (25.2%) | 1242915 (25.7%) |

Table S6: Participant characteristics for infants over all annual cohorts.

| *Maternally-Linked Infants* | | | | | | | | | |
|---|---|---|---|---|---|---|---|---|---|
| | Annual Cohort | 2016-17 | 2017-18 | 2018-19 | 2019-20 | 2020-21 | 2021-22 | 2022-23 | 2023-24 |
| *Total (infants)* | | 150935 (100%) | 151355 (100%) | 146935 (100%) | 142465 (100%) | 137125 (100%) | 133345 (100%) | 130355 (100%) | 95125 (100%) |
| *Total (infants months)* | | 1222645 (100%) | 1211995 (100%) | 1185965 (100%) | 1139315 (100%) | 1096465 (100%) | 1062235 (100%) | 1042245 (100%) | 864375 (100%) |
| *Age group* | 0-2m | 131135 (10.7%) | 130335 (10.8%) | 123095 (10.4%) | 118835 (10.4%) | 114215 (10.4%) | 112315 (10.6%) | 113435 (10.9%) | 37865 (4.4%) |
| | 3-5m | 165415 (13.5%) | 162085 (13.4%) | 157705 (13.3%) | 148095 (13%) | 144785 (13.2%) | 142145 (13.4%) | 139675 (13.4%) | 78875 (9.1%) |
| | 6-11m | 313045 (25.6%) | 302205 (24.9%) | 303275 (25.6%) | 280355 (24.6%) | 274365 (25%) | 269395 (25.4%) | 261055 (25%) | 230745 (26.7%) |
| | 12-23m | 613045 (50.1%) | 617375 (50.9%) | 601895 (50.8%) | 592035 (52%) | 563095 (51.4%) | 538385 (50.7%) | 528085 (50.7%) | 516905 (59.8%) |
| *Sex* | Female | 598035 (48.9%) | 595045 (49.1%) | 582035 (49.1%) | 559145 (49.1%) | 538595 (49.1%) | 521075 (49.1%) | 513305 (49.2%) | 426285 (49.3%) |
| | Male | 624615 (51.1%) | 616955 (50.9%) | 603935 (50.9%) | 580165 (50.9%) | 557875 (50.9%) | 541165 (50.9%) | 528945 (50.8%) | 438095 (50.7%) |
| *Ethnicity* | White | 943825 (77.2%) | 956825 (78.9%) | 935425 (78.9%) | 895445 (78.6%) | 855065 (78%) | 821595 (77.3%) | 800645 (76.8%) | 659665 (76.3%) |
| | Mixed | 55535 (4.5%) | 59445 (4.9%) | 61615 (5.2%) | 61005 (5.4%) | 60635 (5.5%) | 61045 (5.7%) | 60975 (5.9%) | 50935 (5.9%) |
| | Asian or Asian British | 96915 (7.9%) | 103915 (8.6%) | 102835 (8.7%) | 99295 (8.7%) | 100455 (9.2%) | 104395 (9.8%) | 107765 (10.3%) | 91975 (10.6%) |
| | Black or Black British | 33745 (2.8%) | 36495 (3%) | 36675 (3.1%) | 34625 (3%) | 31915 (2.9%) | 30765 (2.9%) | 31905 (3.1%) | 28905 (3.3%) |
| | Chinese or Other | 27645 (2.3%) | 28905 (2.4%) | 29895 (2.5%) | 29175 (2.6%) | 28615 (2.6%) | 27935 (2.6%) | 27095 (2.6%) | 22055 (2.6%) |
| | Unknown | 64975 (5.3%) | 26425 (2.2%) | 19515 (1.6%) | 19765 (1.7%) | 19785 (1.8%) | 16495 (1.6%) | 13845 (1.3%) | 10835 (1.3%) |
| *IMD quintile* | 5 (least deprived) | 179825 (14.7%) | 176625 (14.6%) | 174815 (14.7%) | 168635 (14.8%) | 161145 (14.7%) | 159395 (15%) | 158395 (15.2%) | 134645 (15.6%) |
| | 4 | 227405 (18.6%) | 222285 (18.3%) | 217115 (18.3%) | 208805 (18.3%) | 203015 (18.5%) | 198345 (18.7%) | 195415 (18.7%) | 161985 (18.7%) |
| | 3 | 255995 (20.9%) | 257485 (21.2%) | 250905 (21.2%) | 240785 (21.1%) | 233105 (21.3%) | 229915 (21.6%) | 228545 (21.9%) | 187275 (21.7%) |
| | 2 | 267505 (21.9%) | 269695 (22.3%) | 263975 (22.3%) | 248815 (21.8%) | 239825 (21.9%) | 230555 (21.7%) | 223135 (21.4%) | 182965 (21.2%) |
| | 1 (most deprived) | 291915 (23.9%) | 285915 (23.6%) | 279165 (23.5%) | 272265 (23.9%) | 259375 (23.7%) | 244025 (23%) | 236755 (22.7%) | 197505 (22.8%) |

Table S7: Participant characteristics for maternally-linked infants over all annual cohorts.

#### S3. Phenotype Sensitivity

##### S3.1 Older Adults

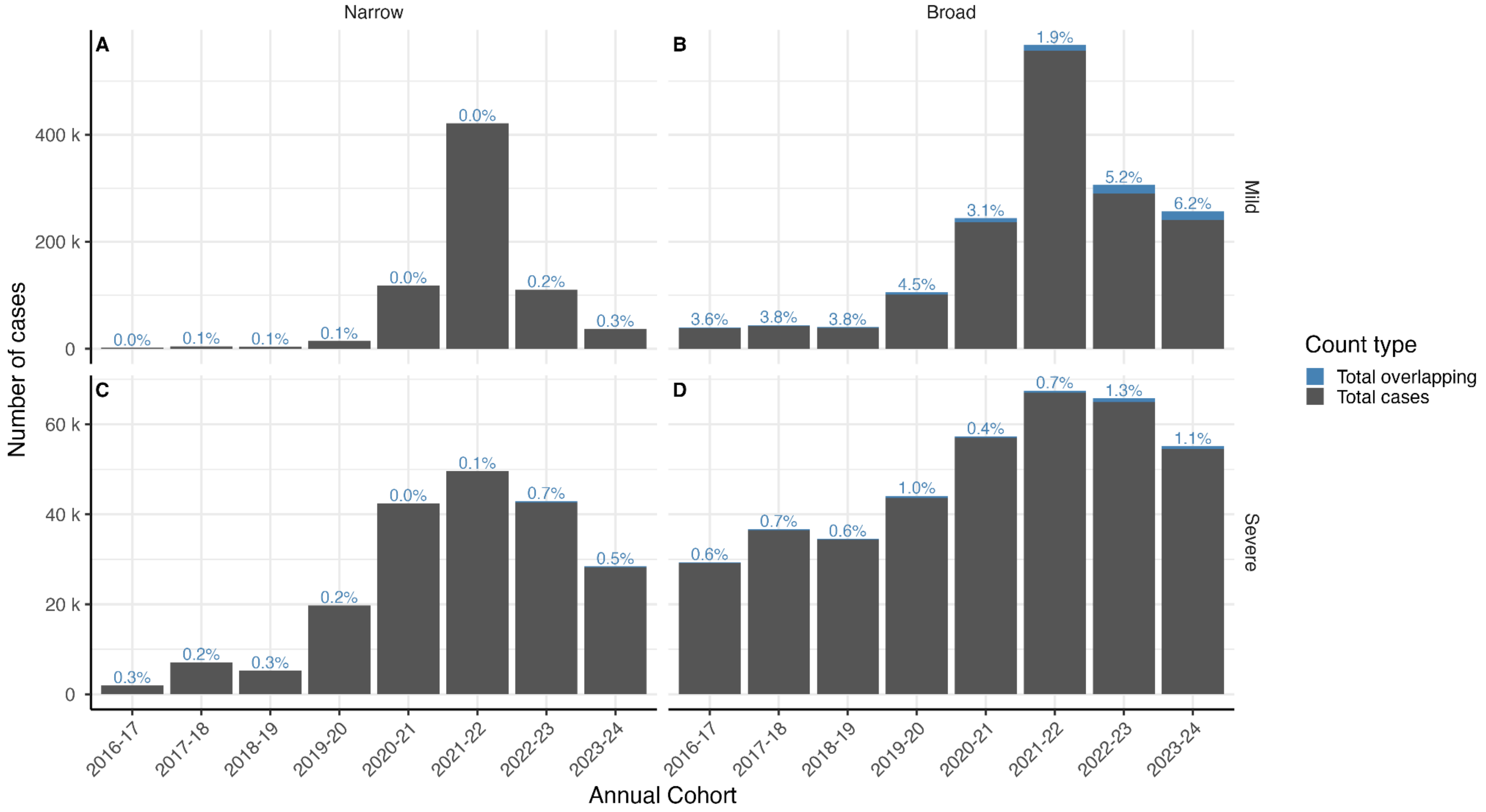

Figure S6: Number of cases in older adults which are identified solely as one case (grey) or as multiple cases (blue), with percentage of cases overlapping above the bar. Results are shown separately for each case severity, phenotype, and annual cohort.

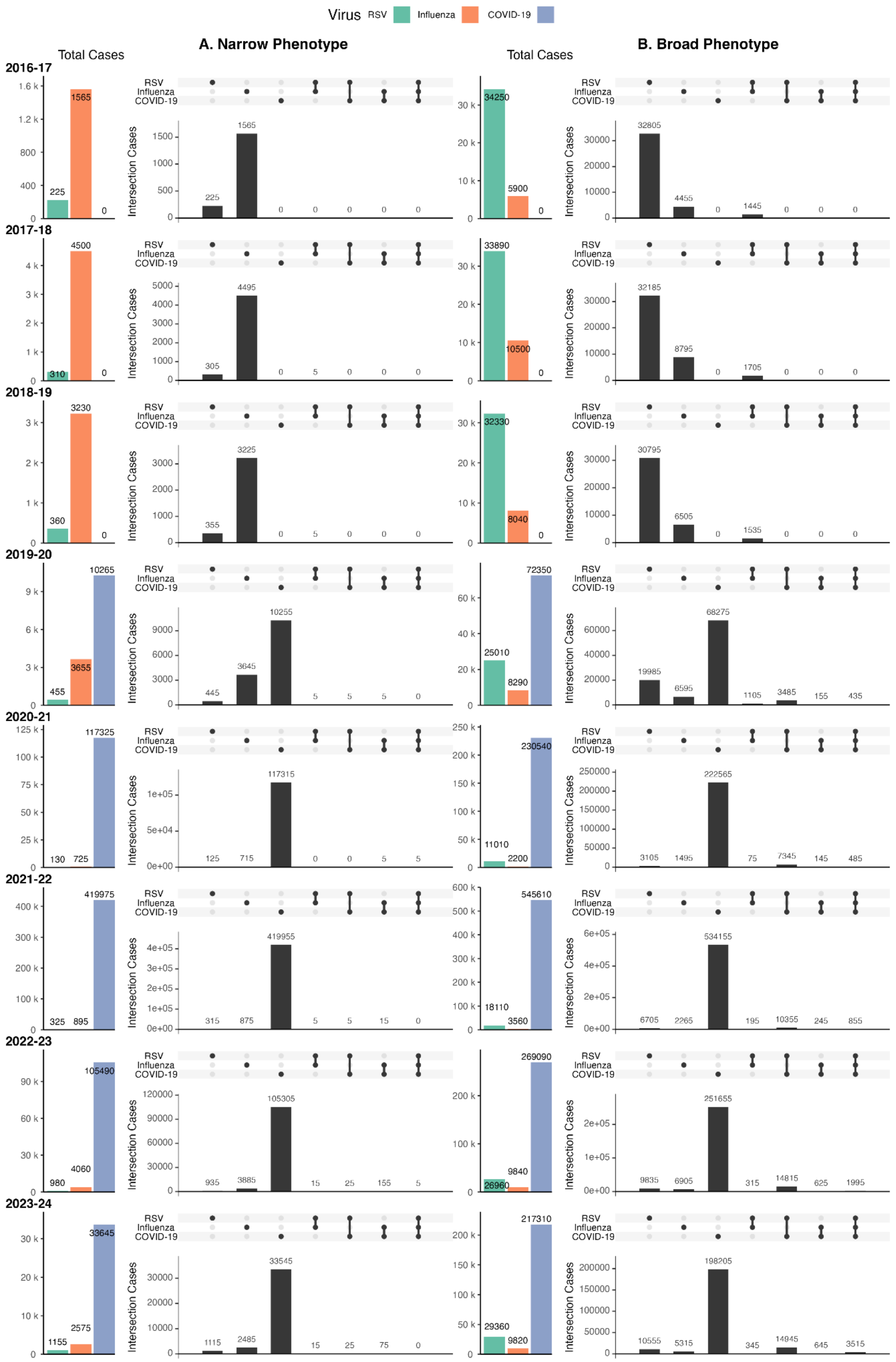

Figure S7: Overlap of mild case identification in older adults, comparing narrow phenotypes (A) and broad phenotypes (B). Cases can be RSV (green), influenza (orange), or COVID-19 (purple).

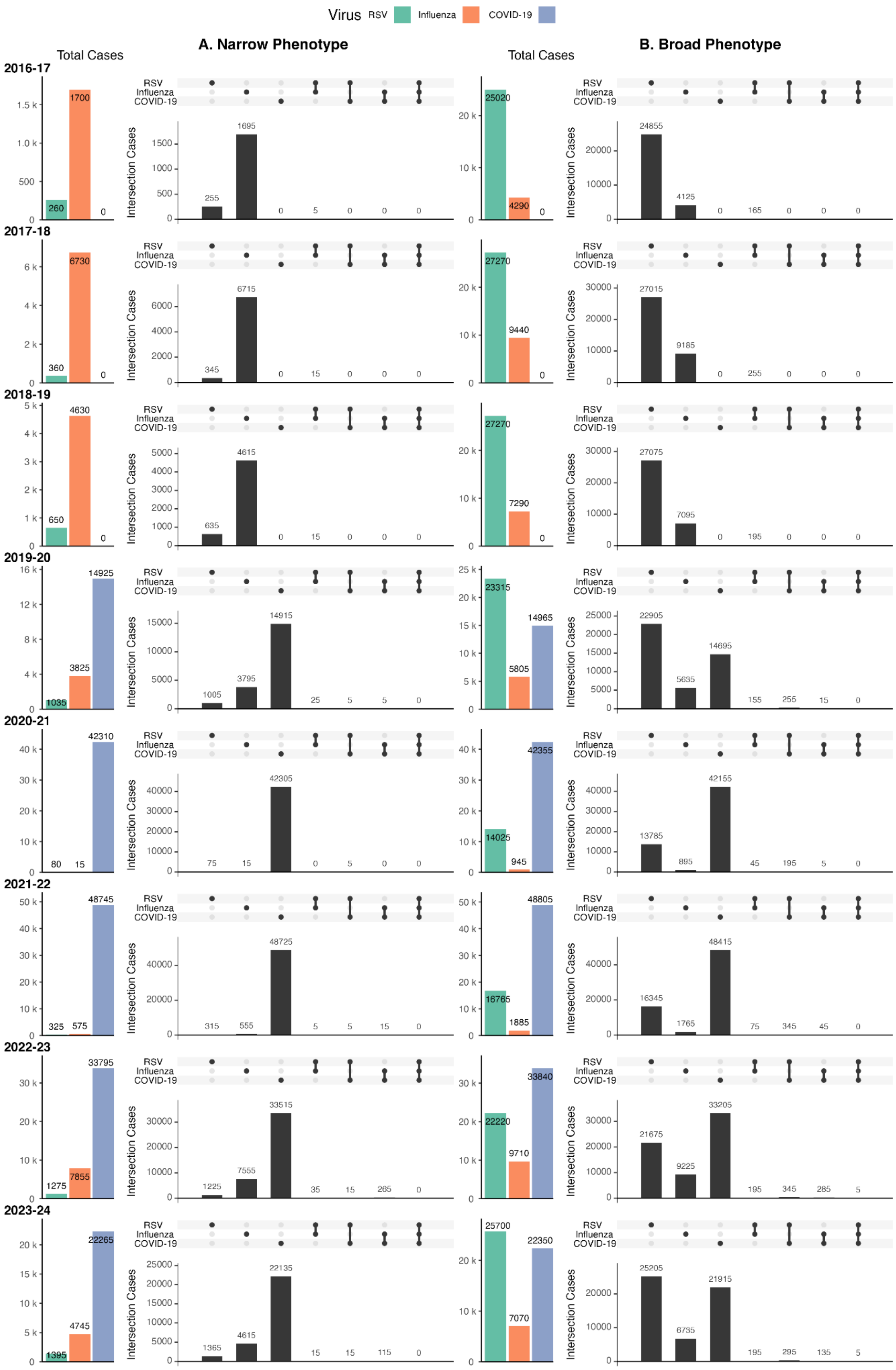

Figure S8: Overlap of severe case identification in older adults, comparing narrow phenotypes (A) and broad phenotypes (B). Cases can be RSV (green), influenza (orange), or COVID-19 (purple).

#### S3.2 Adults

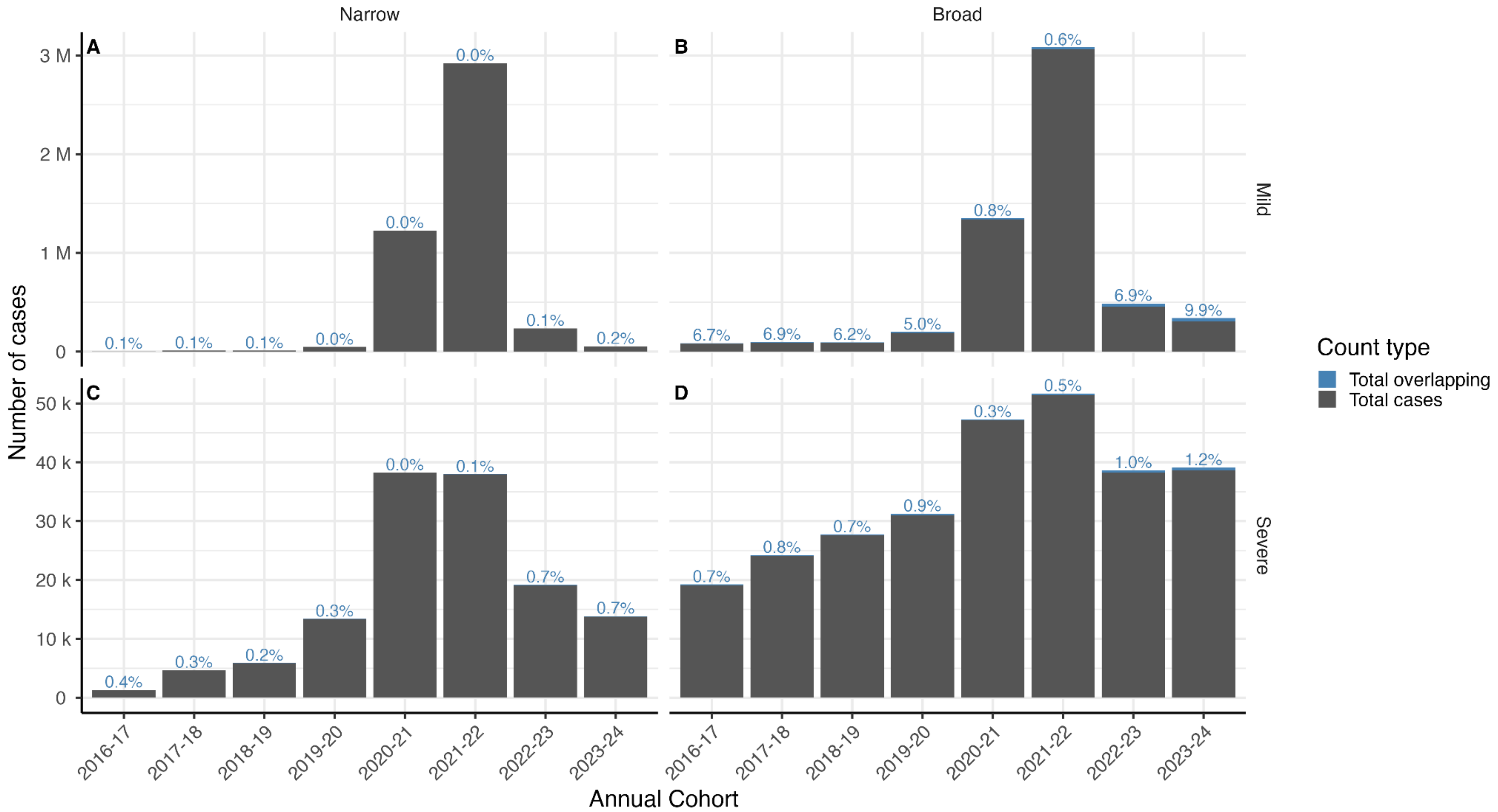

Figure S9: Number of cases in adults which are identified solely as one case (grey) or as multiple cases (blue), with percentage of cases overlapping above the bar. Results are shown separately for each case severity, phenotype, and annual cohort.

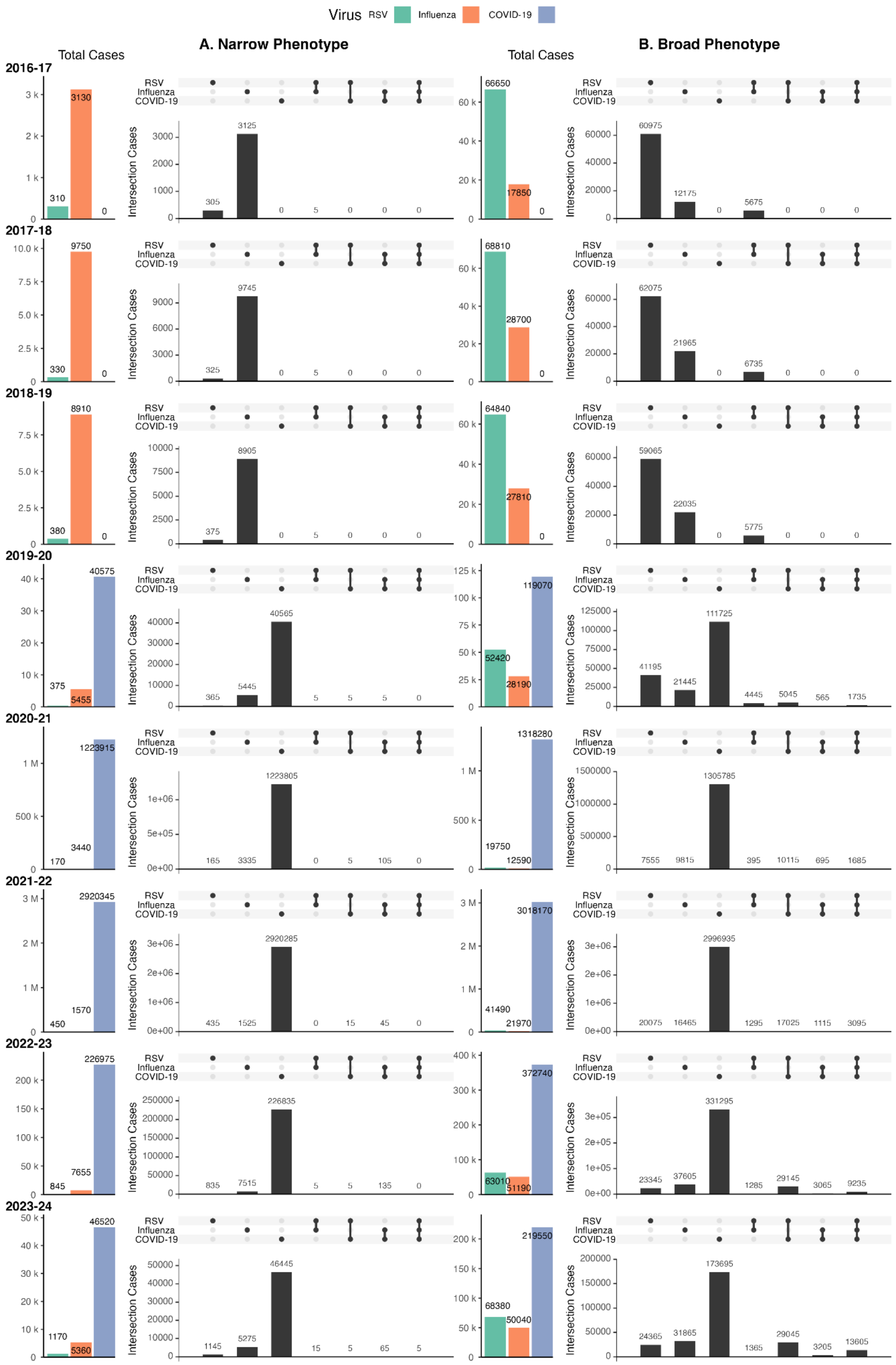

Figure S10: Overlap of mild case identification in adults, comparing narrow phenotypes (A) and broad phenotypes (B). Cases can be RSV (green), influenza (orange), or COVID-19 (purple).

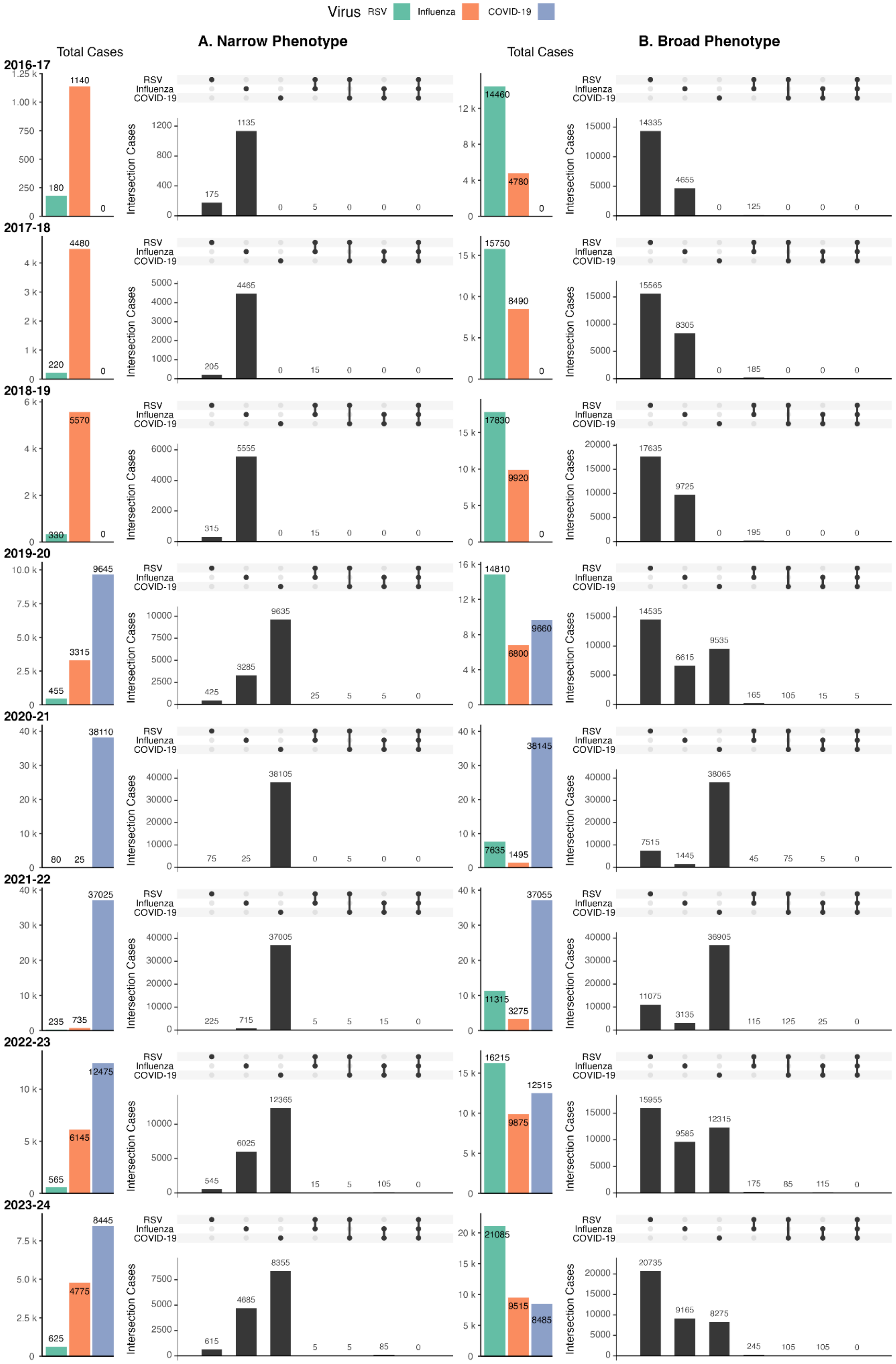

Figure S11: Overlap of severe case identification in adults, comparing narrow phenotypes (A) and broad phenotypes (B). Cases can be RSV (green), influenza (orange), or COVID-19 (purple).

#### S3.3 Children and Young People

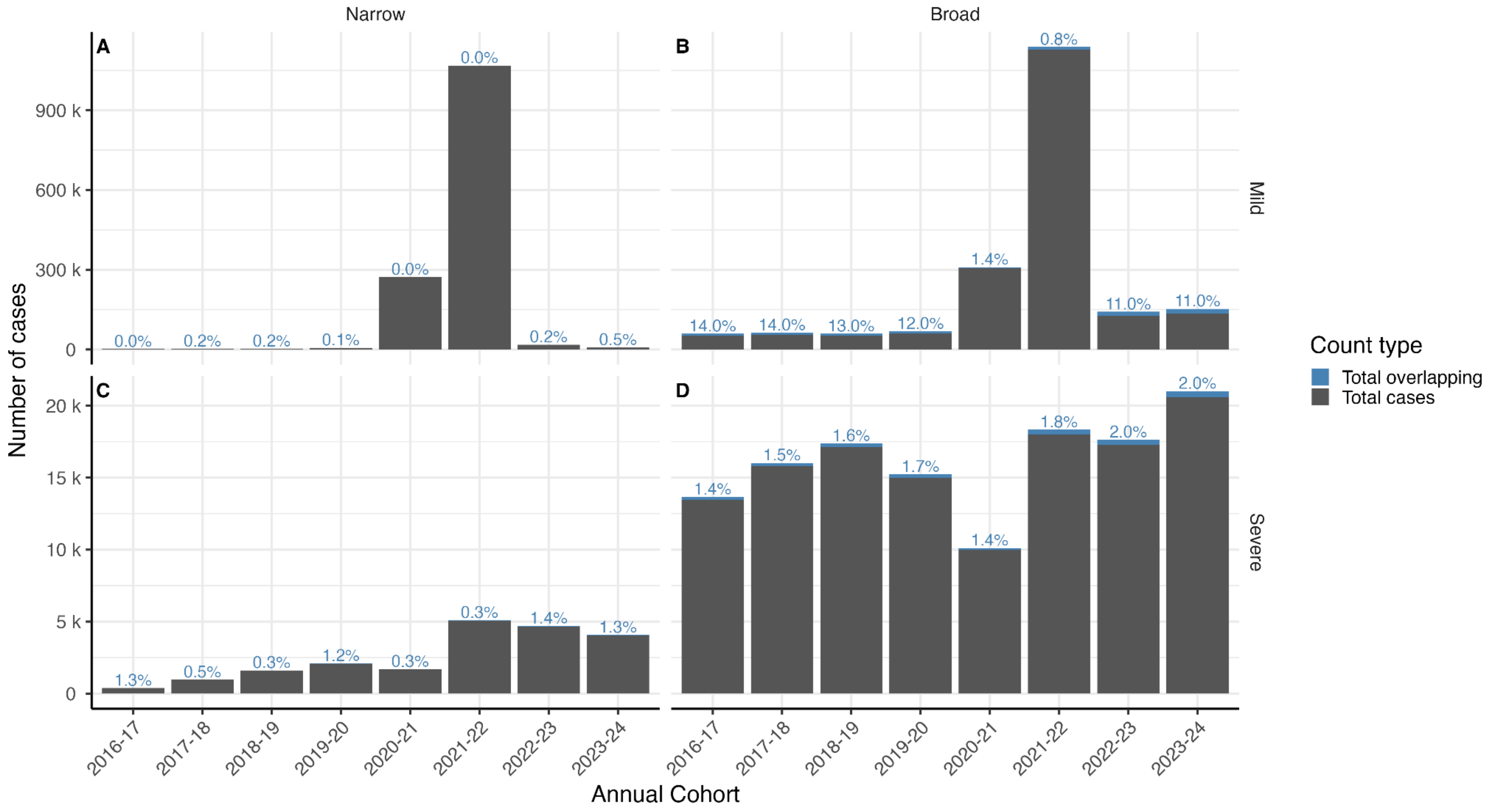

Figure S12: Number of cases in children and young people which are identified solely as one pathogen (grey) or as multiple pathogens (blue), with percentage of cases overlapping above the bar. Results are shown separately for each case severity, phenotype, and annual cohort.

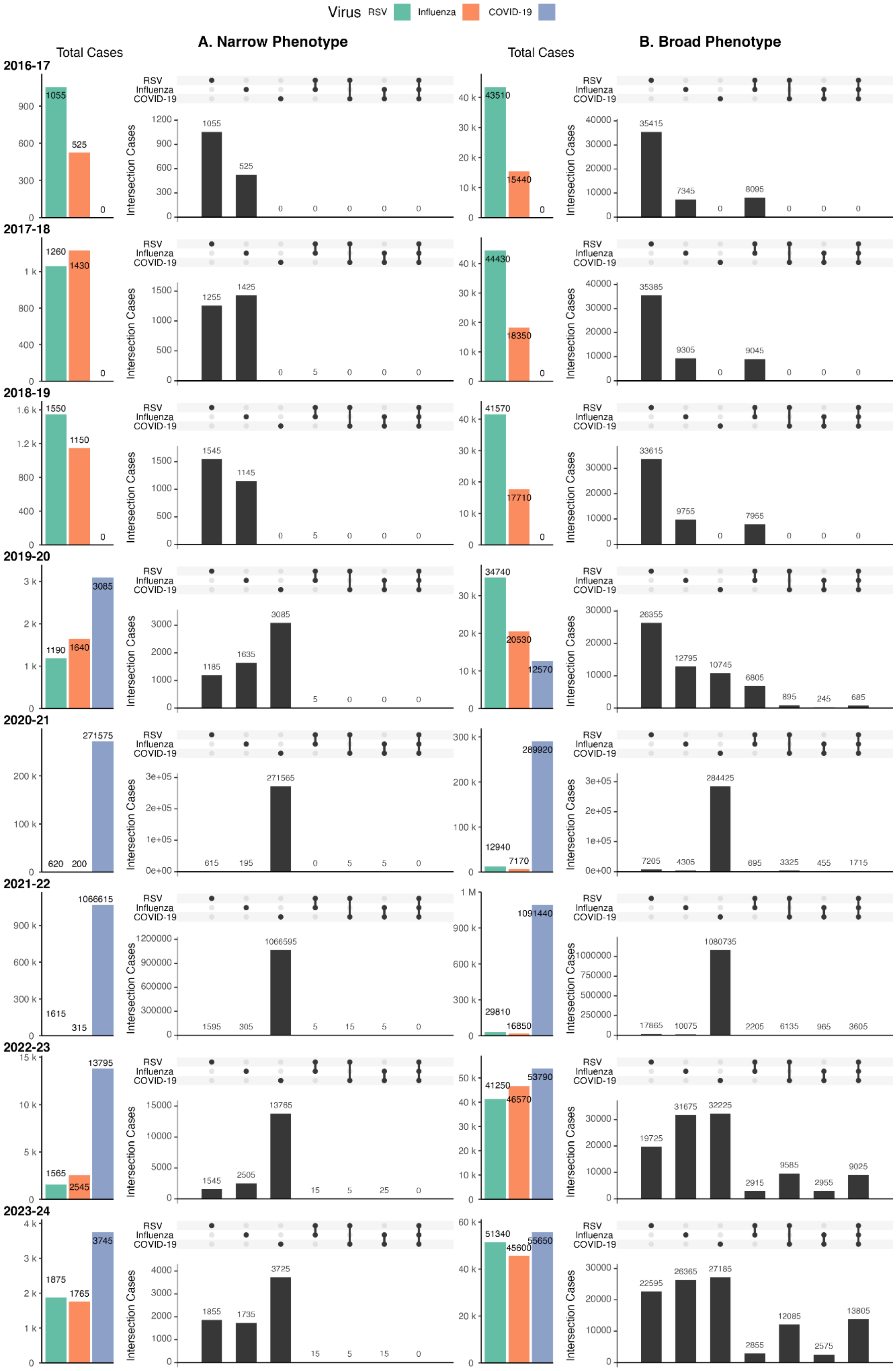

Figure S13: Overlap of mild case identification in children and young people, comparing narrow phenotypes (A) and broad phenotypes (B). Cases can be RSV (green), influenza (orange), or COVID-19 (purple).

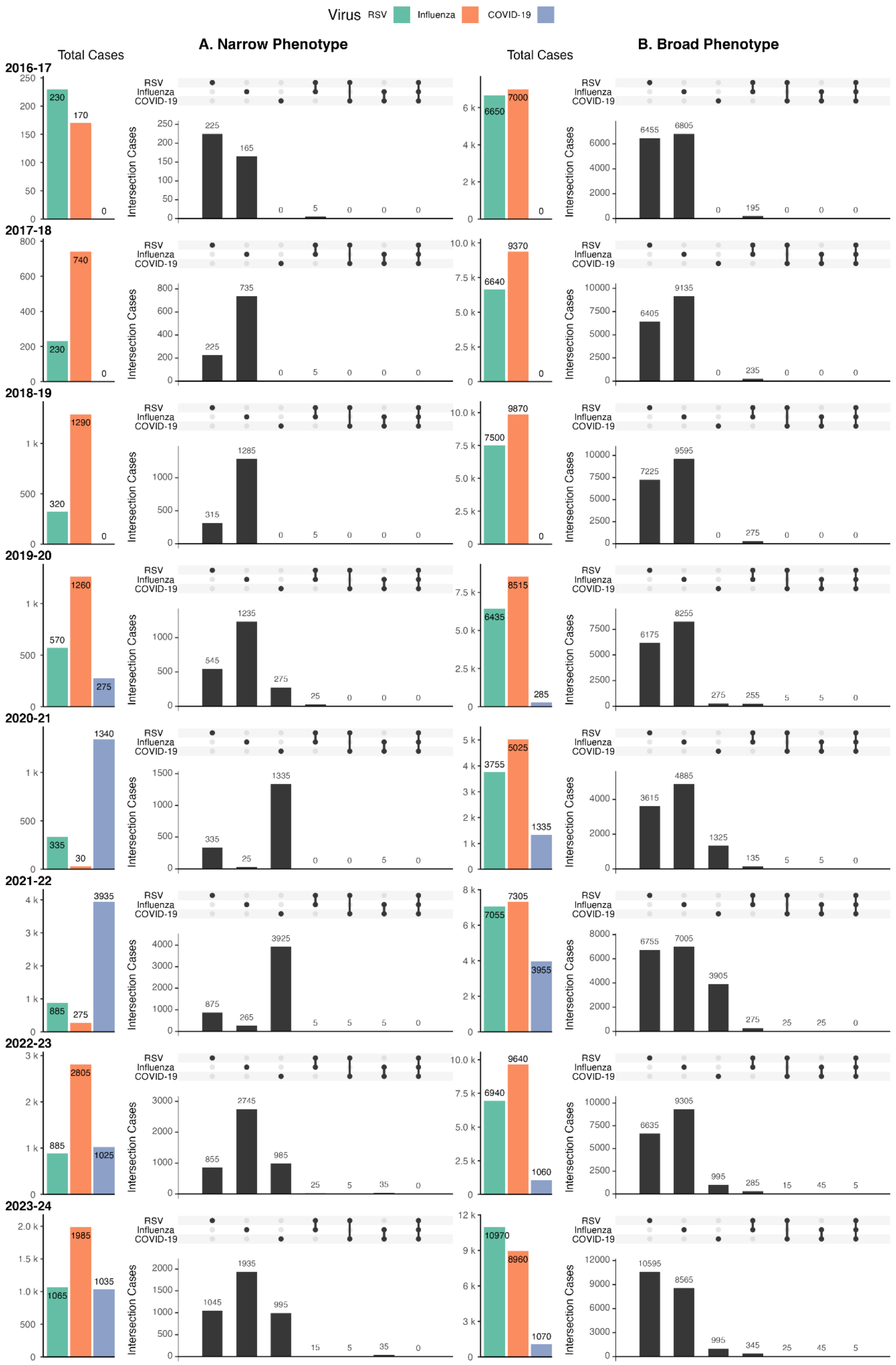

Figure S14: Overlap of severe case identification in children and young people, comparing narrow phenotypes (A) and broad phenotypes (B). Cases can be RSV (green), influenza (orange), or COVID-19 (purple).

#### S3.4 Infants

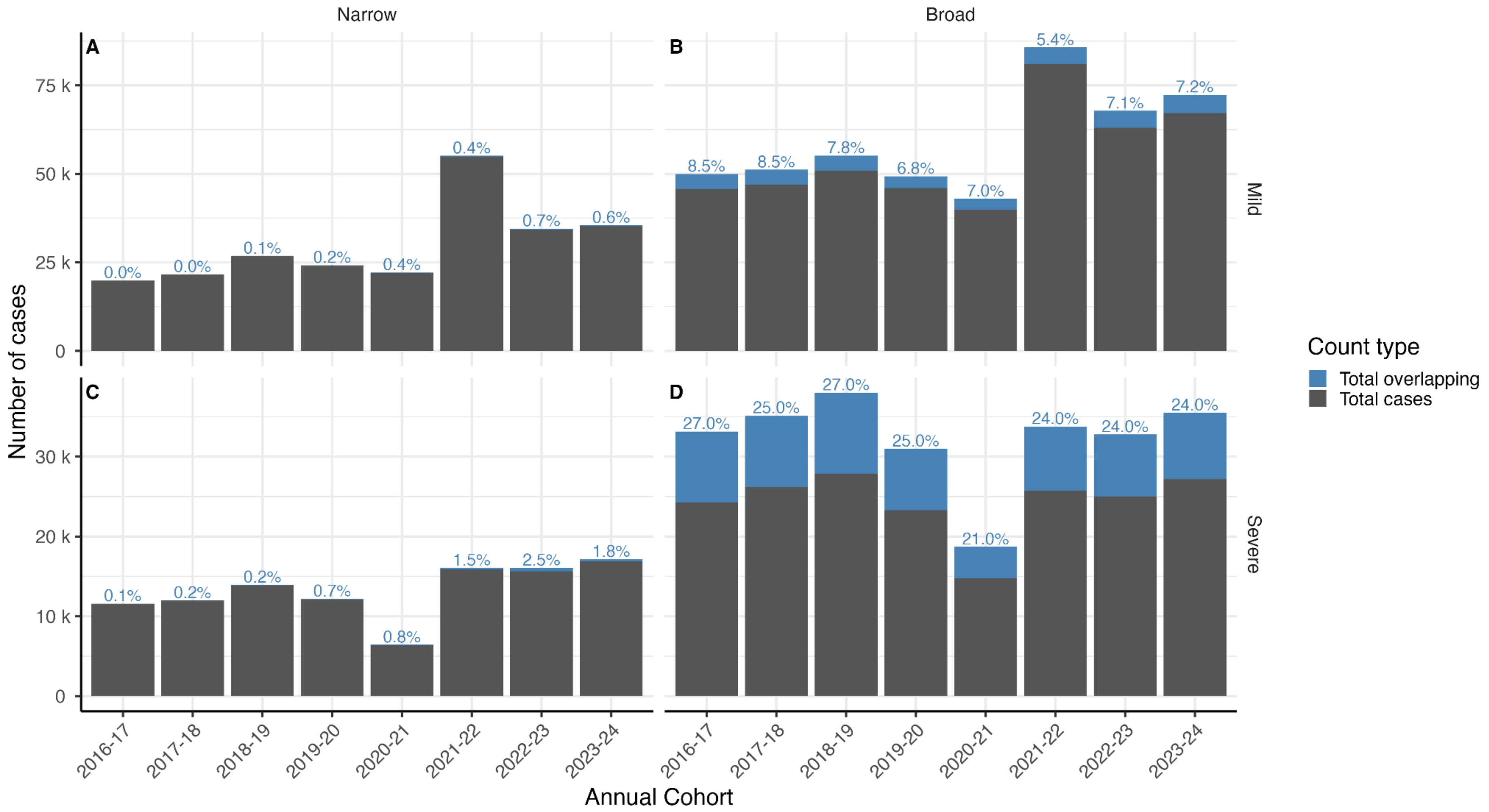

Figure S15: Number of cases in infants which are identified solely as one pathogen (grey) or as multiple pathogens (blue), with percentage of cases overlapping above the bar. Results are shown separately for each case severity, phenotype, and annual cohort.

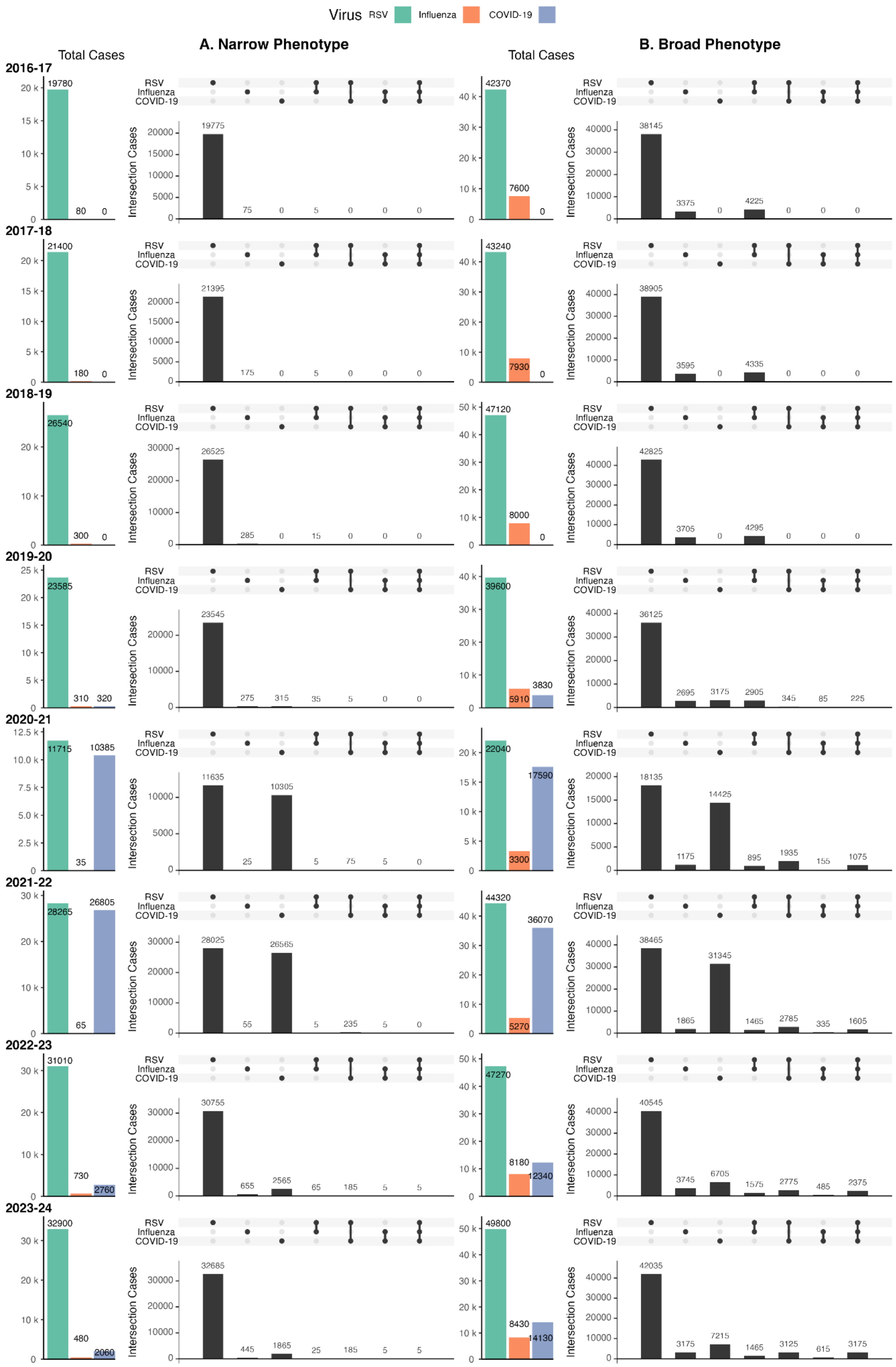

Figure S16: Overlap of mild case identification in infants, comparing narrow phenotypes (A) and broad phenotypes (B). Cases can be RSV (green), influenza (orange), or COVID-19 (purple).

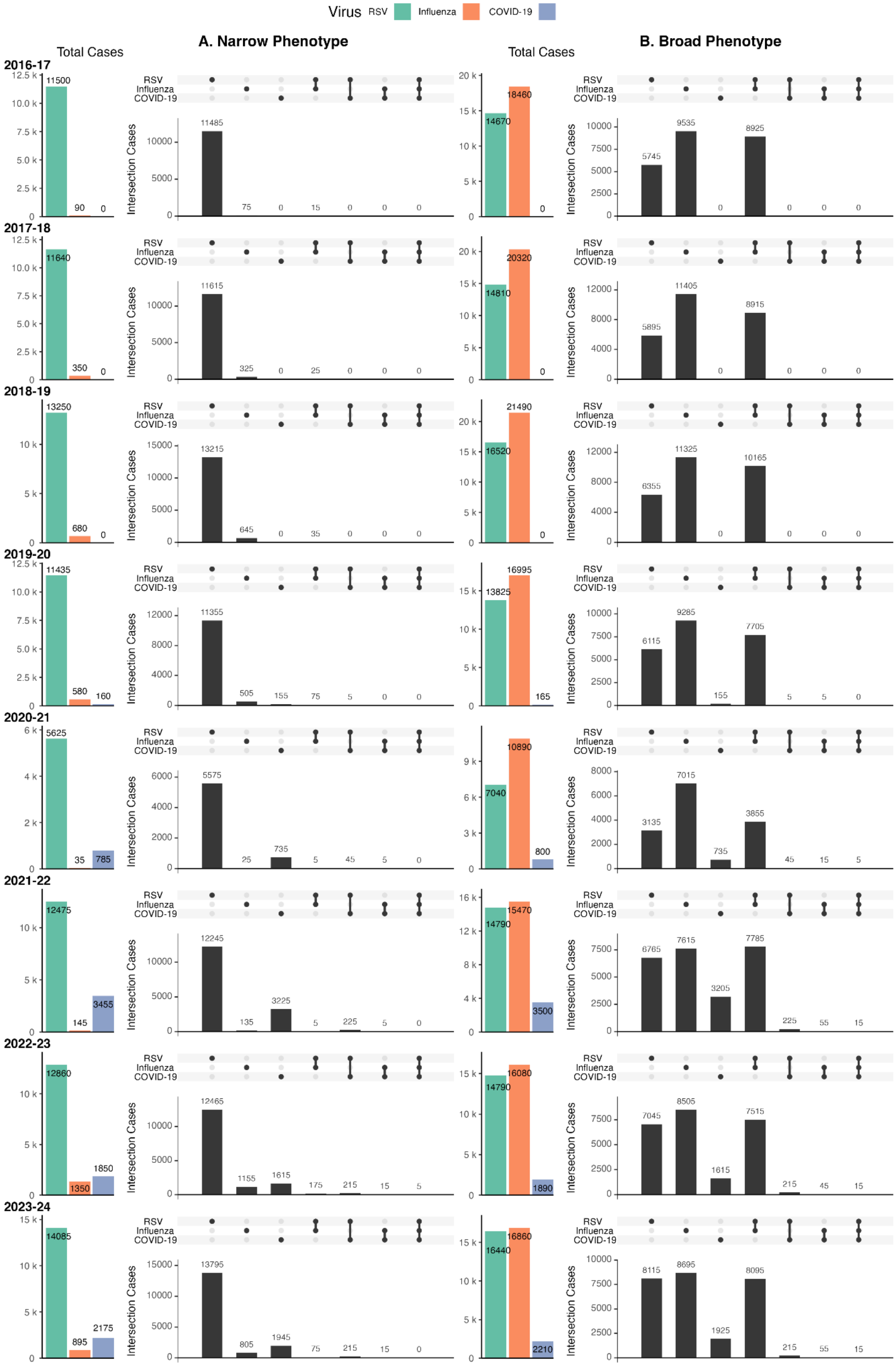

Figure S17: Overlap of severe case identification in infants, comparing narrow phenotypes (A) and broad phenotypes (B). Cases can be RSV (green), influenza (orange), or COVID-19 (purple).

#### S3.5 Maternally-Linked Infants

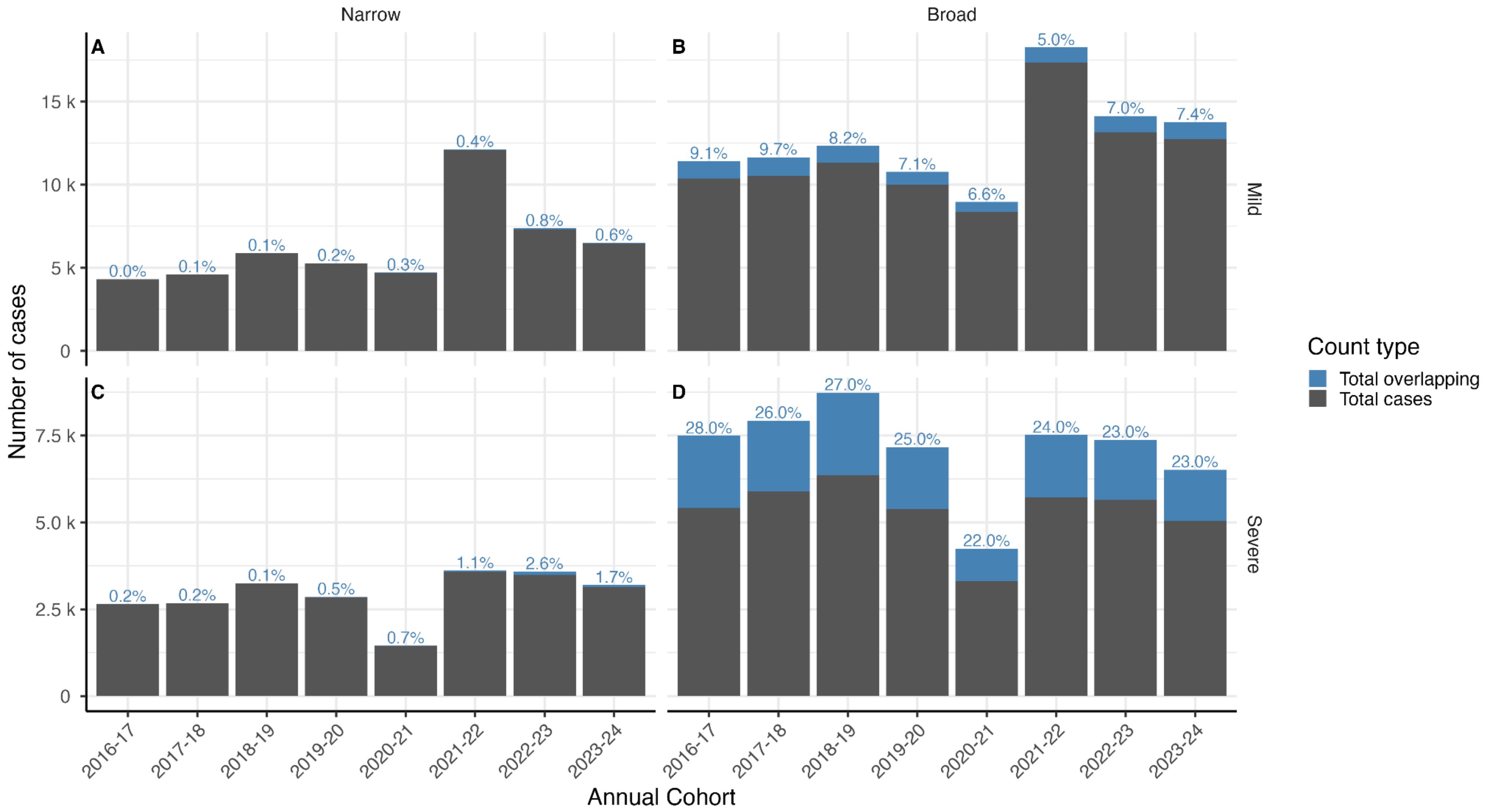

Figure S18: Number of cases in maternally-linked infants which are identified solely as one pathogen (grey) or as multiple pathogens (blue), with percentage of cases overlapping above the bar. Results are shown separately for each case severity, phenotype, and annual cohort.

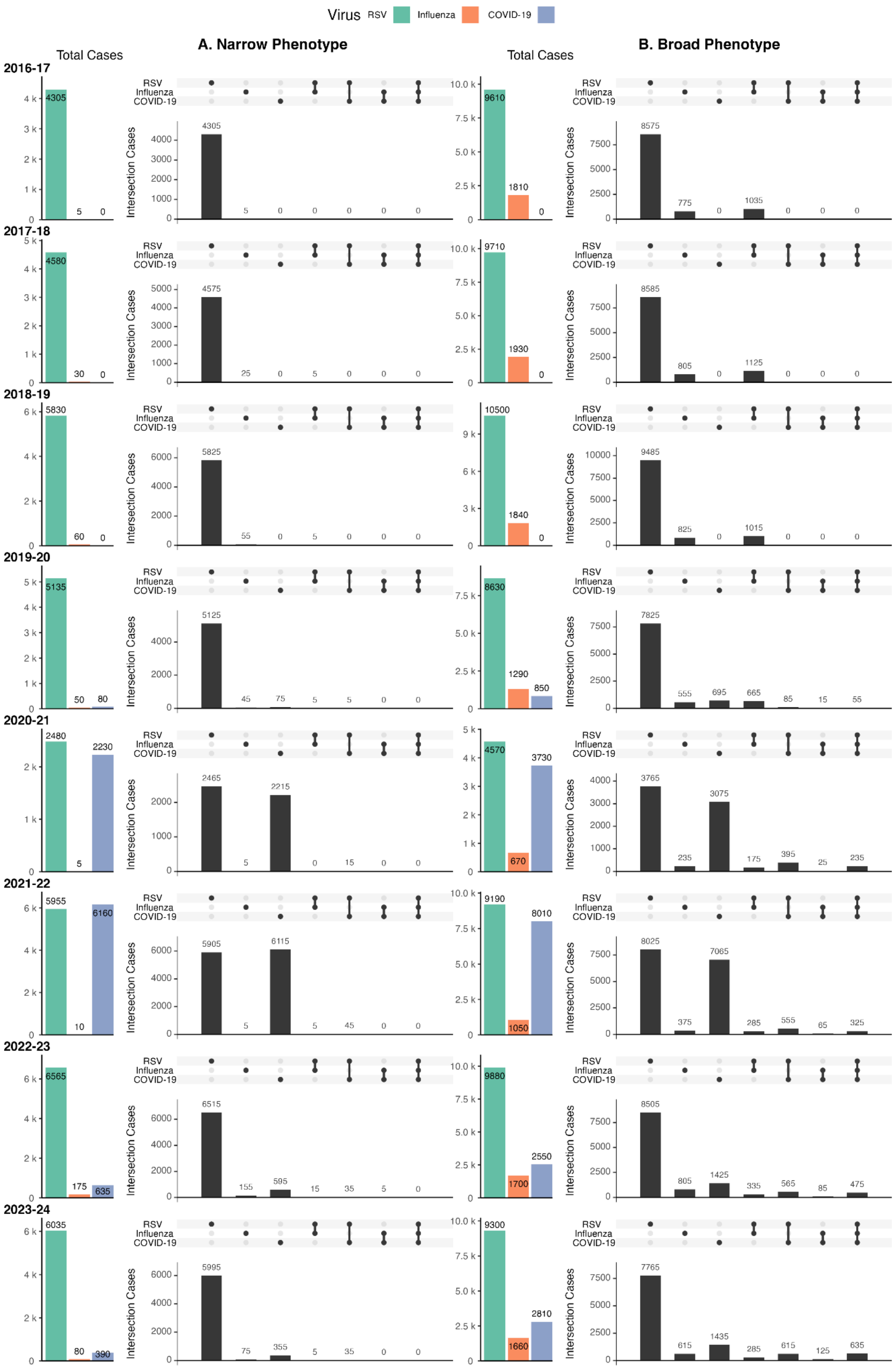

Figure S19: Overlap of mild case identification in maternally-linked infants, comparing narrow phenotypes (A) and broad phenotypes (B). Cases can be RSV (green), influenza (orange), or COVID-19 (purple).

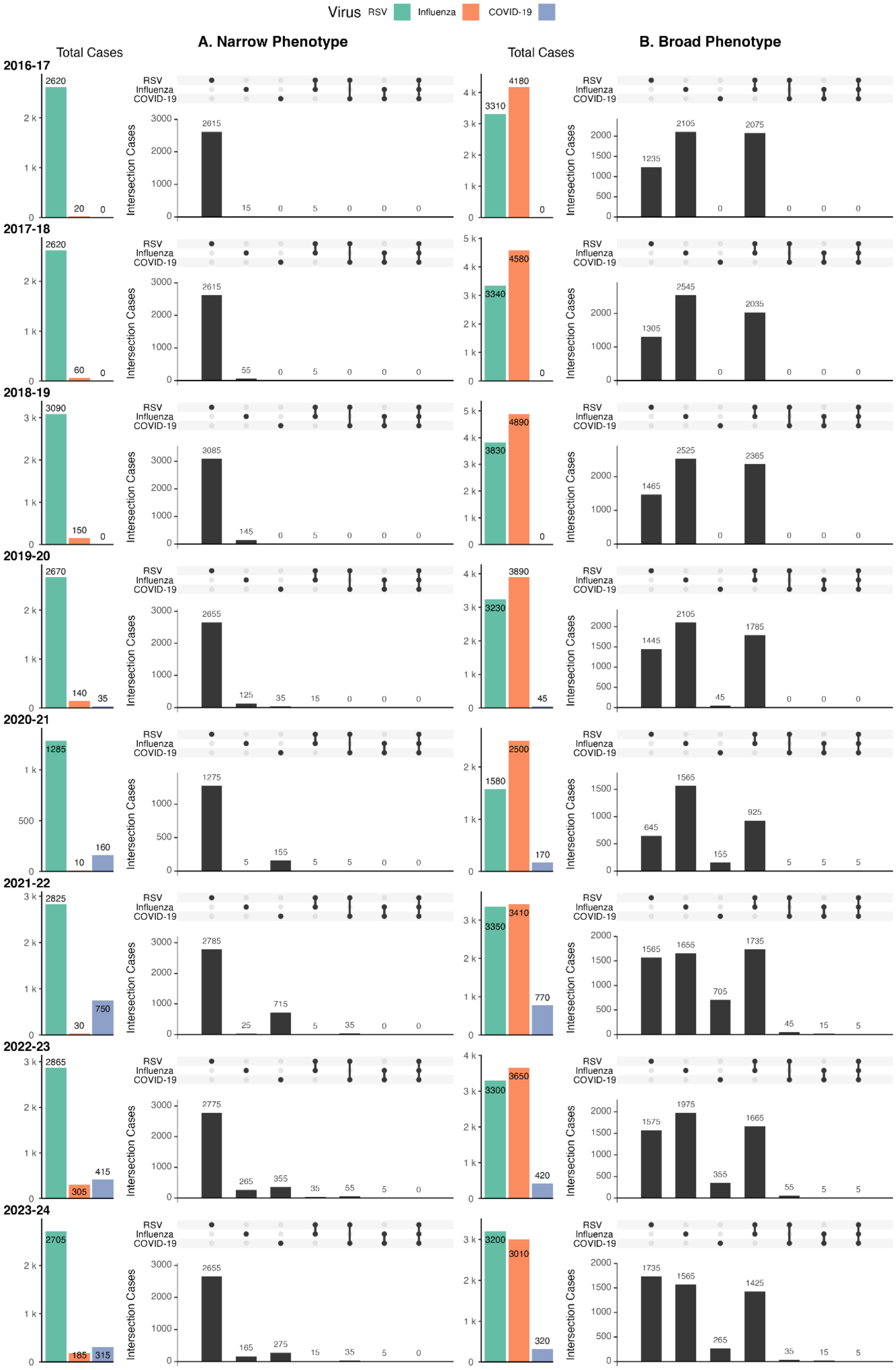

Figure S20: Overlap of severe case identification in maternally-linked infants, comparing narrow phenotypes (A) and broad phenotypes (B). Cases can be RSV (green), influenza (orange), or COVID-19 (purple).

#### S3.6 Case Specification

##### S3.6.1 Phenotype Design Sensitivity Analyses

<table>
<tr><th>Motivation</th><th>Modification</th><th>Difference in results</th></tr>
<tr><td rowspan="4">Sensitivity analysis of COVID-19 window</td><td rowspan="4">For broad phenotypes, the ascertainment window for COVID-19 was increased from 14 to 28 days</td><td>Older Adults</td></tr>
<tr><td><ul><li>Increase in mild case case count from 217,315 to 225,815 (+3.9%)</li><li>Decrease in mild reinfections from 26,325 to 25,695 (-3.4%)</li><li>No change in severe cases (expected)</li><li>Decrease in severe reinfections from 615 to 345 (-43.9%)</li></ul></td></tr>
<tr><td>Infants</td></tr>
<tr><td><ul><li>Increase in mild case count from 14,125 to 14,815 (+4.9%)</li><li>Decrease in mild reinfections from 1,765 to 1,705 (-3.4%)</li><li>No change in severe cases (expected)</li><li>Decrease in severe reinfections from 25 to 15 (-40% )</li></ul></td></tr>
<tr><td rowspan="10">Breakdown of cases by codes: we did this by choosing the codes most used from opencodecounts and re-running our extract without them</td><td rowspan="4">Added antivirals to mild RSV broad</td><td>Older Adults</td></tr>
<tr><td><ul><li>Increase in mild case count from 29,365 to 29,385 (+0.07%)</li><li>No change in mild reinfections</li></ul></td></tr>
<tr><td>Infants</td></tr>
<tr><td><ul><li>No change in mild case count</li><li>No change in mild reinfections</li></ul></td></tr>
<tr><td rowspan="4">Removed ‘cough (finding)’ from mild RSV broad</td><td>Older Adults</td></tr>
<tr><td><ul><li>Decrease in mild case count from 29,365 to 16,995 (-42.1%)</li><li>Decrease in mild reinfections from 1,255 to 595 (-52.6%)</li><li>Decrease in overlapping mild cases from 4,590 to 3,360 (-26.8%)</li></ul></td></tr>
<tr><td>Infants</td></tr>
<tr><td><ul><li>Decrease in mild case count from 49,785 to 43,985 (-11.7%)</li><li>Decrease in mild reinfections from 7,085 to 5,805 (-18.1%)</li><li>Decrease in overlapping mild cases from 15,290 to 8,330 (-45.5%)</li></ul></td></tr>
<tr><td rowspan="2">Removed ‘cough (finding)’ from the ARI codelist used in the mild influenza broad phenotype</td><td>Older Adults</td></tr>
<tr><td><ul><li>Decrease in mild case count from 9,825 to 7,515 (-23.5%)</li><li>Decrease in mild reinfections from 195 to 125 (-35.9%)</li><li>Decrease in overlapping mild cases from 990 to 870 (-12.1%)</li></ul></td></tr>
</table>

| | | |
|---|---|---|
| | | *Infants* |
| | | • Decrease in mild case count from 8,435 to 6,715 (-20.4%)<br>• Decrease in mild reinfections from 525 to 255 (-51.4%)<br>• Increase in overlapping mild cases from 2,080 to 2,170 (4.3%) |
| | Removed 'cough (finding)' from mild COVID-19 broad | *Older Adults* |
| | | • Decrease in mild case count from 217,315 to 203,945 (-6.2%)<br>• Decrease in mild reinfections from 26,325 to 24,135 (-8.3%)<br>• Decrease in overlapping mild cases from 15,590 to 8,730 (-44%) |
| | | *Infants* |
| | | • Decrease in mild case count from 14,125 to 10,175 (-28.0%)<br>• Decrease in mild reinfections from 1,765 to 1,045 (-40.8%)<br>• Decrease in overlapping mild cases from 3,740 to 2,100 (-43.9%) |
| | Removed J069 ('acute upper respiratory infection, unspecified') from severe influenza broad | *Older Adults* |
| | | • Decrease in severe case count from 7,065 to 6,105 (-13.6%)<br>• Decrease in severe reinfections from 85 to 75 (-11.8%) |
| | | *Infants* |
| | | • Decrease in severe case count from 16,865 to 11,265 (-33.2%)<br>• Decrease in severe reinfections from 1,435 to 905 (-36.9%) |
| | Removed J219 ('acute bronchiolitis, unspecified') from severe influenza broad | *Older Adults* |
| | | • Decrease in severe case count from 7,065 to 6,995 (-0.99%)<br>• No change in reinfections |
| | | *Infants* |
| | | • Decrease in severe case count from 16,865 to 9,465 (-43.9%)<br>• Decrease in severe reinfections from 1,435 to 455 (-68.3%) |
| | Removed B972 ('Coronavirus as the cause of diseases classified elsewhere') from severe COVID-19 broad | *Older Adults* |
| | | • Decrease in severe case count from 22,345 to 22,285 (-0.27%)<br>• No change in reinfections |
| | | *Infants* |
| | | • Decrease in severe case count from 2,205 to 2,185 (-0.9%)<br>• No change in reinfections |

Table S13: Sensitivity analysis showing impact of changes in phenotype definitions on case classification for older adults and infants in 2023-24.

#### S3.6.2 Internal Validation of Mild Phenotypes

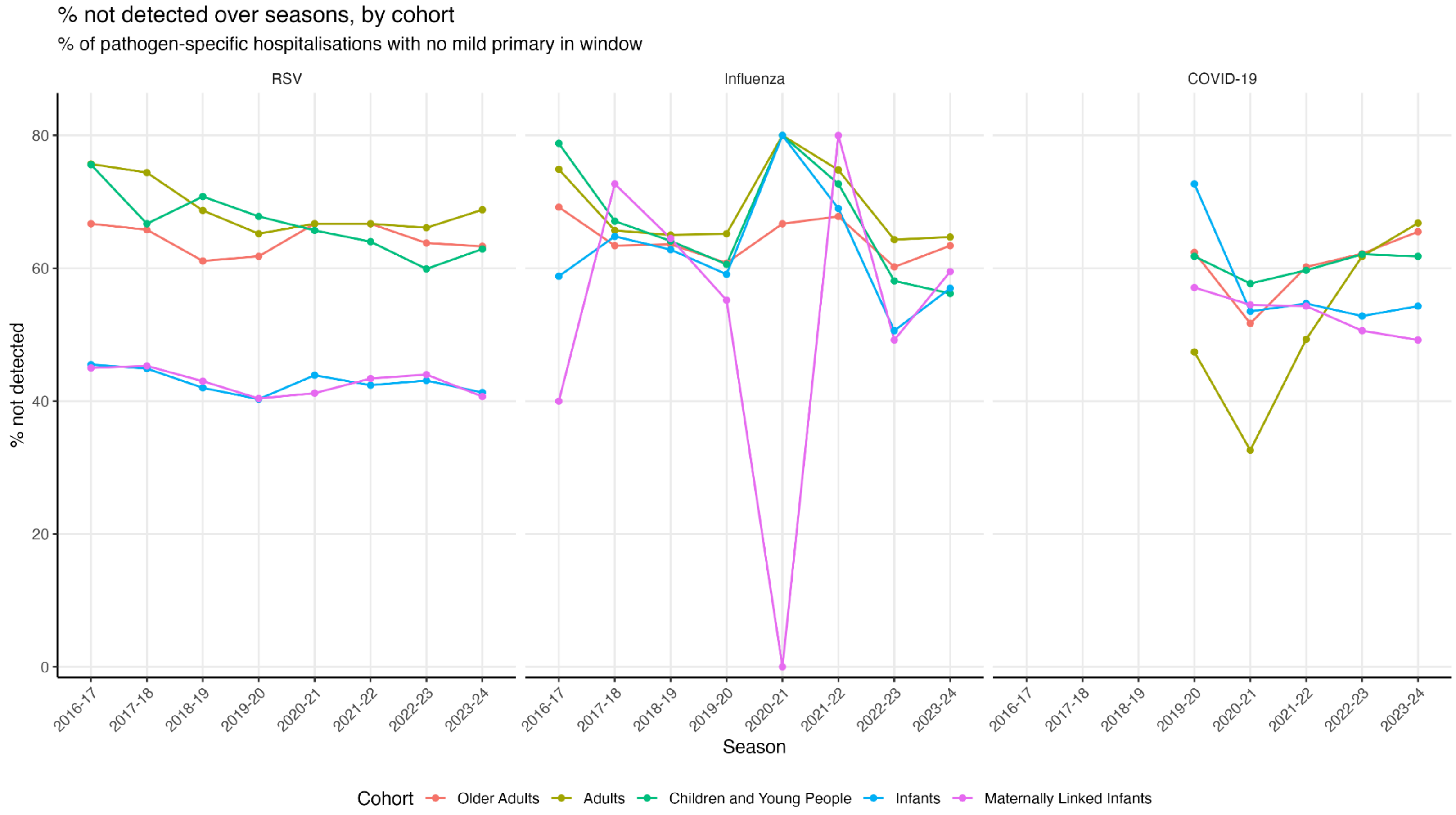

Figure S21: Variation over time of the proportion of severe cases detected as a mild case, within 14 days prior to hospitalisation. Proportions shown over time, grouped by age cohort: red (older adults), khaki (adults), green (children and young people), blue (infants), and pink (maternally-linked infants).

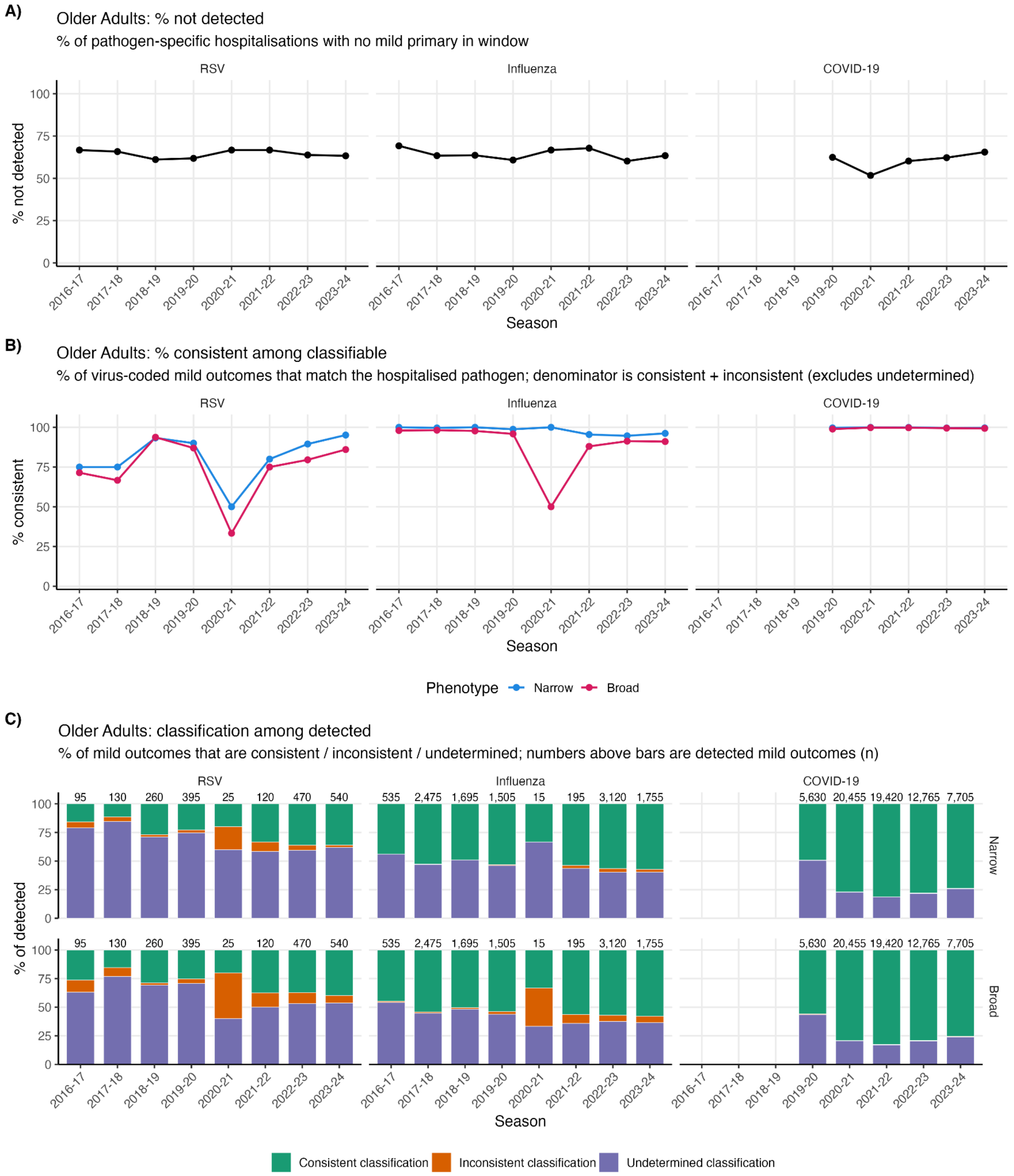

Figure S22: Breakdown of internal validation trends in older adults. A: percentage of severe cases (classified by our narrow phenotypes) which did not have a mild attendance recorded in the 14 days prior to hospitalisation. B: percentage of classifiable mild cases matching the severe reference case. C: percentage of each classification type for mild cases.

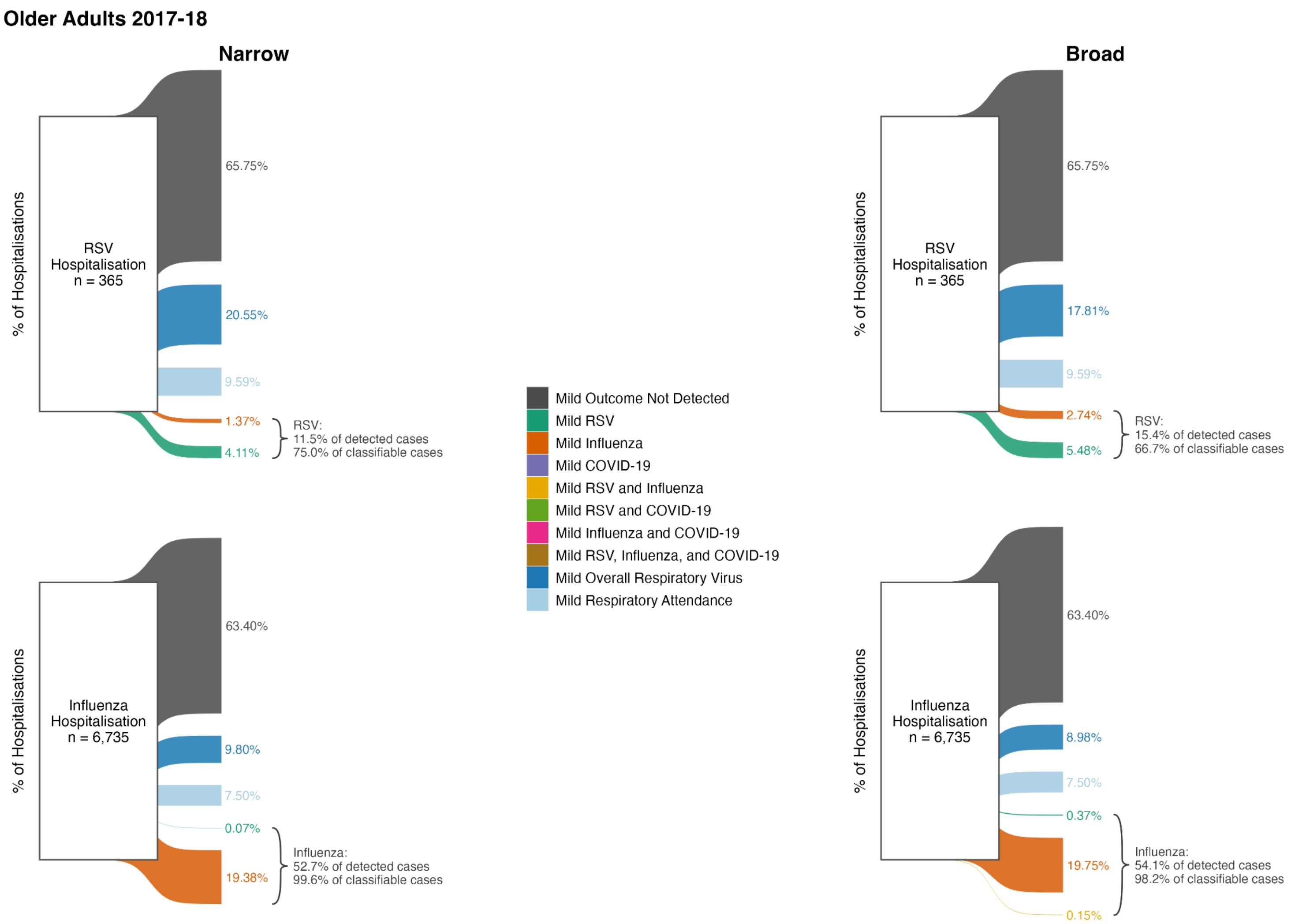

Figure S23: Sankey diagram showing the breakdown of mild presentations within 14 days prior to severe cases for older adults in 2017-18. Reference cases were detected by our severe narrow phenotypes. Curly brackets indicate the percentage of detected mild cases and percentage of classifiable mild cases for which classification was consistent with the severe reference case.

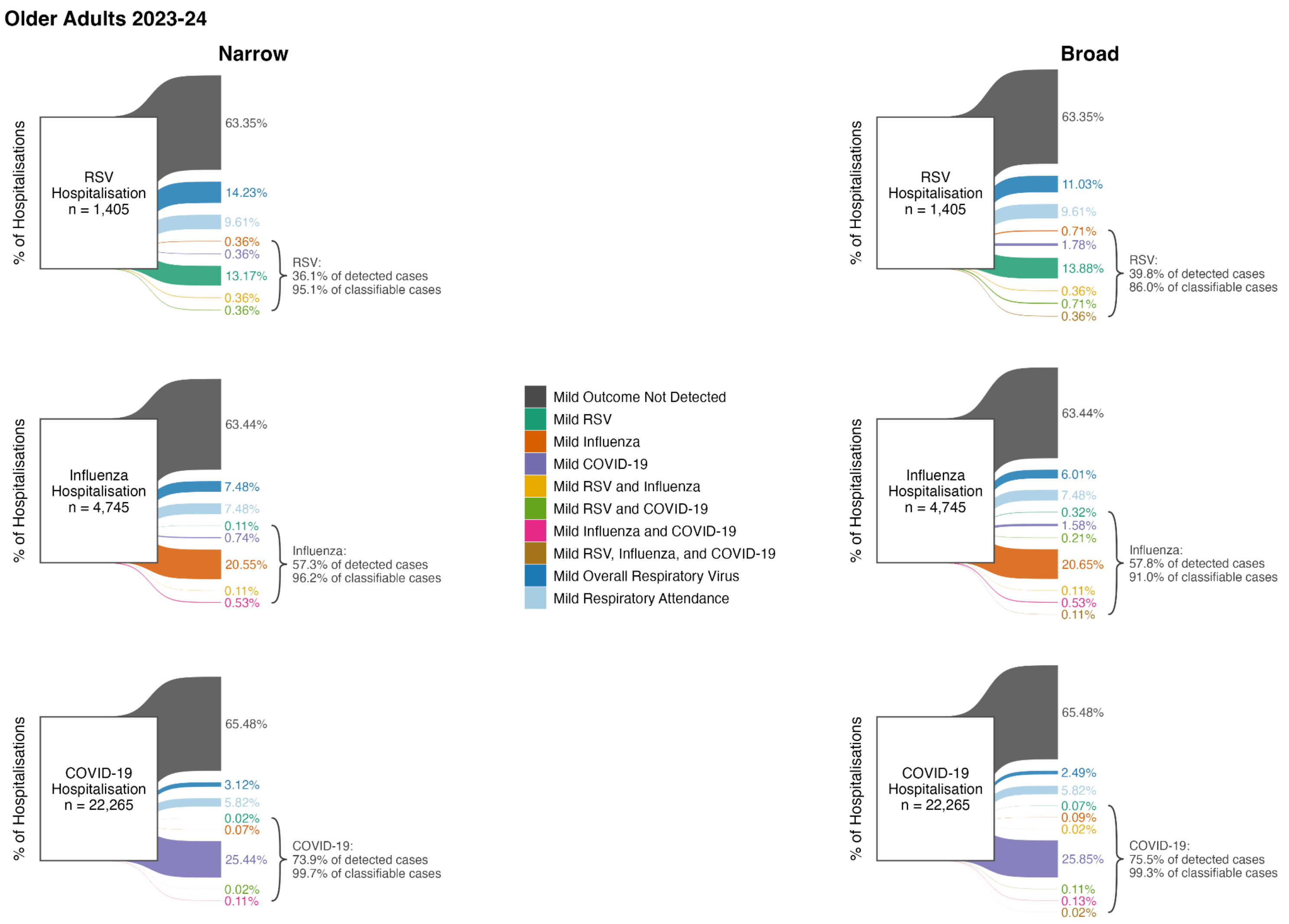

Figure S24: Sankey diagram showing the breakdown of mild presentations within 14 days prior to severe cases for older adults in 2023-24. Reference cases were detected by our severe narrow phenotypes. Curly brackets indicate the percentage of detected mild cases and percentage of classifiable mild cases for which classification was consistent with the severe reference case.

| Older Adults | | | | | | | | | | |
|---|---|---|---|---|---|---|---|---|---|---|
| *Narrow Phenotype (Mild)* | | | | | | | | | | |
| | Annual Cohort | 2016-17 | 2017-18 | 2018-19 | 2019-20 | 2020-21 | 2021-22 | 2022-23 | 2023-24 | Median |
| Severe Case | Mild Case | | | | | | | | | |
| RSV | Not detected | 170 (66.7%) | 240 (65.8%) | 400 (61.1%) | 640 (61.8%) | 50 (66.7%) | 210 (66.7%) | 820 (63.8%) | 890 (63.3%) | 64.80% |
| | Undetermined classification | 75 (29.4%) | 110 (30.1%) | 185 (28.2%) | 295 (28.5%) | 15 (20.0%) | 70 (22.2%) | 280 (21.8%) | 335 (23.8%) | 26.00% |
| | Consistent classification | 15 (75.0%) | 15 (75.0%) | 70 (93.3%) | 90 (90.0%) | 5 (50.0%) | 40 (80.0%) | 170 (89.5%) | 195 (95.1%) | 84.70% |
| | Inconsistent classification | 5 (25.0%) | 5 (25.0%) | 5 (6.7%) | 10 (10.0%) | 5 (50.0%) | 10 (20.0%) | 20 (10.5%) | 10 (4.9%) | 15.30% |
| Influenza | Not detected | 1,180 (69.2%) | 4,270 (63.4%) | 2,950 (63.6%) | 2,320 (60.8%) | 10 (66.7%) | 390 (67.8%) | 4,730 (60.2%) | 3,010 (63.4%) | 63.50% |
| | Undetermined classification | 300 (17.6%) | 1,165 (17.3%) | 865 (18.7%) | 695 (18.2%) | 10 (66.7%) | 85 (14.8%) | 1,255 (16.0%) | 710 (15.0%) | 17.40% |
| | Consistent classification | 235 (100.0%) | 1,305 (99.6%) | 830 (100.0%) | 800 (98.8%) | 5 (100.0%) | 105 (95.5%) | 1,765 (94.6%) | 1,005 (96.2%) | 99.20% |
| | Inconsistent classification | NA | 5 (0.4%) | NA | 10 (1.2%) | NA | 5 (4.5%) | 100 (5.4%) | 40 (3.8%) | 3.80% |
| COVID-19 | Not detected | | | | 9,310 (62.4%) | 21,860 (51.7%) | 29,330 (60.2%) | 21,030 (62.2%) | 14,580 (65.5%) | 62.20% |
| | Undetermined classification | | | | 2,850 (19.1%) | 4,680 (11.1%) | 3,625 (7.4%) | 2,770 (8.2%) | 1,990 (8.9%) | 8.90% |
| | Consistent classification | | | | 2,770 (99.6%) | 15,770 (100.0%) | 15,785 (99.9%) | 9,955 (99.6%) | 5,695 (99.7%) | 99.70% |
| | Inconsistent classification | | | | 10 (0.4%) | 5 (0.0%) | 10 (0.1%) | 40 (0.4%) | 20 (0.3%) | 0.30% |

| *Broad Phenotype (Mild)* | | | | | | | | | | |
|---|---|---|---|---|---|---|---|---|---|---|
| | Annual Cohort | 2016-17 | 2017-18 | 2018-19 | 2019-20 | 2020-21 | 2021-22 | 2022-23 | 2023-24 | Median |
| Severe Case | Mild Case | | | | | | | | | |
| RSV | Not detected | 170 (66.7%) | 240 (65.8%) | 400 (61.1%) | 640 (61.8%) | 50 (66.7%) | 210 (66.7%) | 820 (63.8%) | 890 (63.3%) | 64.80% |
| | Undetermined classification | 60 (23.5%) | 100 (27.4%) | 180 (27.5%) | 280 (27.1%) | 10 (13.3%) | 60 (19.0%) | 250 (19.5%) | 290 (20.6%) | 22.10% |
| | Consistent classification | 25 (71.4%) | 20 (66.7%) | 75 (93.8%) | 100 (87.0%) | 5 (33.3%) | 45 (75.0%) | 175 (79.5%) | 215 (86.0%) | 77.30% |
| | Inconsistent classification | 10 (28.6%) | 10 (33.3%) | 5 (6.2%) | 15 (13.0%) | 10 (66.7%) | 15 (25.0%) | 45 (20.5%) | 35 (14.0%) | 22.70% |
| Influenza | Not detected | 1,180 (69.2%) | 4,270 (63.4%) | 2,950 (63.6%) | 2,320 (60.8%) | 10 (66.7%) | 390 (67.8%) | 4,730 (60.2%) | 3,010 (63.4%) | 63.50% |
| | Undetermined classification | 290 (17.0%) | 1,110 (16.5%) | 820 (17.7%) | 660 (17.3%) | 5 (33.3%) | 70 (12.2%) | 1,170 (14.9%) | 640 (13.5%) | 16.70% |
| | Consistent classification | 240 (98.0%) | 1,340 (98.2%) | 855 (97.7%) | 810 (95.9%) | 5 (50.0%) | 110 (88.0%) | 1,780 (91.3%) | 1,015 (91.0%) | 93.60% |
| | Inconsistent classification | 5 (2.0%) | 25 (1.8%) | 20 (2.3%) | 35 (4.1%) | 5 (50.0%) | 15 (12.0%) | 170 (8.7%) | 100 (9.0%) | 6.40% |
| COVID-19 | Not detected | | | | 9,310 (62.4%) | 21,860 (51.7%) | 29,330 (60.2%) | 21,030 (62.2%) | 14,580 (65.5%) | 62.20% |
| | Undetermined classification | | | | 2,450 (16.4%) | 4,230 (10.0%) | 3,320 (6.8%) | 2,620 (7.8%) | 1,850 (8.3%) | 8.30% |
| | Consistent classification | | | | 3,145 (98.9%) | 16,190 (99.8%) | 16,055 (99.7%) | 10,090 (99.5%) | 5,815 (99.3%) | 99.50% |
| | Inconsistent classification | | | | 35 (1.1%) | 35 (0.2%) | 45 (0.3%) | 55 (0.5%) | 40 (0.7%) | 0.50% |

Table S8: Breakdown of mild presentations within 14 days prior to severe cases for older adults over time. Reference cases were detected by our severe narrow phenotypes. Mild cases were detected via narrow or broad phenotypes. ‘Consistent’ and ‘inconsistent’ classification proportions are for classifiable mild cases.

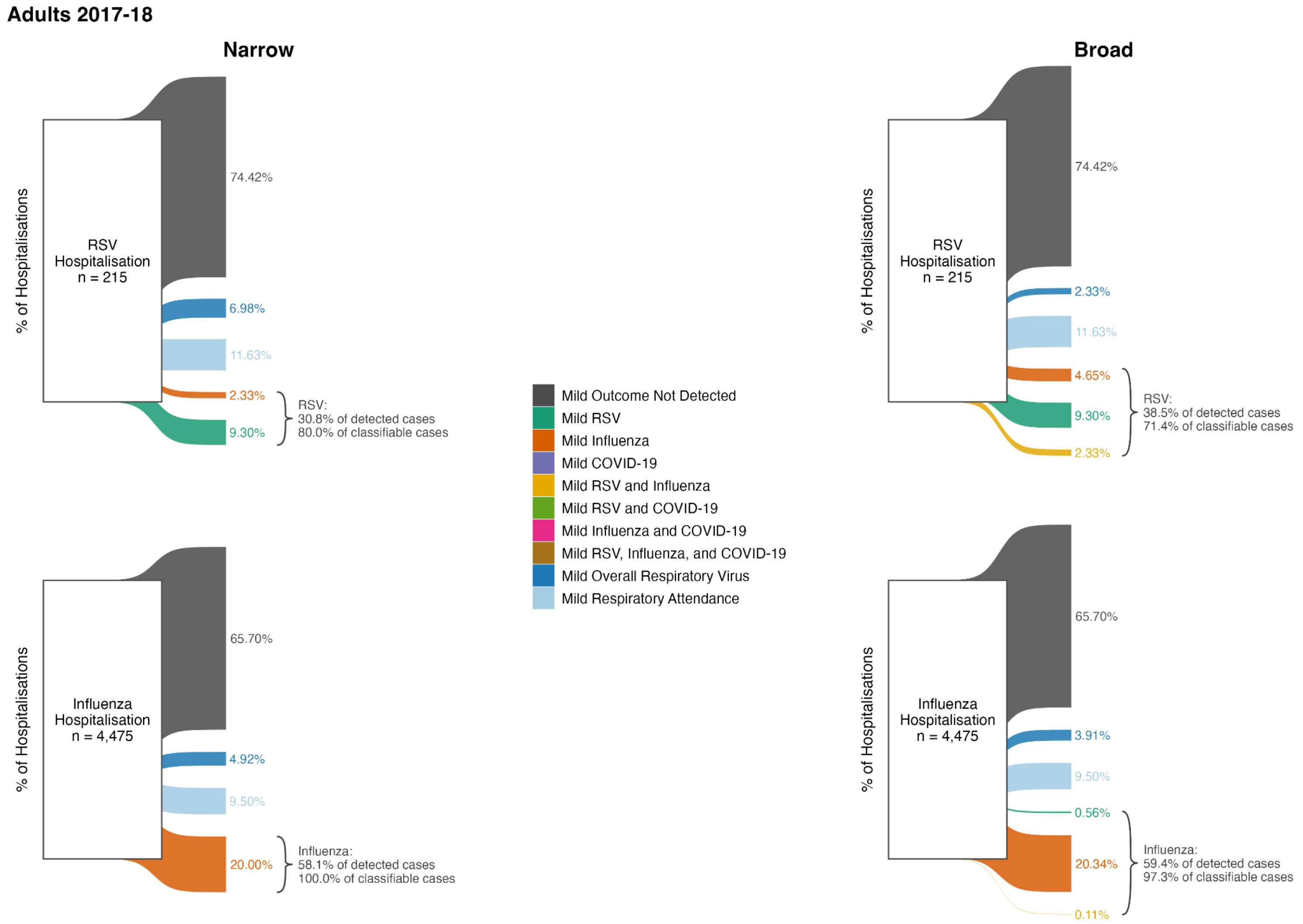

Figure S25: Sankey diagram showing the breakdown of mild presentations within 14 days prior to severe cases for adults in 2017-18. Reference cases were detected by our severe narrow phenotypes. Curly brackets indicate the percentage of detected mild cases and percentage of classifiable mild cases for which classification was consistent with the severe reference case.

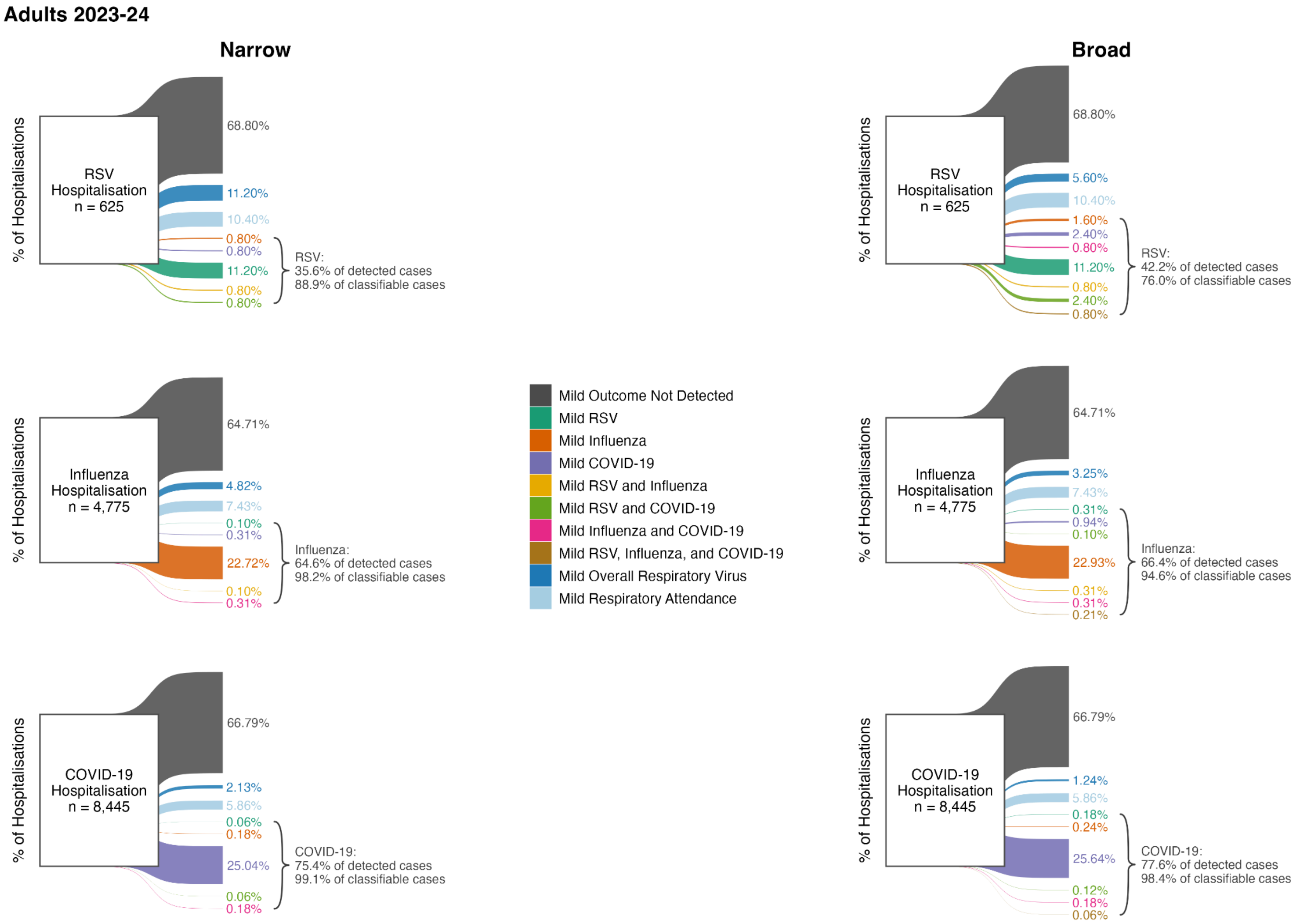

Figure S26: Sankey diagram showing the breakdown of mild presentations within 14 days prior to severe cases for adults in 2023-24. Reference cases were detected by our severe narrow phenotypes. Curly brackets indicate the percentage of detected mild cases and percentage of classifiable mild cases for which classification was consistent with the severe reference case.

| Adults | | | | | | | | | | |
|---|---|---|---|---|---|---|---|---|---|---|
| *Narrow Phenotype (Mild)* | | | | | | | | | | |
| | Annual Cohort | 2016-17 | 2017-18 | 2018-19 | 2019-20 | 2020-21 | 2021-22 | 2022-23 | 2023-24 | Median |
| Severe Case | Mild Case | | | | | | | | | |
| RSV | Not detected | 140 (75.7%) | 160 (74.4%) | 230 (68.7%) | 290 (65.2%) | 50 (66.7%) | 150 (66.7%) | 380 (66.1%) | 430 (68.8%) | 67.70% |
| | Undetermined classification | 35 (18.9%) | 40 (18.6%) | 85 (25.4%) | 100 (22.5%) | 15 (20.0%) | 35 (15.6%) | 120 (20.9%) | 135 (21.6%) | 20.40% |
| | Consistent classification | 15 (100.0%) | 20 (80.0%) | 25 (83.3%) | 65 (92.9%) | 5 (50.0%) | 35 (87.5%) | 95 (90.5%) | 80 (88.9%) | 88.20% |
| | Inconsistent classification | NA | 5 (20.0%) | 5 (16.7%) | 5 (7.1%) | 5 (50.0%) | 5 (12.5%) | 10 (9.5%) | 10 (11.1%) | 12.50% |
| Influenza | Not detected | 850 (74.9%) | 2,940 (65.7%) | 3,620 (65.0%) | 2,160 (65.2%) | 20 (80.0%) | 550 (74.8%) | 3,950 (64.3%) | 3,090 (64.7%) | 65.40% |
| | Undetermined classification | 135 (11.9%) | 645 (14.4%) | 815 (14.6%) | 475 (14.3%) | 10 (40.0%) | 90 (12.2%) | 775 (12.6%) | 585 (12.3%) | 13.50% |
| | Consistent classification | 155 (96.9%) | 895 (100.0%) | 1,145 (99.6%) | 680 (98.6%) | NA | 110 (95.7%) | 1,385 (97.2%) | 1,105 (98.2%) | 98.20% |
| | Inconsistent classification | 5 (3.1%) | NA | 5 (0.4%) | 10 (1.4%) | NA | 5 (4.3%) | 40 (2.8%) | 20 (1.8%) | 2.30% |
| COVID-19 | Not detected | | | | 4,570 (47.4%) | 12,440 (32.6%) | 18,230 (49.3%) | 7,710 (61.8%) | 5,640 (66.8%) | 49.30% |
| | Undetermined classification | | | | 2,560 (26.6%) | 2,860 (7.5%) | 1,930 (5.2%) | 780 (6.3%) | 675 (8.0%) | 7.50% |
| | Consistent classification | | | | 2,515 (99.6%) | 22,820 (100.0%) | 16,860 (100.0%) | 3,985 (99.3%) | 2,135 (99.1%) | 99.60% |
| | Inconsistent classification | | | | 10 (0.4%) | 5 (0.0%) | 5 (0.0%) | 30 (0.7%) | 20 (0.9%) | 0.40% |

| *Broad Phenotype (Mild)* | | | | | | | | | | |
|---|---|---|---|---|---|---|---|---|---|---|
| | Annual Cohort | 2016-17 | 2017-18 | 2018-19 | 2019-20 | 2020-21 | 2021-22 | 2022-23 | 2023-24 | Median |
| Severe Case | Mild Case | | | | | | | | | |
| RSV | Not detected | 140 (75.7%) | 160 (74.4%) | 230 (68.7%) | 290 (65.2%) | 50 (66.7%) | 150 (66.7%) | 380 (66.1%) | 430 (68.8%) | 67.70% |
| | Undetermined classification | 30 (16.2%) | 30 (14.0%) | 70 (20.9%) | 80 (18.0%) | 10 (13.3%) | 30 (13.3%) | 100 (17.4%) | 100 (16.0%) | 16.10% |
| | Consistent classification | 15 (75.0%) | 25 (71.4%) | 35 (77.8%) | 70 (77.8%) | 5 (33.3%) | 35 (77.8%) | 105 (84.0%) | 95 (76.0%) | 76.90% |
| | Inconsistent classification | 5 (25.0%) | 10 (28.6%) | 10 (22.2%) | 20 (22.2%) | 10 (66.7%) | 10 (22.2%) | 20 (16.0%) | 30 (24.0%) | 23.10% |
| Influenza | Not detected | 850 (74.9%) | 2,940 (65.7%) | 3,620 (65.0%) | 2,160 (65.2%) | 20 (80.0%) | 550 (74.8%) | 3,950 (64.3%) | 3,090 (64.7%) | 65.40% |
| | Undetermined classification | 120 (10.6%) | 600 (13.4%) | 750 (13.5%) | 440 (13.3%) | 5 (20.0%) | 60 (8.2%) | 680 (11.1%) | 510 (10.7%) | 12.20% |
| | Consistent classification | 165 (94.3%) | 915 (97.3%) | 1,185 (97.5%) | 695 (95.9%) | NA | 125 (86.2%) | 1,425 (93.8%) | 1,135 (94.6%) | 94.60% |
| | Inconsistent classification | 10 (5.7%) | 25 (2.7%) | 30 (2.5%) | 30 (4.1%) | 5 (100.0%) | 20 (13.8%) | 95 (6.2%) | 65 (5.4%) | 5.60% |
| COVID-19 | Not detected | | | | 4,570 (47.4%) | 12,440 (32.6%) | 18,230 (49.3%) | 7,710 (61.8%) | 5,640 (66.8%) | 49.30% |
| | Undetermined classification | | | | 2,160 (22.4%) | 2,560 (6.7%) | 1,750 (4.7%) | 710 (5.7%) | 600 (7.1%) | 6.70% |
| | Consistent classification | | | | 2,900 (99.1%) | 23,100 (99.9%) | 17,020 (99.9%) | 4,040 (98.9%) | 2,195 (98.4%) | 99.10% |
| | Inconsistent classification | | | | 25 (0.9%) | 25 (0.1%) | 25 (0.1%) | 45 (1.1%) | 35 (1.6%) | 0.90% |

Table S9: Breakdown of mild presentations within 14 days prior to severe cases for adults over time. Reference cases were detected by our severe narrow phenotypes. Mild cases were detected via narrow or broad phenotypes. 'Consistent' and 'inconsistent' classification proportions are for classifiable mild cases.

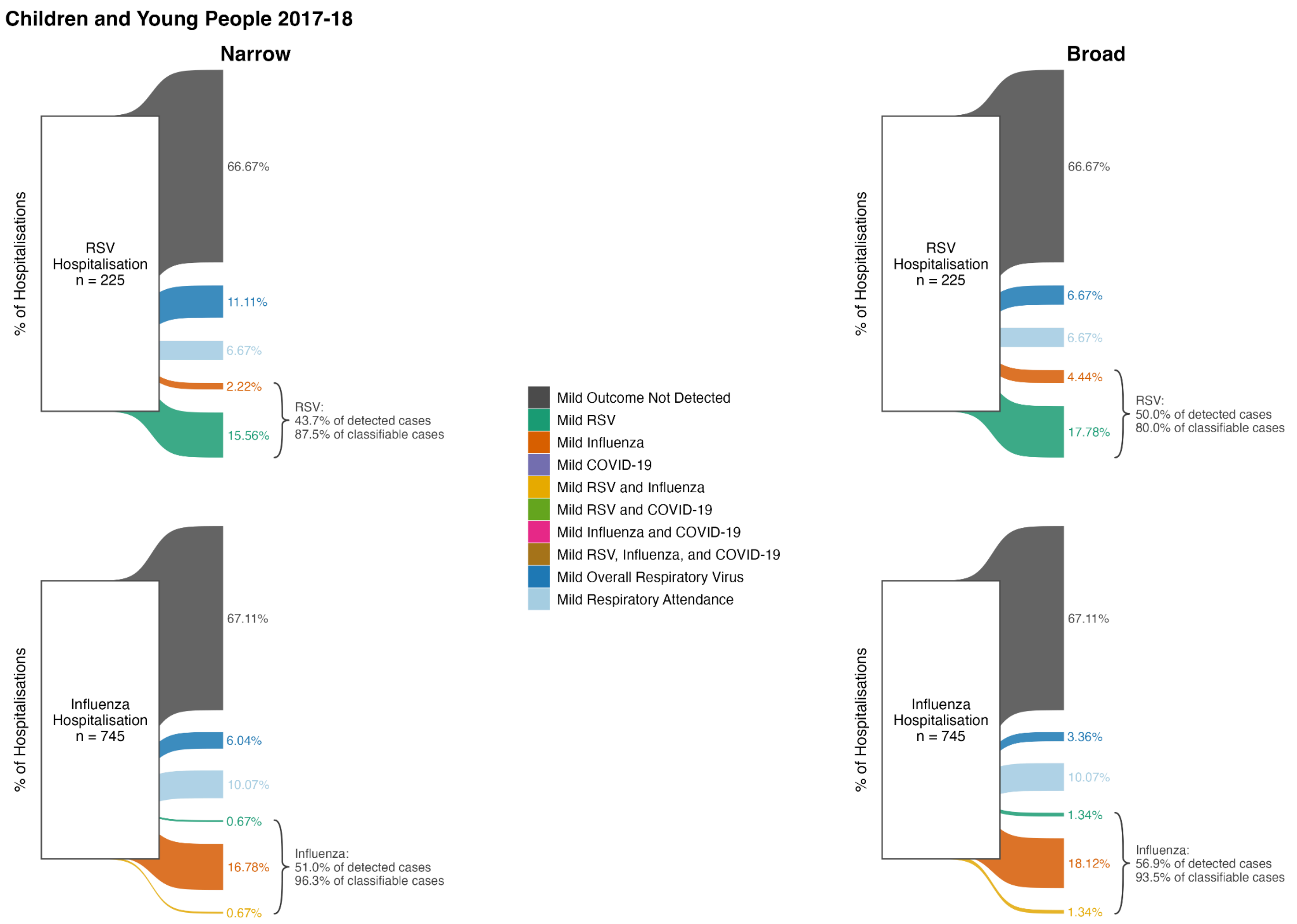

Figure S27: Sankey diagram showing the breakdown of mild presentations within 14 days prior to severe cases for children and young people in 2017-18. Reference cases were detected by our severe narrow phenotypes. Curly brackets indicate the percentage of detected mild cases and percentage of classifiable mild cases for which classification was consistent with the severe reference case.

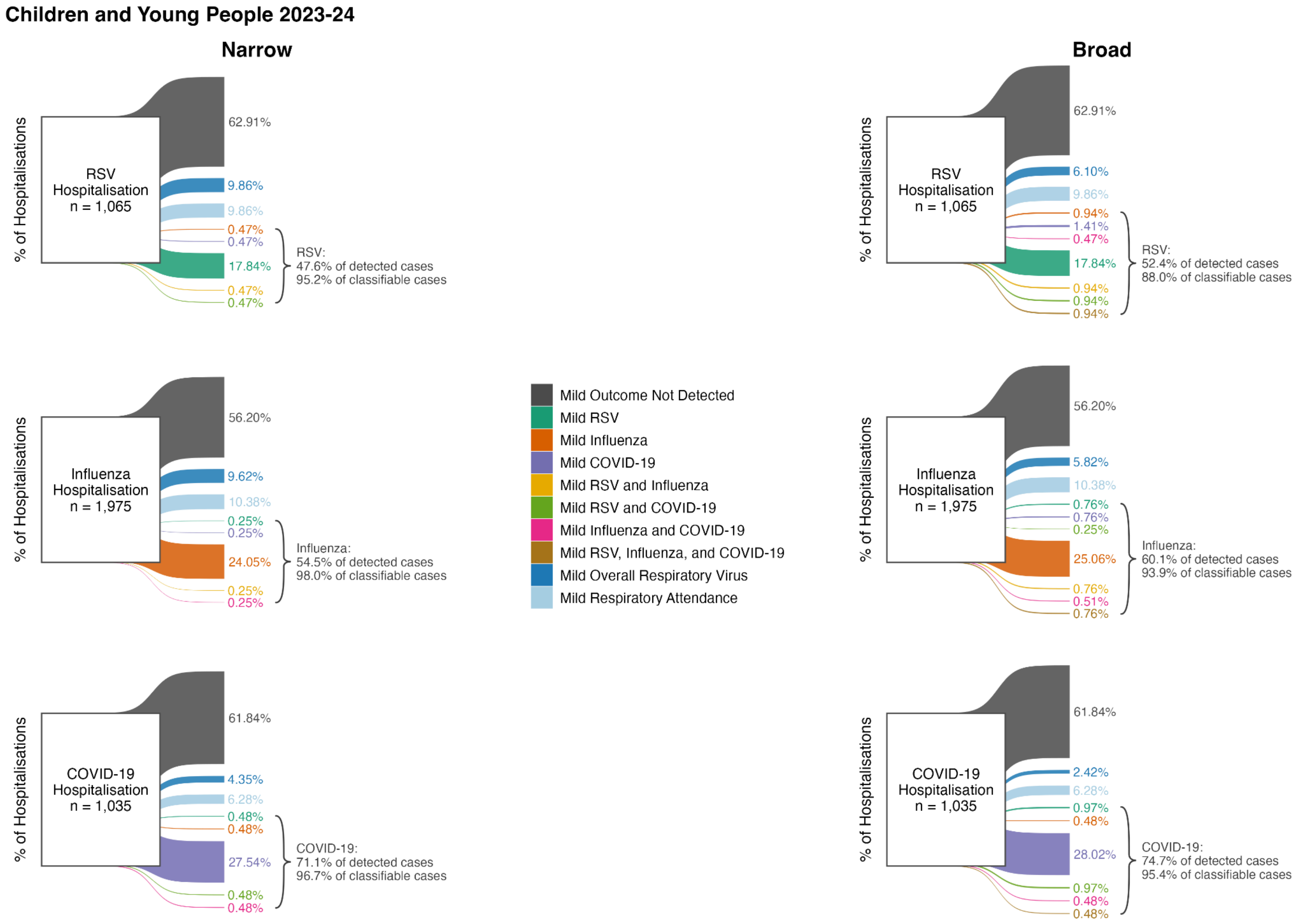

Figure S28: Sankey diagram showing the breakdown of mild presentations within 14 days prior to severe cases for children and young people in 2023-24. Reference cases were detected by our severe narrow phenotypes. Curly brackets indicate the percentage of detected mild cases and percentage of classifiable mild cases for which classification was consistent with the severe reference case.

| Children and Young People | | | | | | | | | | |
|---|---|---|---|---|---|---|---|---|---|---|
| *Narrow Phenotype (Mild)* | | | | | | | | | | |
| | Annual Cohort | 2016-17 | 2017-18 | 2018-19 | 2019-20 | 2020-21 | 2021-22 | 2022-23 | 2023-24 | Median |
| Severe Case | Mild Case | | | | | | | | | |
| RSV | Not detected | 170 (75.6%) | 150 (66.7%) | 230 (70.8%) | 390 (67.8%) | 220 (65.7%) | 560 (64.0%) | 530 (59.9%) | 670 (62.9%) | 66.20% |
| | Undetermined classification | 35 (15.6%) | 40 (17.8%) | 60 (18.5%) | 115 (20.0%) | 75 (22.4%) | 170 (19.4%) | 165 (18.6%) | 210 (19.7%) | 19.00% |
| | Consistent classification | 30 (100.0%) | 35 (87.5%) | 40 (100.0%) | 75 (93.8%) | 70 (100.0%) | 170 (97.1%) | 205 (95.3%) | 200 (95.2%) | 96.20% |
| | Inconsistent classification | NA | 5 (12.5%) | NA | 5 (6.2%) | NA | 5 (2.9%) | 10 (4.7%) | 10 (4.8%) | 4.80% |
| Influenza | Not detected | 130 (78.8%) | 500 (67.1%) | 830 (64.1%) | 760 (60.6%) | 20 (80.0%) | 200 (72.7%) | 1,630 (58.1%) | 1,110 (56.2%) | 65.60% |
| | Undetermined classification | 25 (15.2%) | 120 (16.1%) | 215 (16.6%) | 230 (18.3%) | 5 (20.0%) | 55 (20.0%) | 485 (17.3%) | 395 (20.0%) | 17.80% |
| | Consistent classification | 15 (75.0%) | 130 (96.3%) | 240 (98.0%) | 260 (98.1%) | 5 (50.0%) | 40 (80.0%) | 675 (98.5%) | 485 (98.0%) | 97.10% |
| | Inconsistent classification | 5 (25.0%) | 5 (3.7%) | 5 (2.0%) | 5 (1.9%) | 5 (50.0%) | 10 (20.0%) | 10 (1.5%) | 10 (2.0%) | 2.90% |
| COVID-19 | Not detected | | | | 170 (61.8%) | 770 (57.7%) | 2,350 (59.7%) | 630 (62.1%) | 640 (61.8%) | 61.80% |
| | Undetermined classification | | | | 80 (29.1%) | 110 (8.2%) | 295 (7.5%) | 110 (10.8%) | 110 (10.6%) | 10.60% |
| | Consistent classification | | | | 35 (100.0%) | 465 (98.9%) | 1,325 (99.6%) | 285 (96.6%) | 295 (96.7%) | 98.90% |
| | Inconsistent classification | | | | NA | 5 (1.1%) | 5 (0.4%) | 10 (3.4%) | 10 (3.3%) | 2.20% |

| | | *Broad Phenotype (Mild)* | | | | | | | | |
|---|---|---|---|---|---|---|---|---|---|---|
| | Annual Cohort | 2016-17 | 2017-18 | 2018-19 | 2019-20 | 2020-21 | 2021-22 | 2022-23 | 2023-24 | Median |
| Severe Case | Mild Case | | | | | | | | | |
| RSV | Not detected | 170 (75.6%) | 150 (66.7%) | 230 (70.8%) | 390 (67.8%) | 220 (65.7%) | 560 (64.0%) | 530 (59.9%) | 670 (62.9%) | 66.20% |
| | Undetermined classification | 30 (13.3%) | 30 (13.3%) | 40 (12.3%) | 100 (17.4%) | 50 (14.9%) | 140 (16.0%) | 130 (14.7%) | 170 (16.0%) | 14.80% |
| | Consistent classification | 35 (100.0%) | 40 (80.0%) | 50 (83.3%) | 85 (89.5%) | 90 (94.7%) | 185 (90.2%) | 225 (90.0%) | 220 (88.0%) | 89.70% |
| | Inconsistent classification | NA | 10 (20.0%) | 10 (16.7%) | 10 (10.5%) | 5 (5.3%) | 20 (9.8%) | 25 (10.0%) | 30 (12.0%) | 10.50% |
| Influenza | Not detected | 130 (78.8%) | 500 (67.1%) | 830 (64.1%) | 760 (60.6%) | 20 (80.0%) | 200 (72.7%) | 1,630 (58.1%) | 1,110 (56.2%) | 65.60% |
| | Undetermined classification | 20 (12.1%) | 100 (13.4%) | 190 (14.7%) | 210 (16.7%) | 5 (20.0%) | 40 (14.5%) | 420 (15.0%) | 320 (16.2%) | 14.80% |
| | Consistent classification | 20 (80.0%) | 145 (93.5%) | 255 (94.4%) | 270 (94.7%) | 5 (50.0%) | 50 (76.9%) | 710 (94.7%) | 535 (93.9%) | 93.70% |
| | Inconsistent classification | 5 (20.0%) | 10 (6.5%) | 15 (5.6%) | 15 (5.3%) | 5 (50.0%) | 15 (23.1%) | 40 (5.3%) | 35 (6.1%) | 6.30% |
| COVID-19 | Not detected | | | | 170 (61.8%) | 770 (57.7%) | 2,350 (59.7%) | 630 (62.1%) | 640 (61.8%) | 61.80% |
| | Undetermined classification | | | | 60 (21.8%) | 100 (7.5%) | 240 (6.1%) | 90 (8.9%) | 90 (8.7%) | 8.70% |
| | Consistent classification | | | | 55 (100.0%) | 470 (97.9%) | 1,360 (98.2%) | 300 (95.2%) | 310 (95.4%) | 97.90% |
| | Inconsistent classification | | | | NA | 10 (2.1%) | 25 (1.8%) | 15 (4.8%) | 15 (4.6%) | 3.30% |

Table S10: Breakdown of mild presentations within 14 days prior to severe cases for children and young people over time. Reference cases were detected by our severe narrow phenotypes. Mild cases were detected via narrow or broad phenotypes. ‘Consistent’ and ‘inconsistent’ classification proportions are for classifiable mild cases.

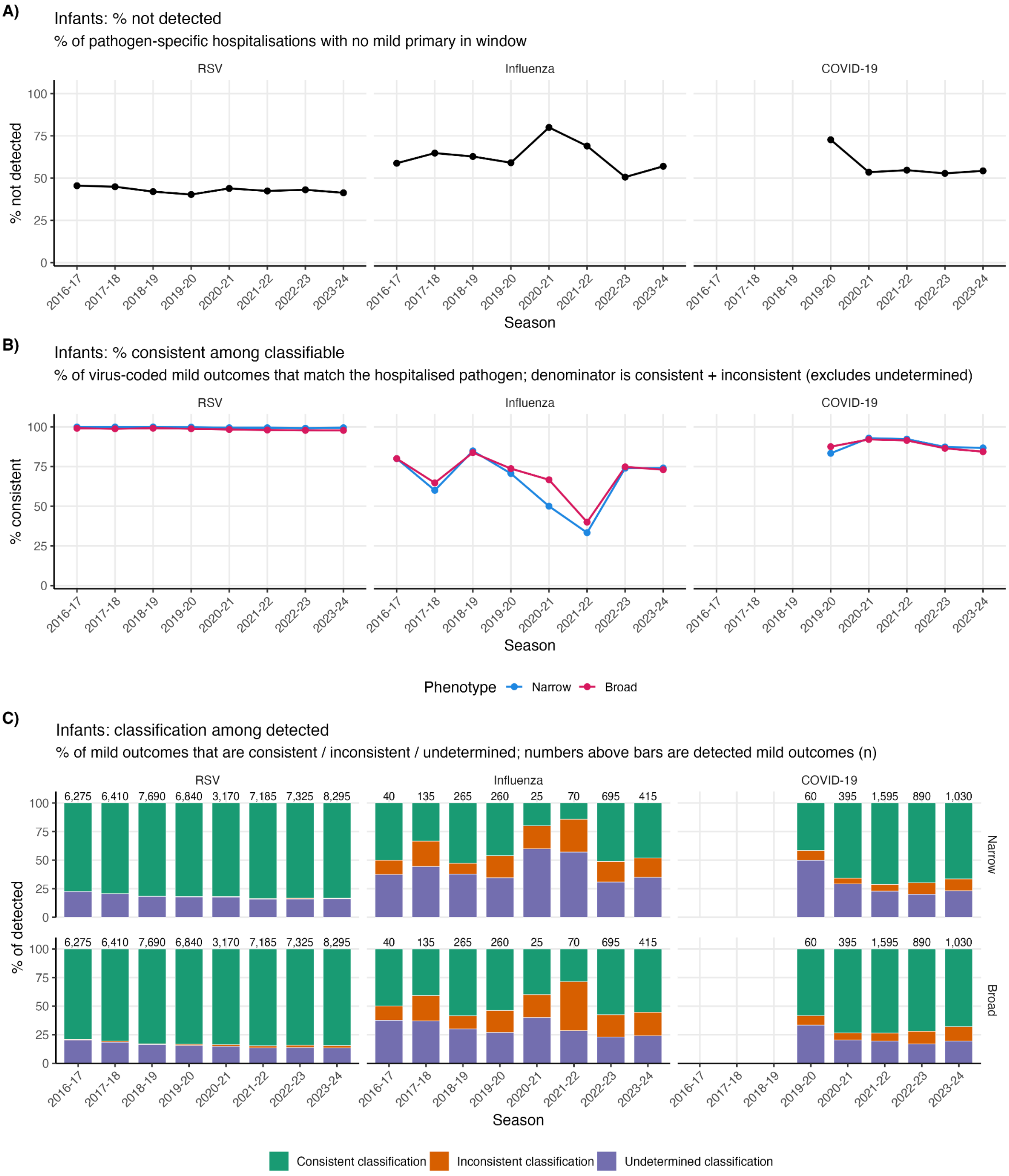

Figure S29: Breakdown of internal validation trends in infants. A: percentage of severe cases (classified by our narrow phenotypes) which did not have a mild attendance recorded in the 14 days prior to hospitalisation. B: percentage of classifiable mild cases matching the severe reference case. C: percentage of each classification type for mild cases.

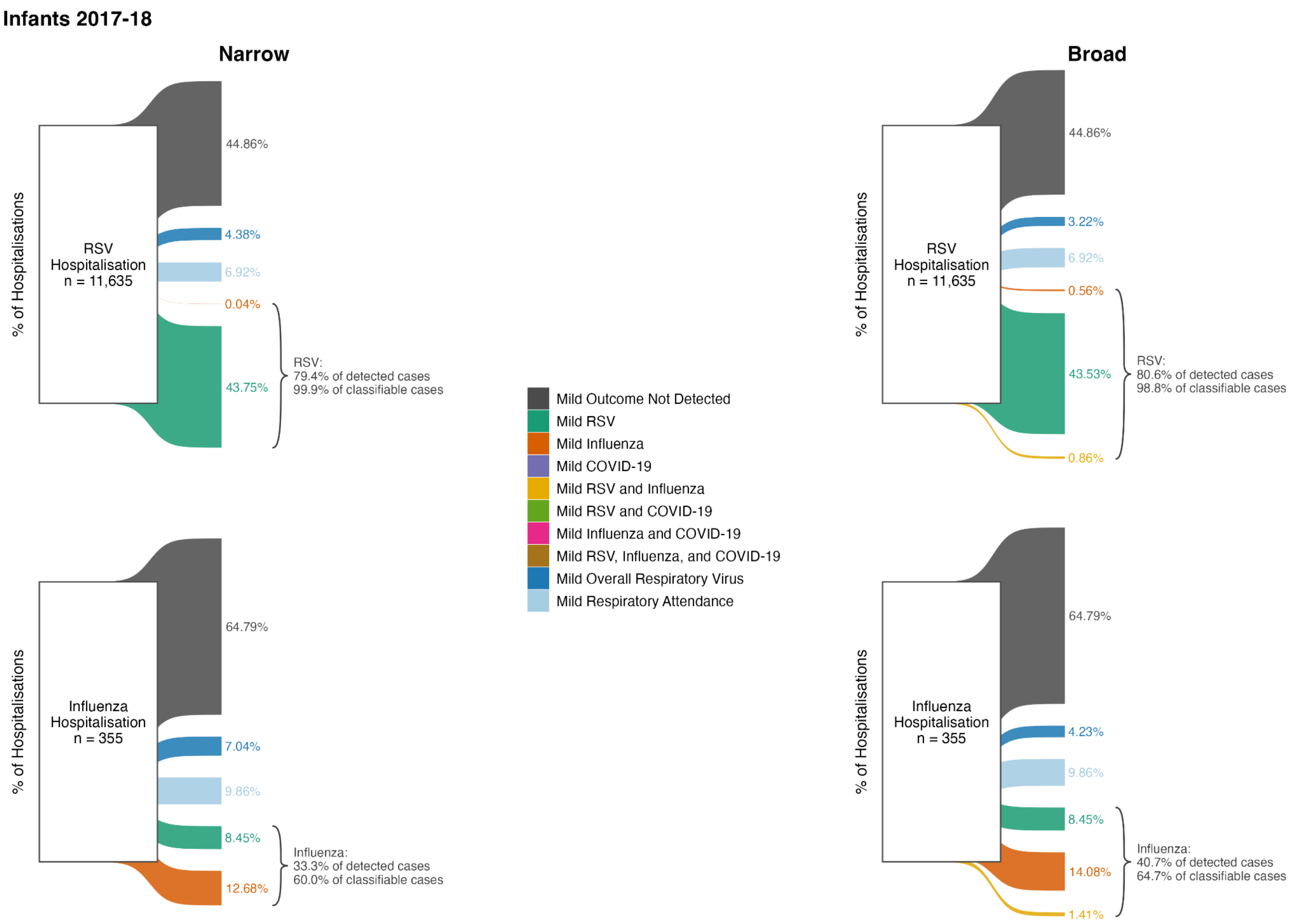

Figure S30: Sankey diagram showing the breakdown of mild presentations within 14 days prior to severe cases for infants in 2017-18. Reference cases were detected by our severe narrow phenotypes. Curly brackets indicate the percentage of detected mild cases and percentage of classifiable mild cases for which classification was consistent with the severe reference case.

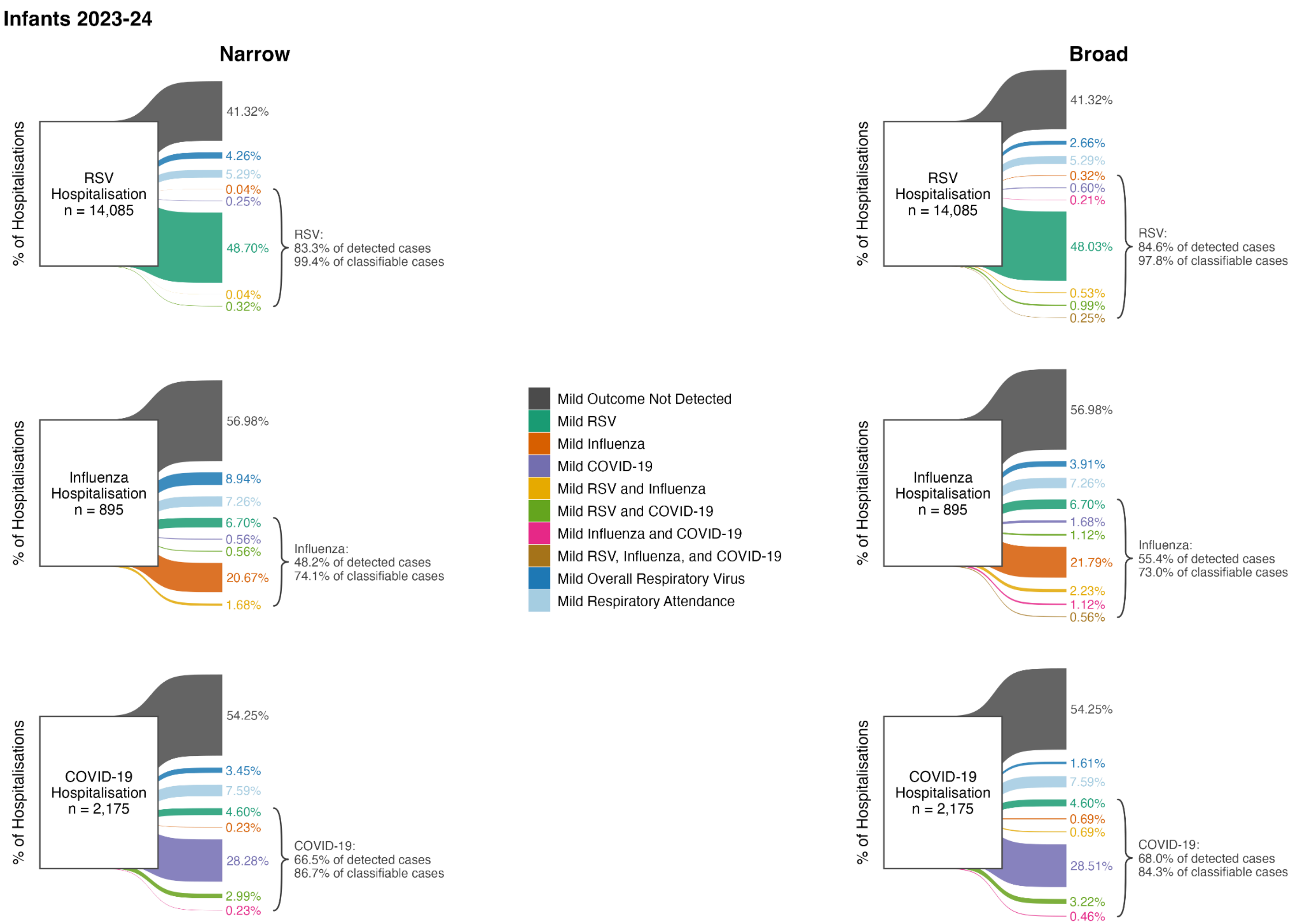

Figure S31: Sankey diagram showing the breakdown of mild presentations within 14 days prior to severe cases for infants in 2023-24. Reference cases were detected by our severe narrow phenotypes. Curly brackets indicate the percentage of detected mild cases and percentage of classifiable mild cases for which classification was consistent with the severe reference case.

| Infants | | | | | | | | | | |
|---|---|---|---|---|---|---|---|---|---|---|
| *Narrow Phenotype (Mild)* | | | | | | | | | | |
| | Annual Cohort | 2016-17 | 2017-18 | 2018-19 | 2019-20 | 2020-21 | 2021-22 | 2022-23 | 2023-24 | Median |
| Severe Case | Mild Case | | | | | | | | | |
| RSV | Not detected | 5,230 (45.5%) | 5,220 (44.9%) | 5,560 (42.0%) | 4,610 (40.3%) | 2,470 (43.9%) | 5,290 (42.4%) | 5,540 (43.1%) | 5,820 (41.3%) | 42.80% |
| | Undetermined classification | 1,410 (12.3%) | 1,315 (11.3%) | 1,410 (10.6%) | 1,225 (10.7%) | 560 (10.0%) | 1,145 (9.2%) | 1,170 (9.1%) | 1,345 (9.5%) | 10.30% |
| | Consistent classification | 4,860 (99.9%) | 5,090 (99.9%) | 6,275 (99.9%) | 5,605 (99.8%) | 2,595 (99.4%) | 6,005 (99.4%) | 6,100 (99.1%) | 6,910 (99.4%) | 99.60% |
| | Inconsistent classification | 5 (0.1%) | 5 (0.1%) | 5 (0.1%) | 10 (0.2%) | 15 (0.6%) | 35 (0.6%) | 55 (0.9%) | 40 (0.6%) | 0.40% |
| Influenza | Not detected | 50 (58.8%) | 230 (64.8%) | 430 (62.8%) | 340 (59.1%) | 20 (80.0%) | 100 (69.0%) | 680 (50.6%) | 510 (57.0%) | 61.00% |
| | Undetermined classification | 15 (17.6%) | 60 (16.9%) | 100 (14.6%) | 90 (15.7%) | 15 (60.0%) | 40 (27.6%) | 215 (16.0%) | 145 (16.2%) | 16.60% |
| | Consistent classification | 20 (80.0%) | 45 (60.0%) | 140 (84.8%) | 120 (70.6%) | 5 (50.0%) | 10 (33.3%) | 355 (74.0%) | 200 (74.1%) | 72.30% |
| | Inconsistent classification | 5 (20.0%) | 30 (40.0%) | 25 (15.2%) | 50 (29.4%) | 5 (50.0%) | 20 (66.7%) | 125 (26.0%) | 70 (25.9%) | 27.70% |
| COVID-19 | Not detected | | | | 120 (72.7%) | 420 (53.5%) | 1,890 (54.7%) | 980 (52.8%) | 1,180 (54.3%) | 54.30% |
| | Undetermined classification | | | | 30 (18.2%) | 115 (14.6%) | 365 (10.6%) | 180 (9.7%) | 240 (11.0%) | 11.00% |
| | Consistent classification | | | | 25 (83.3%) | 260 (92.9%) | 1,135 (92.3%) | 620 (87.3%) | 685 (86.7%) | 87.30% |
| | Inconsistent classification | | | | 5 (16.7%) | 20 (7.1%) | 95 (7.7%) | 90 (12.7%) | 105 (13.3%) | 12.70% |

| *Broad Phenotype (Mild)* | | | | | | | | | | |
|---|---|---|---|---|---|---|---|---|---|---|
| | Annual Cohort | 2016-17 | 2017-18 | 2018-19 | 2019-20 | 2020-21 | 2021-22 | 2022-23 | 2023-24 | Median |
| Severe Case | Mild Case | | | | | | | | | |
| RSV | Not detected | 5,230 (45.5%) | 5,220 (44.9%) | 5,560 (42.0%) | 4,610 (40.3%) | 2,470 (43.9%) | 5,290 (42.4%) | 5,540 (43.1%) | 5,820 (41.3%) | 42.80% |
| | Undetermined classification | 1,270 (11.0%) | 1,180 (10.1%) | 1,260 (9.5%) | 1,070 (9.4%) | 470 (8.4%) | 970 (7.8%) | 1,010 (7.9%) | 1,120 (8.0%) | 8.90% |
| | Consistent classification | 4,960 (99.1%) | 5,165 (98.8%) | 6,375 (99.1%) | 5,700 (98.8%) | 2,655 (98.3%) | 6,090 (98.0%) | 6,175 (97.8%) | 7,015 (97.8%) | 98.50% |
| | Inconsistent classification | 45 (0.9%) | 65 (1.2%) | 55 (0.9%) | 70 (1.2%) | 45 (1.7%) | 125 (2.0%) | 140 (2.2%) | 160 (2.2%) | 1.50% |
| Influenza | Not detected | 50 (58.8%) | 230 (64.8%) | 430 (62.8%) | 340 (59.1%) | 20 (80.0%) | 100 (69.0%) | 680 (50.6%) | 510 (57.0%) | 61.00% |
| | Undetermined classification | 15 (17.6%) | 50 (14.1%) | 80 (11.7%) | 70 (12.2%) | 10 (40.0%) | 20 (13.8%) | 160 (11.9%) | 100 (11.2%) | 13.00% |
| | Consistent classification | 20 (80.0%) | 55 (64.7%) | 155 (83.8%) | 140 (73.7%) | 10 (66.7%) | 20 (40.0%) | 400 (74.8%) | 230 (73.0%) | 73.40% |
| | Inconsistent classification | 5 (20.0%) | 30 (35.3%) | 30 (16.2%) | 50 (26.3%) | 5 (33.3%) | 30 (60.0%) | 135 (25.2%) | 85 (27.0%) | 26.60% |
| COVID-19 | Not detected | | | | 120 (72.7%) | 420 (53.5%) | 1,890 (54.7%) | 980 (52.8%) | 1,180 (54.3%) | 54.30% |
| | Undetermined classification | | | | 20 (12.1%) | 80 (10.2%) | 310 (9.0%) | 150 (8.1%) | 200 (9.2%) | 9.20% |
| | Consistent classification | | | | 35 (87.5%) | 290 (92.1%) | 1,175 (91.4%) | 640 (86.5%) | 700 (84.3%) | 87.50% |
| | Inconsistent classification | | | | 5 (12.5%) | 25 (7.9%) | 110 (8.6%) | 100 (13.5%) | 130 (15.7%) | 12.50% |

Table S11: Breakdown of mild presentations within 14 days prior to severe cases for infants over time. Reference cases were detected by our severe narrow phenotypes. Mild cases were detected via narrow or broad phenotypes. ‘Consistent’ and ‘inconsistent’ classification proportions are for classifiable mild cases.

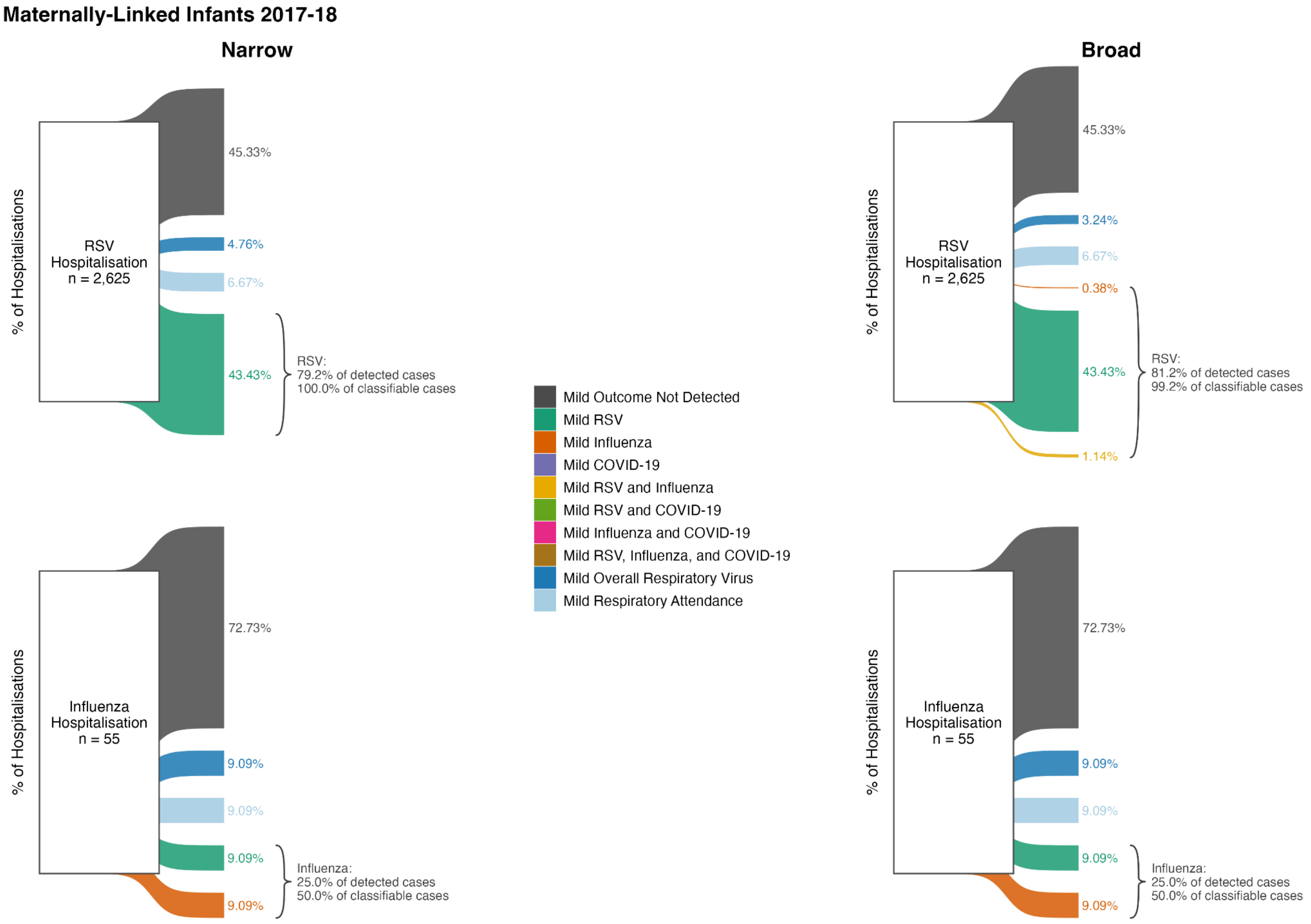

Figure S32: Sankey diagram showing the breakdown of mild presentations within 14 days prior to severe cases for maternally-linked infants in 2017-18. Reference cases were detected by our severe narrow phenotypes. Curly brackets indicate the percentage of detected mild cases and percentage of classifiable mild cases for which classification was consistent with the severe reference case.

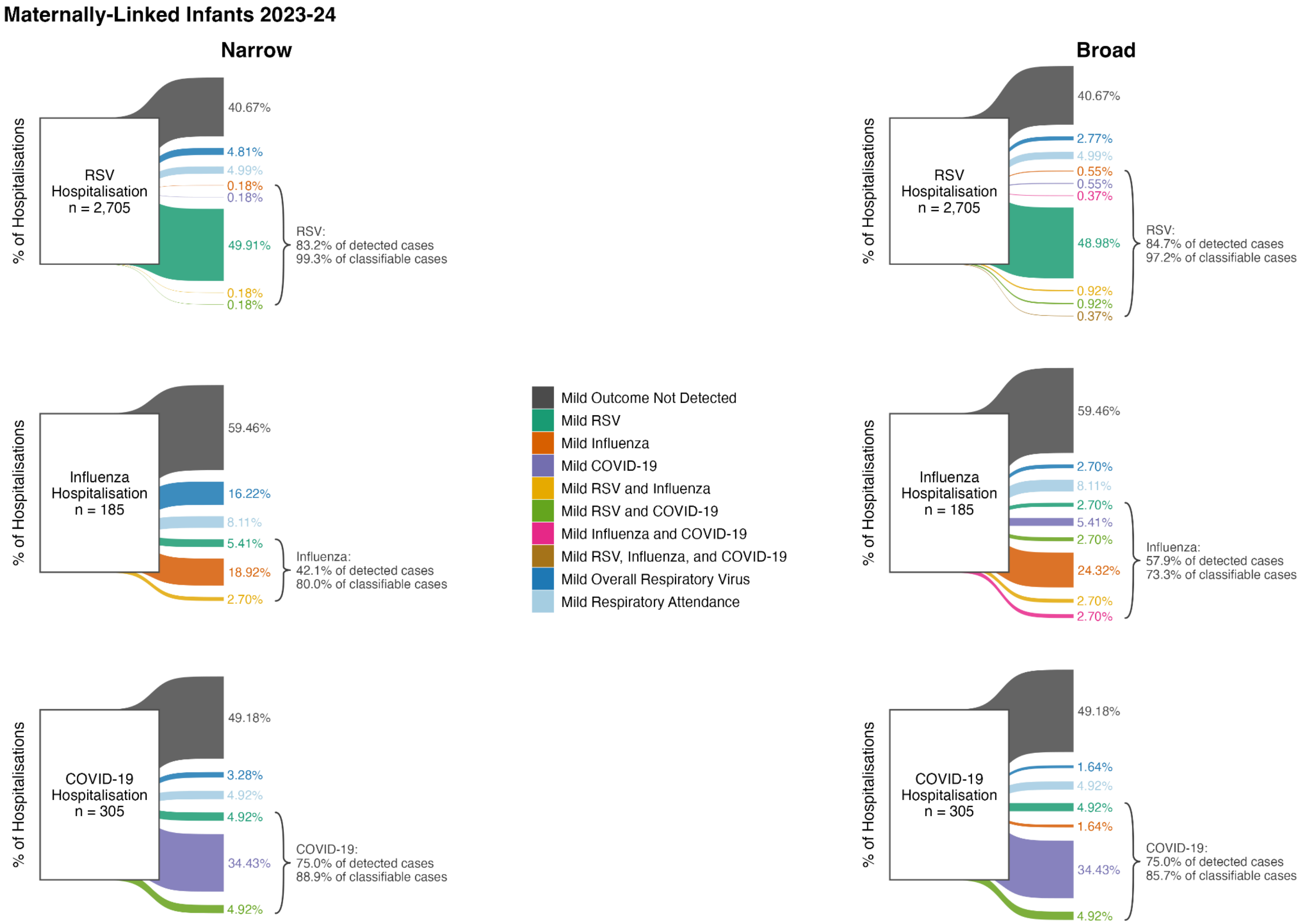

Figure S33: Sankey diagram showing the breakdown of mild presentations within 14 days prior to severe cases for maternally-linked infants in 2023-24. Reference cases were detected by our severe narrow phenotypes. Curly brackets indicate the percentage of detected mild cases and percentage of classifiable mild cases for which classification was consistent with the severe reference case.

| Maternally-Linked Infants | | | | | | | | | | |
|---|---|---|---|---|---|---|---|---|---|---|
| *Narrow Phenotype (Mild)* | | | | | | | | | | |
| | Annual Cohort | 2016-17 | 2017-18 | 2018-19 | 2019-20 | 2020-21 | 2021-22 | 2022-23 | 2023-24 | Median |
| Severe Case | Mild Case | | | | | | | | | |
| RSV | Not detected | 1,180 (45.0%) | 1,190 (45.3%) | 1,330 (43.0%) | 1,080 (40.4%) | 530 (41.2%) | 1,230 (43.4%) | 1,260 (44.0%) | 1,100 (40.7%) | 43.20% |
| | Undetermined classification | 325 (12.4%) | 300 (11.4%) | 330 (10.7%) | 300 (11.2%) | 125 (9.7%) | 260 (9.2%) | 285 (9.9%) | 265 (9.8%) | 10.30% |
| | Consistent classification | 1,120 (99.6%) | 1,140 (100.0%) | 1,440 (99.7%) | 1,315 (99.6%) | 635 (99.2%) | 1,345 (99.6%) | 1,340 (99.3%) | 1,360 (99.3%) | 99.60% |
| | Inconsistent classification | 5 (0.4%) | NA | 5 (0.3%) | 5 (0.4%) | 5 (0.8%) | 5 (0.4%) | 10 (0.7%) | 10 (0.7%) | 0.40% |
| Influenza | Not detected | 10 (40.0%) | 40 (72.7%) | 100 (64.5%) | 80 (55.2%) | 0 (0.0%) | 20 (80.0%) | 150 (49.2%) | 110 (59.5%) | 57.30% |
| | Undetermined classification | 5 (20.0%) | 10 (18.2%) | 15 (9.7%) | 25 (17.2%) | 10 (200.0%) | 10 (40.0%) | 55 (18.0%) | 45 (24.3%) | 19.10% |
| | Consistent classification | 5 (50.0%) | 5 (50.0%) | 40 (88.9%) | 30 (60.0%) | NA | 5 (50.0%) | 90 (78.3%) | 40 (80.0%) | 60.00% |
| | Inconsistent classification | 5 (50.0%) | 5 (50.0%) | 5 (11.1%) | 20 (40.0%) | NA | 5 (50.0%) | 25 (21.7%) | 10 (20.0%) | 40.00% |
| COVID-19 | Not detected | | | | 20 (57.1%) | 90 (54.5%) | 410 (54.3%) | 210 (50.6%) | 150 (49.2%) | 54.30% |
| | Undetermined classification | | | | 15 (42.9%) | 35 (21.2%) | 75 (9.9%) | 50 (12.0%) | 25 (8.2%) | 12.00% |
| | Consistent classification | | | | 5 (100.0%) | 65 (92.9%) | 260 (96.3%) | 145 (85.3%) | 120 (88.9%) | 92.90% |
| | Inconsistent classification | | | | NA | 5 (7.1%) | 10 (3.7%) | 25 (14.7%) | 15 (11.1%) | 9.10% |

| *Broad Phenotype (Mild)* | | | | | | | | | | |
|---|---|---|---|---|---|---|---|---|---|---|
| | Annual Cohort | 2016-17 | 2017-18 | 2018-19 | 2019-20 | 2020-21 | 2021-22 | 2022-23 | 2023-24 | Median |
| Severe Case | Mild Case | | | | | | | | | |
| RSV | Not detected | 1,180 (45.0%) | 1,190 (45.3%) | 1,330 (43.0%) | 1,080 (40.4%) | 530 (41.2%) | 1,230 (43.4%) | 1,260 (44.0%) | 1,100 (40.7%) | 43.20% |
| | Undetermined classification | 290 (11.0%) | 260 (9.9%) | 280 (9.0%) | 250 (9.3%) | 110 (8.6%) | 210 (7.4%) | 230 (8.0%) | 210 (7.8%) | 8.80% |
| | Consistent classification | 1,140 (98.3%) | 1,170 (99.2%) | 1,480 (99.0%) | 1,340 (97.8%) | 645 (98.5%) | 1,370 (97.9%) | 1,365 (97.2%) | 1,385 (97.2%) | 98.10% |
| | Inconsistent classification | 20 (1.7%) | 10 (0.8%) | 15 (1.0%) | 30 (2.2%) | 10 (1.5%) | 30 (2.1%) | 40 (2.8%) | 40 (2.8%) | 1.90% |
| Influenza | Not detected | 10 (40.0%) | 40 (72.7%) | 100 (64.5%) | 80 (55.2%) | 0 (0.0%) | 20 (80.0%) | 150 (49.2%) | 110 (59.5%) | 57.30% |
| | Undetermined classification | 5 (20.0%) | 10 (18.2%) | 10 (6.5%) | 20 (13.8%) | 5 (100.0%) | 10 (40.0%) | 30 (9.8%) | 20 (10.8%) | 16.00% |
| | Consistent classification | 5 (50.0%) | 5 (50.0%) | 45 (90.0%) | 35 (63.6%) | 5 (100.0%) | 5 (50.0%) | 105 (75.0%) | 55 (73.3%) | 68.50% |
| | Inconsistent classification | 5 (50.0%) | 5 (50.0%) | 5 (10.0%) | 20 (36.4%) | NA | 5 (50.0%) | 35 (25.0%) | 20 (26.7%) | 36.40% |
| COVID-19 | Not detected | | | | 20 (57.1%) | 90 (54.5%) | 410 (54.3%) | 210 (50.6%) | 150 (49.2%) | 54.30% |
| | Undetermined classification | | | | 10 (28.6%) | 20 (12.1%) | 60 (7.9%) | 30 (7.2%) | 20 (6.6%) | 7.90% |
| | Consistent classification | | | | 10 (100.0%) | 75 (88.2%) | 270 (94.7%) | 160 (84.2%) | 120 (85.7%) | 88.20% |
| | Inconsistent classification | | | | NA | 10 (11.8%) | 15 (5.3%) | 30 (15.8%) | 20 (14.3%) | 13.00% |

Table S12: Breakdown of mild presentations within 14 days prior to severe cases for maternally-linked infants over time. Reference cases were detected by our severe narrow phenotypes. Mild cases were detected via narrow or broad phenotypes. ‘Consistent’ and ‘inconsistent’ classification proportions are for classifiable mild cases.

##### S3.6.3 Reinfections

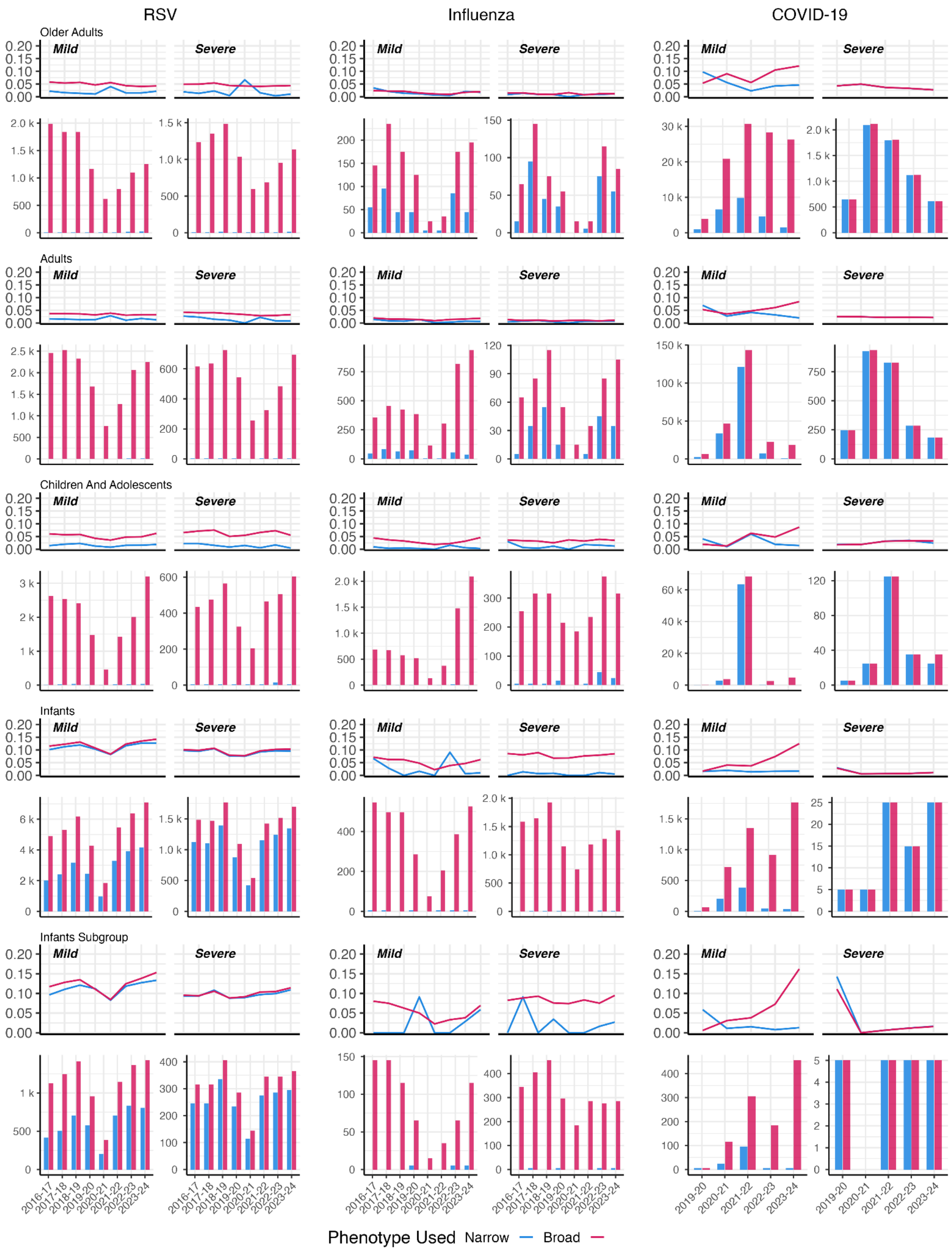

Figure S34: Proportion (lines) and number (bars) of individuals with an initial infection who had a subsequent second infection of the same virus within the same annual cohort. Proportions/counts are shown separately by case severity, age cohort, and annual cohort, with narrow and broad cases on the same panel indicated by blue (narrow) and pink (broad) lines/bars.

*Reinfections Within 28 Days*

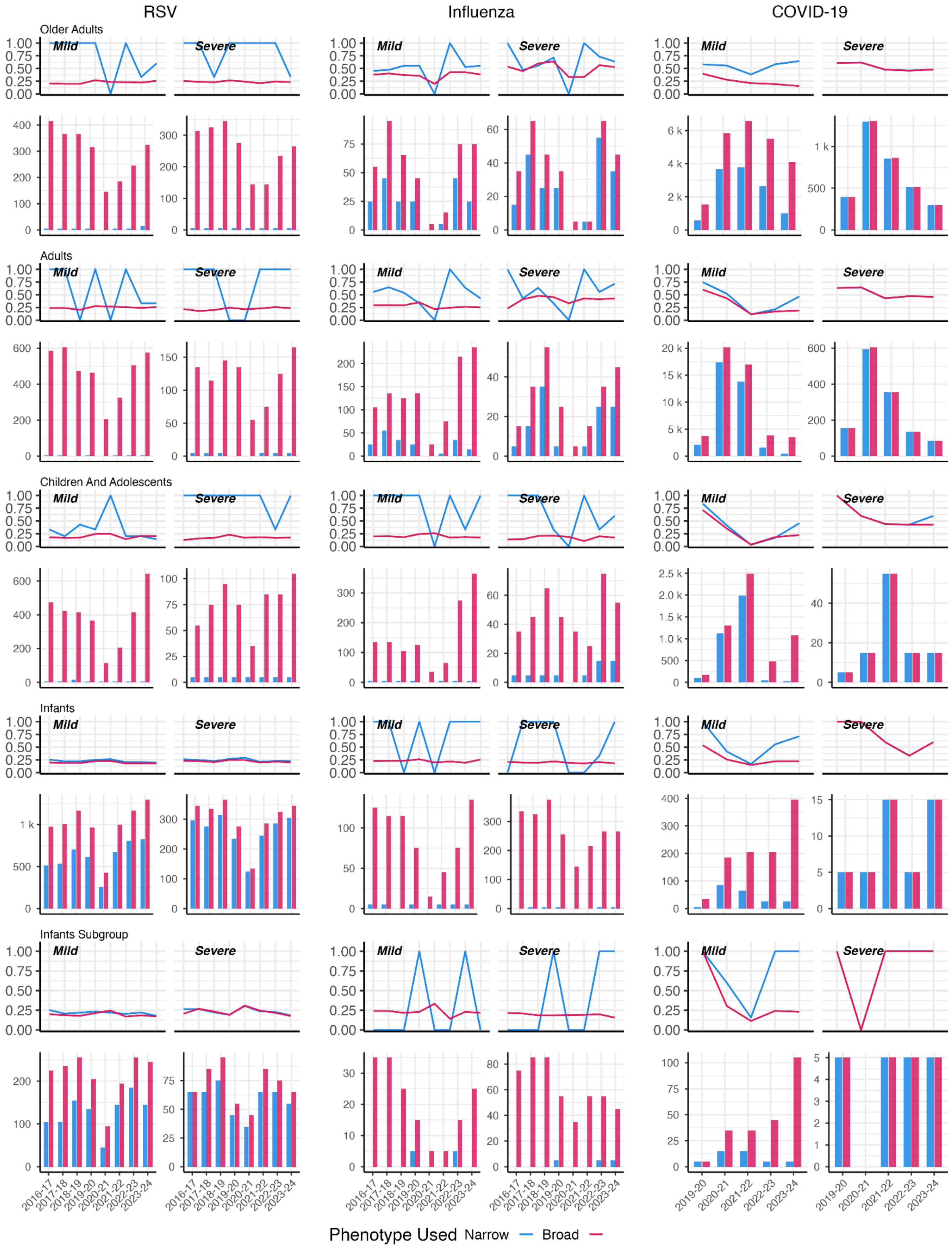

Figure S35: Proportion (lines) and number (bars) of individuals with a reinfection occurring within 28 days of their first infection of the same virus within the same annual cohort. Proportions/counts are shown separately by case severity, age cohort, and annual cohort, with narrow and broad cases on the same panel indicated by blue (narrow) and pink (broad) lines/bars.

#### S3.6.4 Overall Respiratory Virus Cases

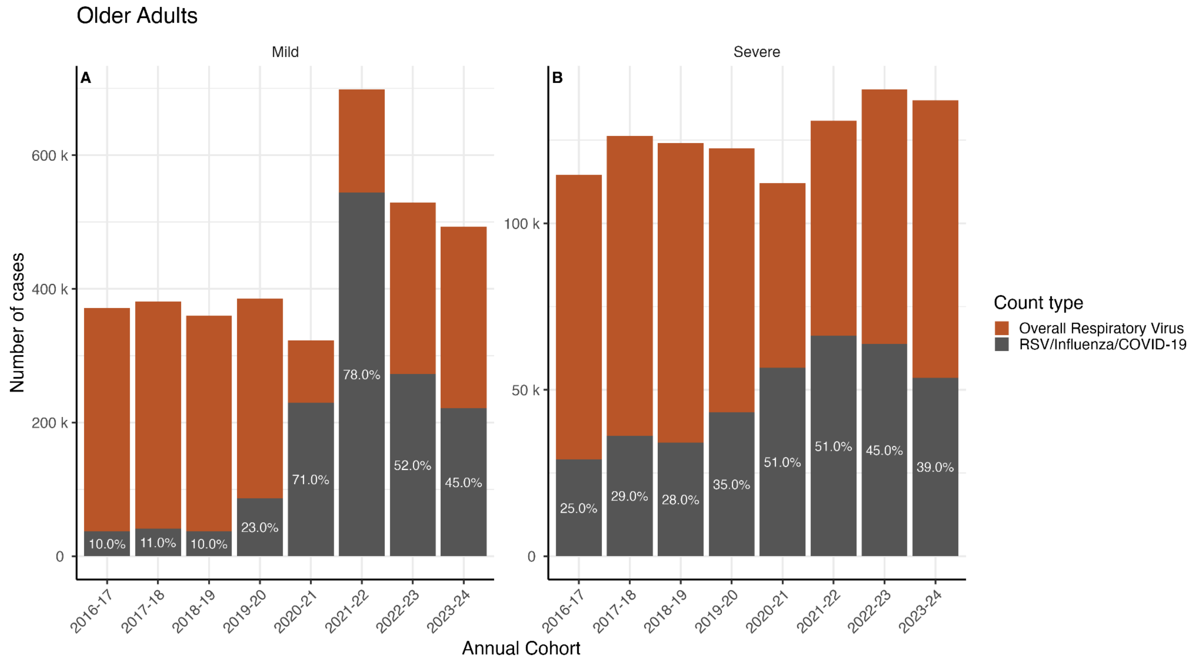

Figure S36: Number of cases in older adults classified as either RSV/influenza/COVID-19 using broad phenotypes (grey bar) or as overall respiratory virus (orange bar). Values indicate the percentage of overall respiratory virus cases classified as RSV/influenza/COVID-19. Results are shown by case severity and annual cohort.

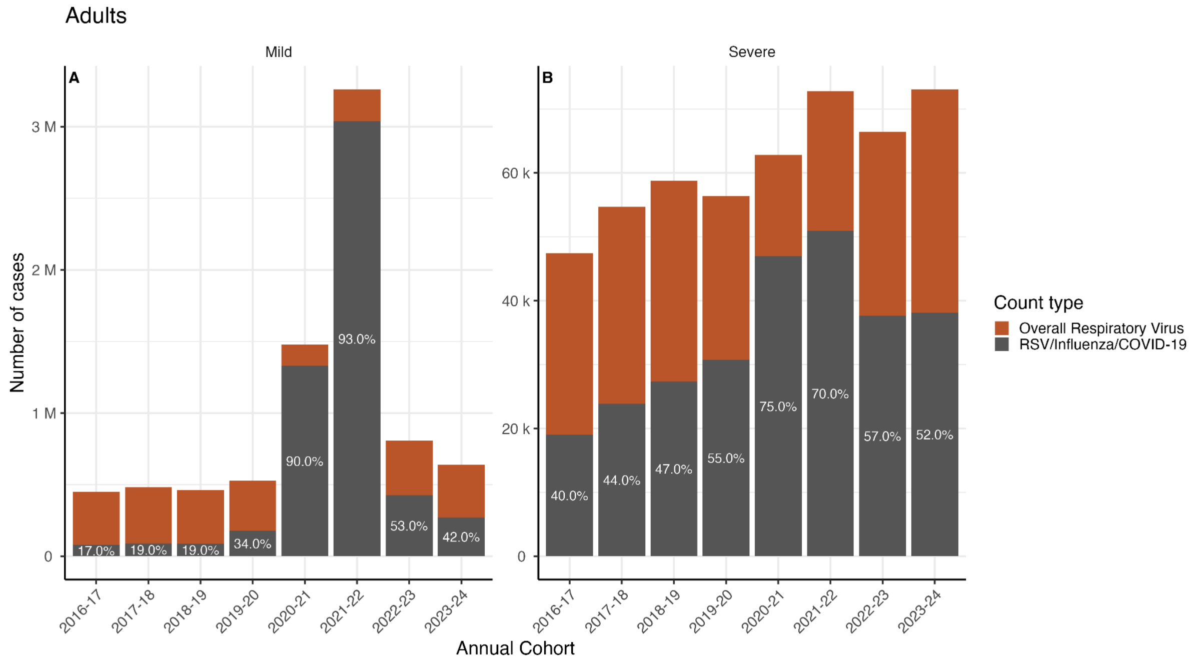

Figure S37: Number of cases in adults classified as either RSV/influenza/COVID-19 using broad phenotypes (grey bar) or as overall respiratory virus (orange bar). Values indicate the percentage of overall respiratory virus cases classified as RSV/influenza/COVID-19. Results are shown by case severity and annual cohort.

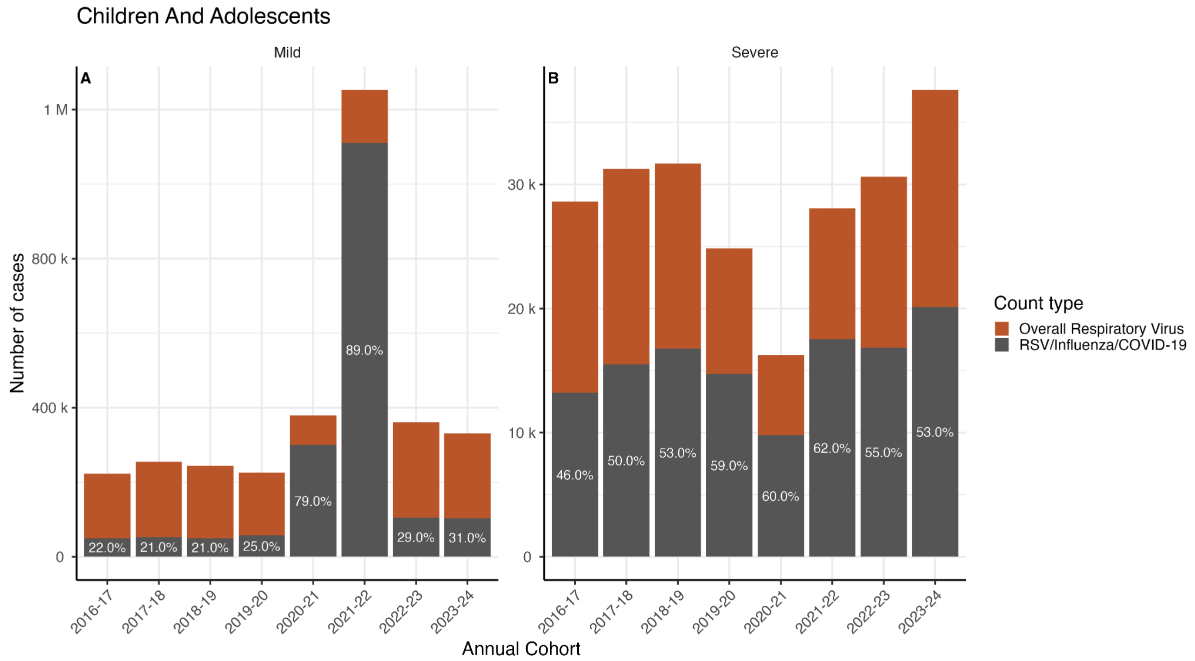

Figure S38: Number of cases in children and young people classified as either RSV/influenza/COVID-19 using broad phenotypes (grey bar) or as overall respiratory virus (orange bar). Values indicate the percentage of overall respiratory virus cases classified as RSV/influenza/COVID-19. Results are shown by case severity and annual cohort.

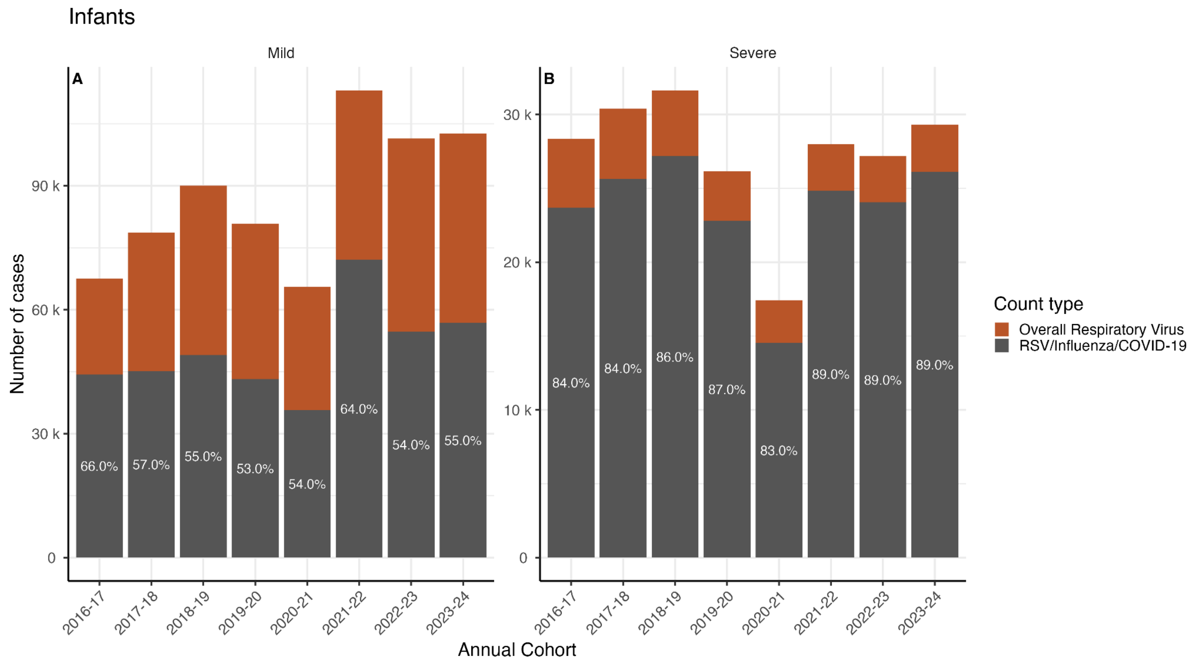

Figure S39: Number of cases in infants classified as either RSV/influenza/COVID-19 using broad phenotypes (grey bar) or as overall respiratory virus (orange bar). Values indicate the percentage of overall respiratory virus cases classified as RSV/influenza/COVID-19. Results are shown by case severity and annual cohort.

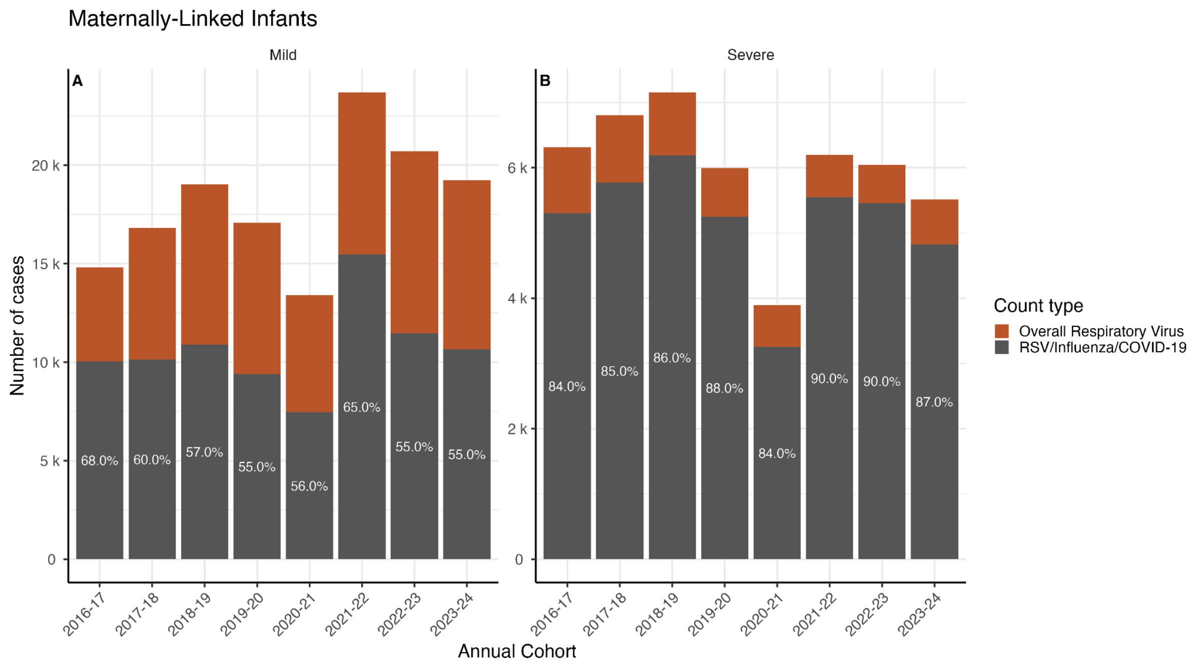

Figure S40: Number of cases in maternally-linked infants classified as either RSV/influenza/COVID-19 using broad phenotypes (grey bar) or as overall respiratory virus (orange bar). Values indicate the percentage of overall respiratory virus cases classified as RSV/influenza/COVID-19. Results are shown by case severity and annual cohort.

### S4. Comparisons to Surveillance

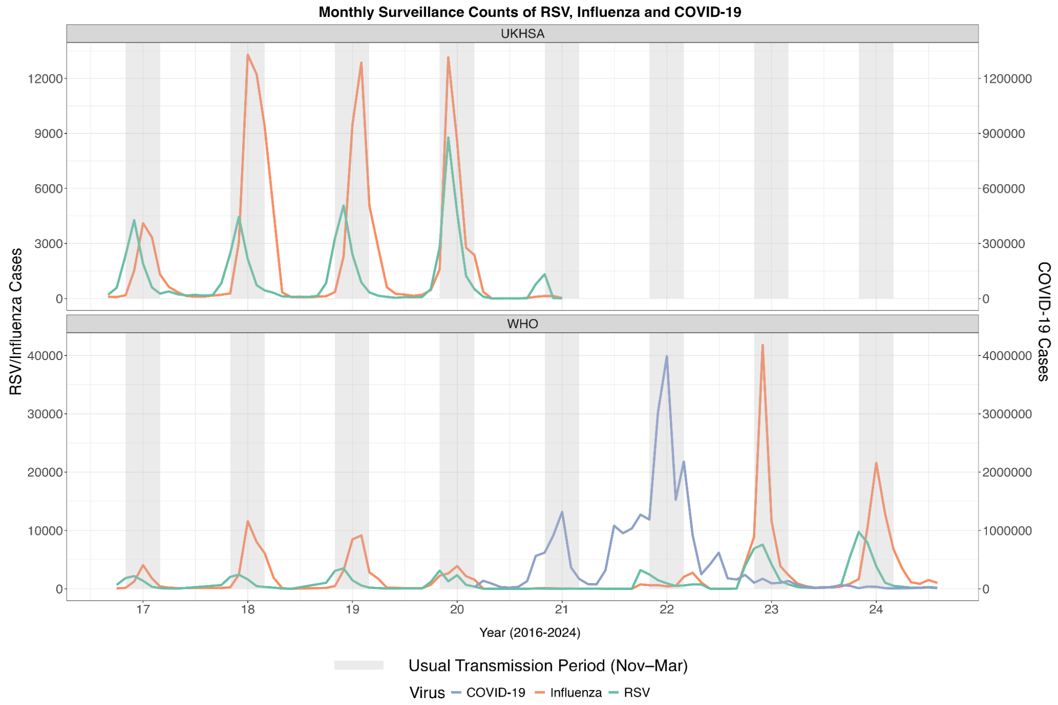

Figure S41: Comparison of monthly case counts from two sources of surveillance data, UKHSA (top panel) and WHO (bottom panel). UKHSA data was available from 2016-2020 for RSV (green) and influenza (orange), WHO data was available from 2016-2024 for RSV (green), influenza (orange), and COVID-19 (purple).

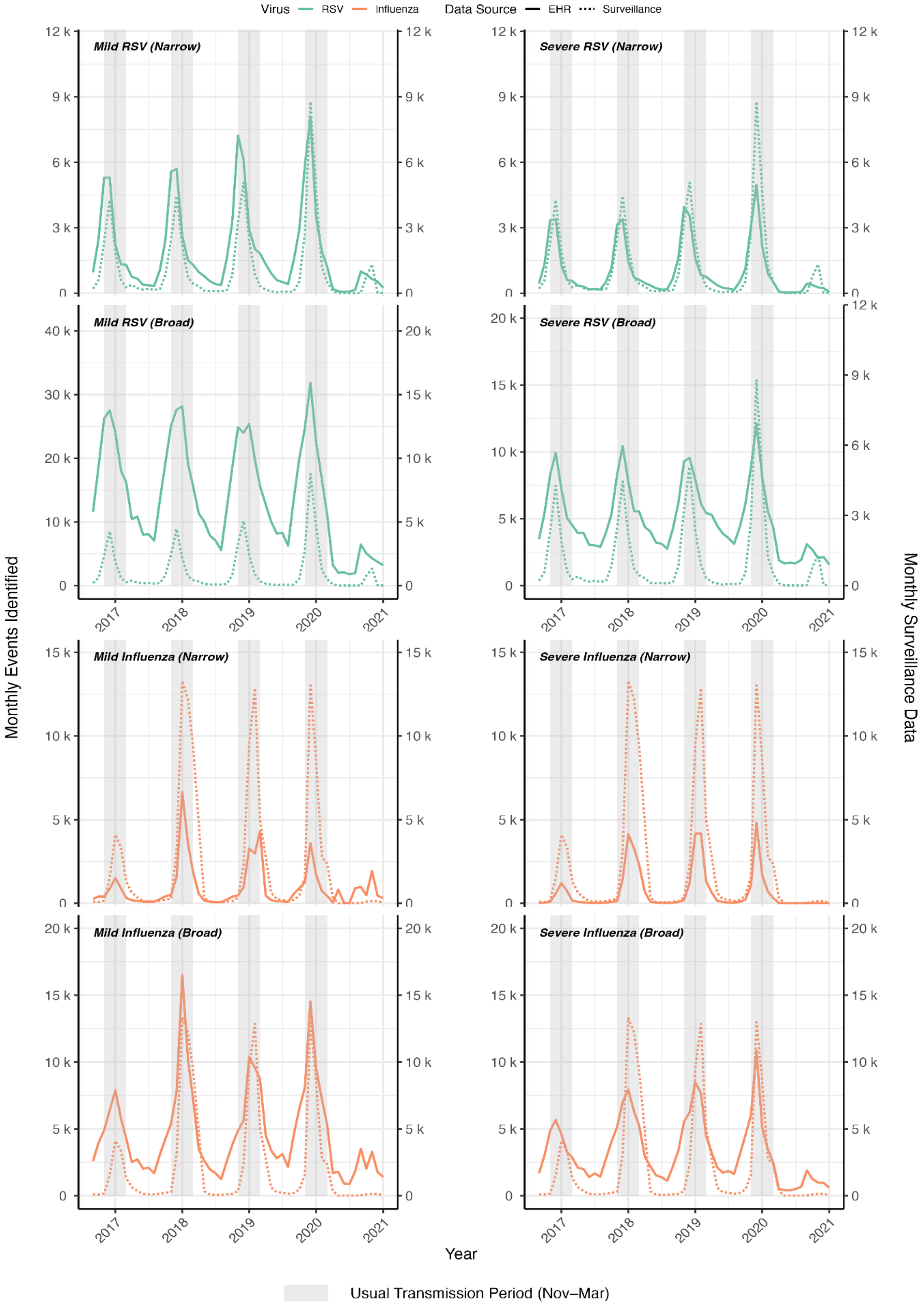

Figure S42: Comparison of cases identified via OpenSAFELY (EHR Data) across all ages, left hand axis, and surveillance data (UKHSA), right hand axis. Cases are shown separately for mild (left hand side) and severe (right hand side), with narrow cases appearing above broad cases for each virus respectively - where RSV is shown in green and influenza is shown in orange. Surveillance data is indicated by dotted lines, with phenotyped cases indicated by solid lines.

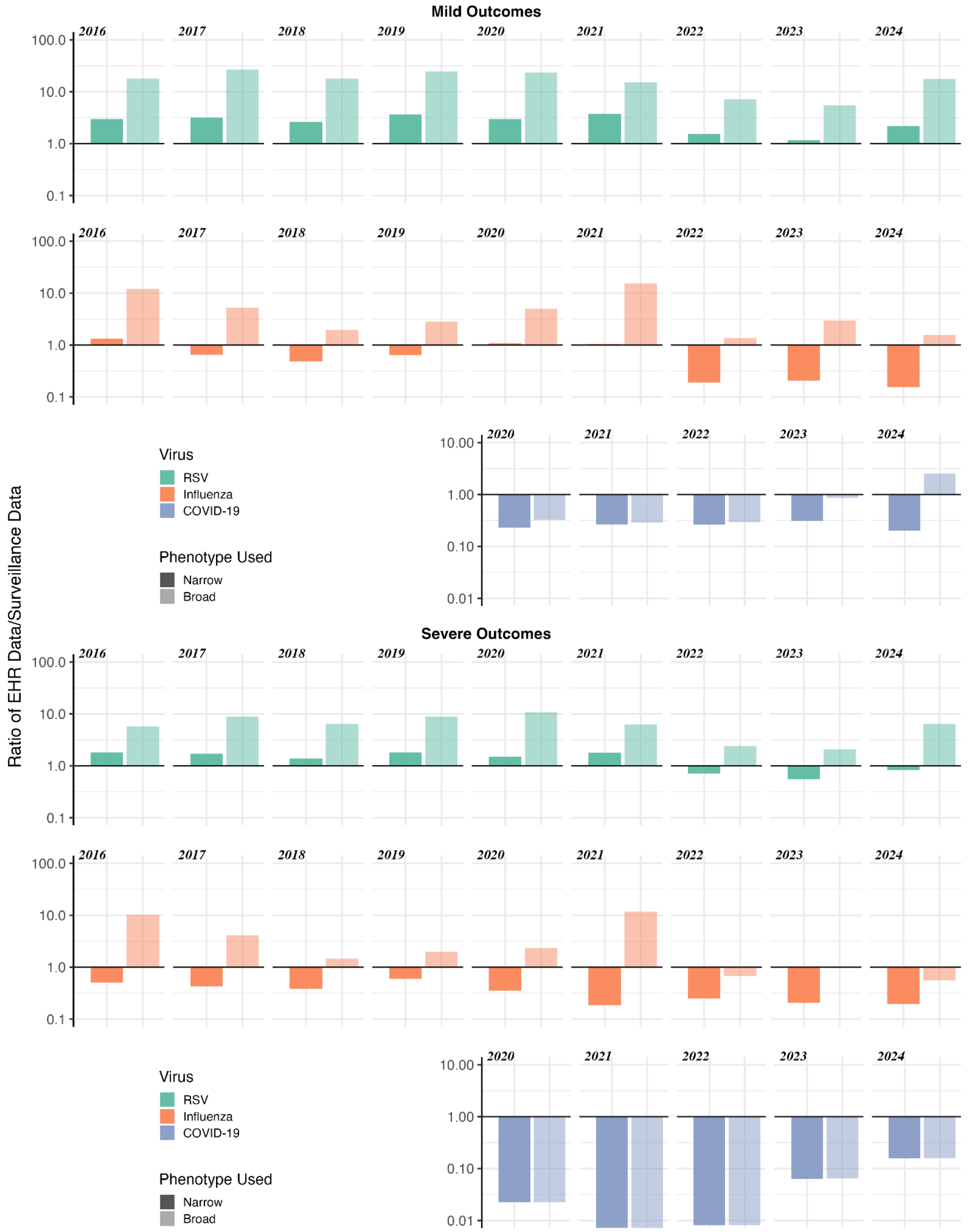

Figure S43: Ratio of EHR data over WHO RespiMart surveillance data; where bars below y = 1 indicate more cases reported in surveillance data and bars above y =1 indicate more cases identified from EHR data. Bars are shaded to indicate the virus (RSV in green; influenza in orange; COVID-19 in purple) and phenotype (narrow phenotypes in darker shades and broad phenotypes in lighter shades).

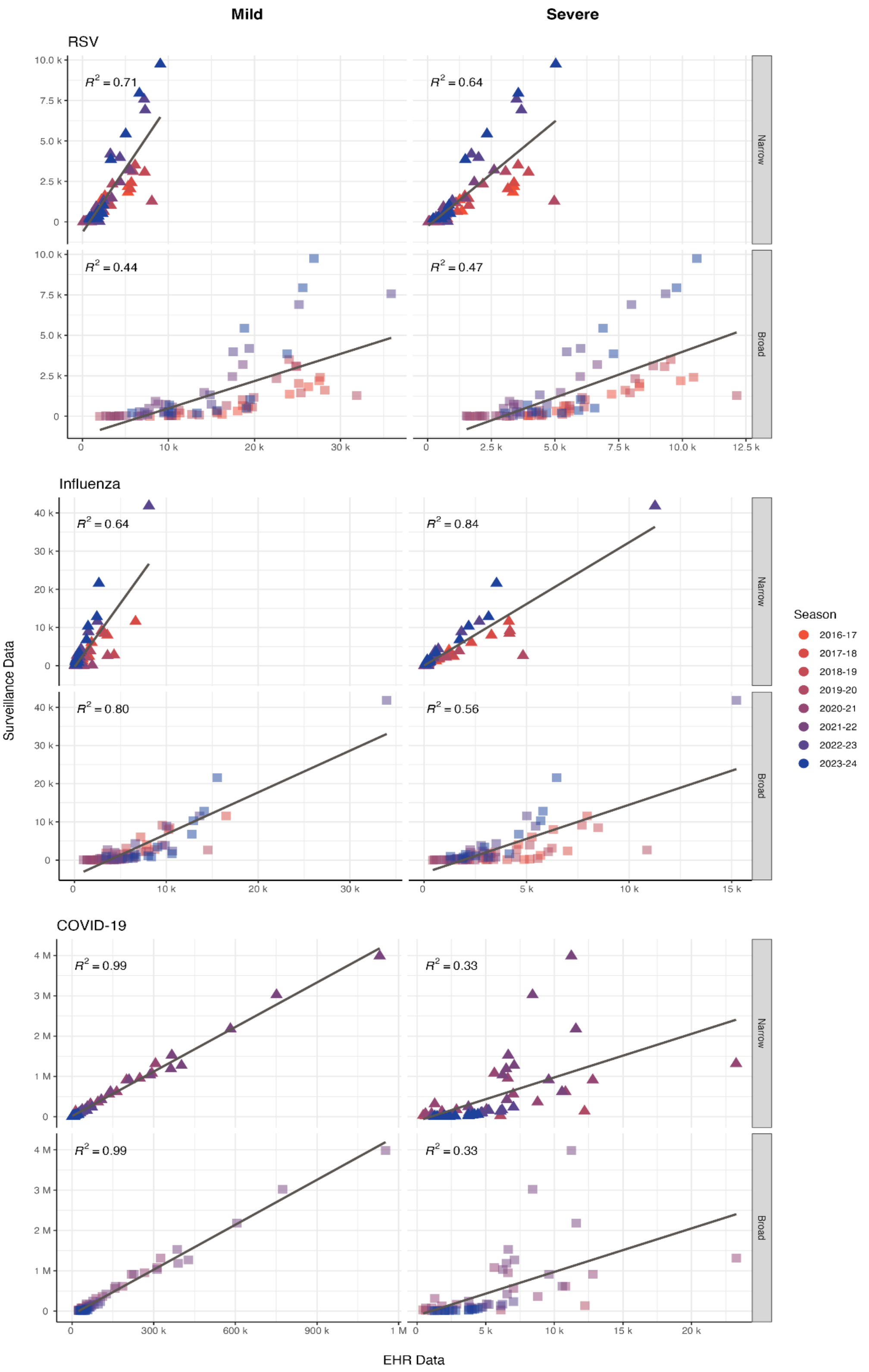

Figure S44: Comparison of cases identified via OpenSAFELY (EHR Data) across all ages, x-axis, and surveillance data (WHO RespiMart), y-axis. Surveillance data for RSV and influenza are for England and for COVID-19 are for the entire UK. Monthly case counts are shown separately for mild (left hand side) and severe (right hand side), with narrow cases appearing darker than broad cases for each virus respectively - where each annual cohort is coloured differently according to a gradient. Included on the plot is the value of $R^2$ which is estimated for the overall relationship across available annual cohorts for each case respectively.

| Case | Phenotype | Correlation Coefficient (95% confidence interval) | | | | | | | |
|---|---|---|---|---|---|---|---|---|---|
| | Annual Cohort | 2016-17 | 2017-18 | 2018-19 | 2019-20 | 2020-21 | 2021-22 | 2022-23 | 2023-24 |
| Mild RSV | Narrow | 0.96 (0.85, 0.99) | 0.96 (0.87, 0.99) | 0.96 (0.88, 0.99) | 0.77 (0.35, 0.93) | -0.55 (-0.85, 0.04) | 0.71 (0.22, 0.91) | 0.97 (0.89, 0.99) | 0.98 (0.93, 0.99) |
| Mild RSV | Broad | 0.93 (0.76, 0.98) | 0.9 (0.69, 0.97) | 0.83 (0.48, 0.95) | 0.79 (0.4, 0.94) | -0.5 (-0.83, 0.11) | 0.81 (0.43, 0.94) | 0.95 (0.81, 0.98) | 0.81 (0.43, 0.94) |
| Severe RSV | Narrow | 0.96 (0.85, 0.99) | 0.97 (0.88, 0.99) | 0.98 (0.93, 0.99) | 0.73 (0.27, 0.92) | -0.54 (-0.85, 0.05) | 0.6 (0.05, 0.87) | 0.98 (0.93, 0.99) | 0.98 (0.93, 0.99) |
| Severe RSV | Broad | 0.98 (0.93, 1) | 0.97 (0.89, 0.99) | 0.94 (0.79, 0.98) | 0.79 (0.39, 0.94) | -0.53 (-0.85, 0.06) | 0.78 (0.36, 0.93) | 0.97 (0.91, 0.99) | 0.9 (0.68, 0.97) |
| Mild Influenza | Narrow | 0.96 (0.85, 0.99) | 0.97 (0.88, 0.99) | 0.77 (0.35, 0.93) | 0.67 (0.15, 0.9) | 0.74 (0.29, 0.92) | 0.23 (-0.39, 0.71) | 1 (0.99, 1) | 0.96 (0.87, 0.99) |
| Mild Influenza | Broad | 0.85 (0.55, 0.96) | 0.94 (0.78, 0.98) | 0.91 (0.71, 0.98) | 0.85 (0.54, 0.96) | 0.05 (-0.54, 0.6) | 0.11 (-0.5, 0.64) | 0.98 (0.94, 1) | 0.85 (0.55, 0.96) |
| Severe Influenza | Narrow | 0.98 (0.94, 1) | 0.99 (0.97, 1) | 1 (0.99, 1) | 0.68 (0.17, 0.9) | -0.36 (-0.78, 0.27) | 0.95 (0.82, 0.99) | 1 (0.99, 1) | 0.97 (0.89, 0.99) |
| Severe Influenza | Broad | 0.58 (0, 0.86) | 0.78 (0.37, 0.93) | 0.89 (0.64, 0.97) | 0.74 (0.3, 0.92) | -0.68 (-0.9, -0.17) | 0.09 (-0.51, 0.63) | 0.99 (0.98, 1) | 0.88 (0.62, 0.97) |
| Mild COVID-19 | Narrow | | | | 0.77 (-0.1, 0.97) | 0.99 (0.97, 1) | 0.99 (0.97, 1) | 0.99 (0.97, 1) | 0.95 (0.84, 0.99) |
| Mild COVID-19 | Broad | | | | 0.85 (0.14, 0.98) | 0.99 (0.97, 1) | 0.99 (0.97, 1) | 0.98 (0.94, 1) | 0.66 (0.15, 0.9) |
| Severe COVID-19 | Narrow | | | | 0.83 (0.07, 0.98) | 0.79 (0.41, 0.94) | 0.62 (0.08, 0.88) | 0.94 (0.8, 0.98) | 0.91 (0.72, 0.98) |
| Severe COVID-19 | Broad | | | | 0.83 (0.06, 0.98) | 0.79 (0.41, 0.94) | 0.62 (0.07, 0.88) | 0.94 (0.8, 0.98) | 0.91 (0.71, 0.98) |

Table S14: Pearson's correlation coefficient test results with corresponding correlation coefficient for each case and phenotype combination. The results are for each annual cohort separately, for each virus/severity/phenotype respectively
